# MOATAI-VIR - an AI algorithm that predicts severe adverse events and molecular features for COVID-19’s complications

**DOI:** 10.1101/2021.01.29.21250712

**Authors:** Courtney Astore, Hongyi Zhou, Joshy Jacob, Jeffrey Skolnick

**Affiliations:** Center for the Study of Systems Biology, School of Biological Sciences, Georgia Institute of Technology, 950 Atlantic Drive, N.W., Atlanta, GA 30332, USA; Emory Vaccine Center, Emory University, Atlanta, GA30329, USA; Yerkes National Primate Research Center, Emory University, Atlanta, GA30329, USA; Department of Microbiology and Immunology, Emory Vaccine Center, School of Medicine, Emory University, Atlanta, GA 30329, USA

## Abstract

Following SARS-CoV-2 infection, some COVID-19 patients experience severe adverse events caused by pathogenic host responses. To treat these complications, their underlying etiology must be identified. Thus, a novel AI-based methodology, **MOATAI-VIR**, which predicts disease-protein-pathway relationships for 22 clinical manifestations attributed to COVID-19 was developed. SARS-CoV-2 interacting human proteins and GWAS identified respiratory failure associated risk genes provide the input from which the mode-of-action (MOA) proteins/pathways of the resulting disease comorbidities are predicted. These comorbidities are then mapped to their clinical manifestations. Three uncharacterized manifestation categories are found: neoplasms, mental and behavioral disorders, and congenital malformations, deformations, and chromosomal abnormalities. The prevalence of neoplasms suggests a possible association between COVID-19 and cancer, whether by shared molecular mechanisms between oncogenesis and viral replication, or perhaps, SARS-CoV-2 is an oncovirus. To assess the molecular basis of each manifestation, the proteins shared across each group of comorbidities were prioritized and subject to global pathway analysis. From these most frequent pathways, the molecular features associated with hallmark COVID-19 phenotypes, such as loss of sense of smell/taste, unusual neurological symptoms, cytokine storm, and blood clots were explored. Results of **MOATAI-VIR** are available for academic users at: http://pwp.gatech.edu/cssb/MOATAI-VIR/.

## Introduction

The COVID-19 pandemic is caused by SARS-CoV-2, a positive-sense, single-stranded, rapidly mutating RNA coronavirus(1). The societal impact of COVID-19 is amplified by the minority of individuals experiencing significant complications/death following infection. Some victims experience acute respiratory distress syndrome(2), while others have clotting issues, cytokine storms, hypoxemia, low white blood cell counts, and bone marrow failure(3-6). Despite the development of COVID-19 vaccines, until they are widely administered, there will be new cases of COVID-19 with complications including loss of smell/taste and/or unusual neurological symptoms (7, 8). Thus, systematic approaches are desperately needed to combat COVID-19’s catastrophic long-term effects(9).

The primary objective of this work is to provide the molecular mechanisms for each clinical manifestation attributed to COVID-19. We define clinical manifestations as in Ref.(10) which provides 30 respiratory and non-respiratory COVID-19 in-hospital clinical complications (excluding the “other” category. To assist in the effort to further define these complications, a systematic method to identify comorbid diseases and the molecular features of these complications is preferred over a random, anecdotal approach.

To address this pressing need, we describe a new algorithm, **MOATAI-VIR, M**ode **O**f **A**ction proteins & **T**argeted therapeutic discovery driven by **A**rtificial **I**ntelligence for **VIR**uses designed to predict the mode-of-action (MOA) proteins of COVID-19s severe patient responses based on predicted COVID-19 disease comorbidities. To accomplish this, we independently input either the experimentally determined human-SARS-CoV-2 interactome or COVID-19 GWAS survival-associated risk genes in a MOA indication profile(5). These profiles are used to determine the disease comorbidities associated with the MOA proteins presumably causing a particular complication. In practice, each COVID-19 comorbid disease is mapped to its respective clinical manifestation group. Then, the top comorbidity enriched MOA proteins are subject to pathway analysis to identify the molecular processes underlying the etiology of each complication. There were also comorbidities for both the interactome and GWAS sets that did not map to a characterized COVID-19 clinical manifestation group; one example is cancers. These “uncharacterized” complications were grouped by their ICD-10 main classification(11). We next determined the enriched MOA proteins and performed global pathway analysis.

## Results

### Prediction of COVID-19’s complications and underlying molecular mechanisms

An overview of the **MOATAI-VIR** approach is shown in Figure 1, with a more detailed flowchart in Figure S1. The goal of **MOATAI-VIR** is to identify severe adverse responses associated with SARS-CoV-2 and their corresponding human mode-of-action proteins. To accomplish these objectives, we input either the experimentally determined human proteins from the human-SARS-CoV-2 interactome(6) or COVID-19 GWAS survival associated risk genes(2, 5) as a MOA profile in **MEDICASCY**. We then employ our recently developed **LeMeDISCO** algorithm which predicts disease comorbidity. **LeMeDISCO** then outputs indications that likely cause the adverse events due to SARS-CoV-2 infection. The set of top identified proteins is then passed to CoPathway and the pathways are ranked by their statistical significance in LIST_CoPathway_.

**Figure 1.**
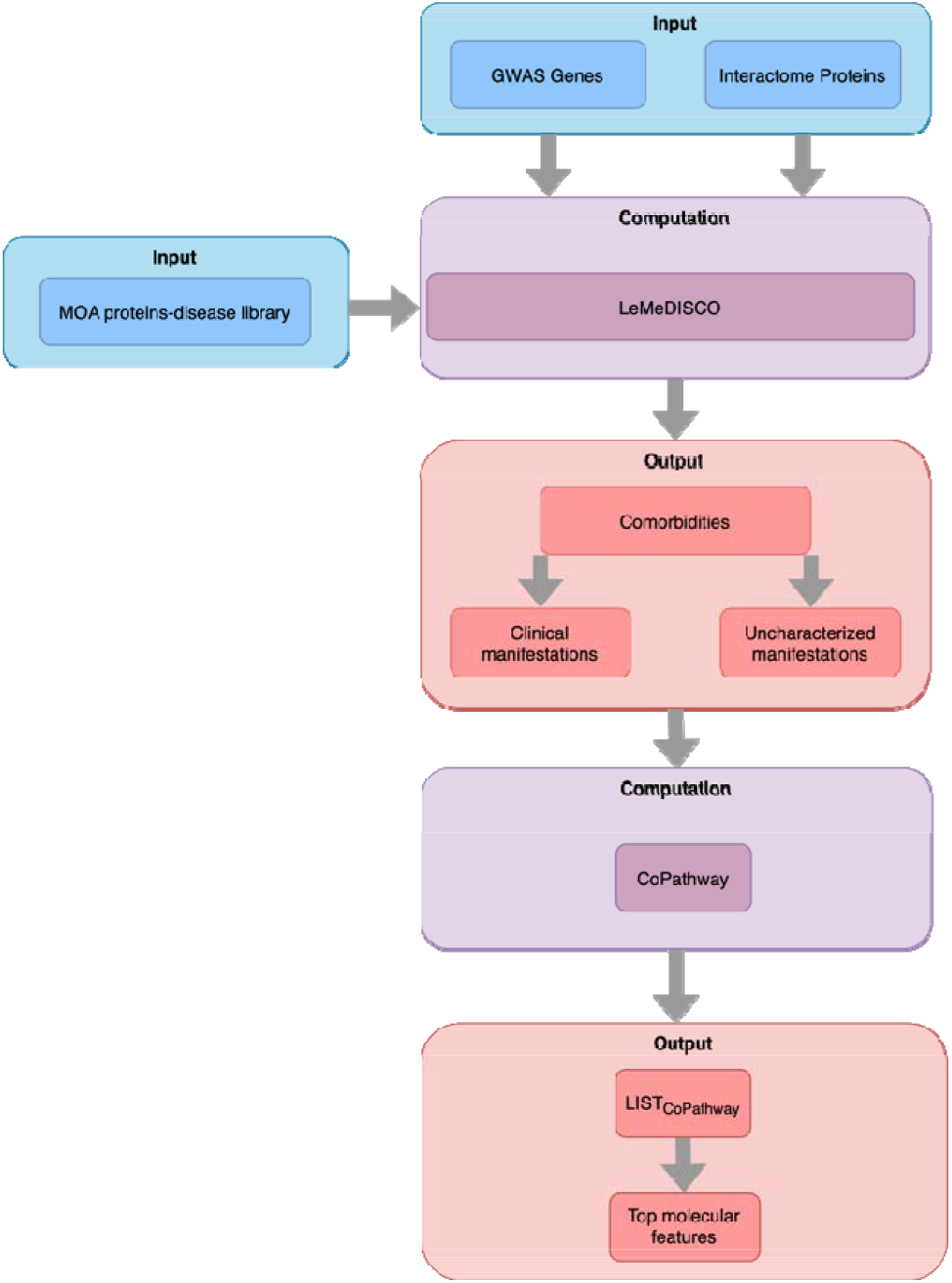
Overview of the **MOATAI-VIR** approach that predicts comorbid human diseases, their MOA proteins and molecular features associated with COVID-19’s severe secondary adverse events. Blue are **MOATAI-VIR** inputs, purple are computational algorithms and pink are output predictions.

### Large Scale Benchmarking of MEDICASCY MOA Predictions

The first step uses **MEDICASCY**(12) to predict MOA protein targets of all diseases. For large scale benchmarking for MOA prediction, we mapped all drugs in our indication library to DrugBank drugs(13) (v5.09) and obtained their respective human protein targets. These are combined with those from the Therapeutic Target Database(14). Using the drug-indication relationships in our training library of **MEDICASCY**, we compiled indication-protein target relationships of 145,722 pairs for 3,539 indications (with an average ∼41 proteins/indication) for benchmarking. In benchmarking, any drug in the training library having a Tanimoto coefficient, TC(15) ≥ 0.8 to the given drug whose indications are predicted is excluded from training. We define a MOA prediction for an indication when its p-value < 0.001 using the upper tailed null hypothesis. 61.0% of predicted protein-indication prediction are correctly predicted. If a training TC=1 cutoff is used, this increases to 76.9%.

### Large Scale Benchmarking of LeMeDISCO

As shown and discussed in SI, Table S0, large scale benchmarking of **LeMeDISCO** on different clinical data sets shows high comorbid disease coverage and accuracy compared to alternative methods(16-19). It is also superior to alternatives that rely solely on symptom data, which lack molecular mechanism-based associations and cannot provide information of confidence-ranked putative protein targets. **LeMeDISCO**’s recall rate is close to 70% for a representative set of 2,630 disease pairs.

### COVID-19’s Clinical Manifestations

We first predicted COVID-19 comorbidities using the 332 high confidence human proteins that interact with SARS-CoV-2(6). There are 697 significant comorbidities (with a Z-score cutoff > 1.65 corresponding to a p-value of 0.05 using the upper tailed null hypothesis). The top five disease comorbidities ranked by their Z-score that mapped to a COVID-19 clinical manifestation group are shown in Table 1, with an expanded list in Table S1. Also provided are comorbidity enriched protein targets. Without extrinsic information or training, these results recapitulate many key COVID-19 phenotypes such as myelosuppression, immunodeficiency, neurotoxicity, blood indications, myocardial infarctions, stroke and cytokine storm symptoms(2, 6, 20). The ICD-10 code of the comorbid diseases was used to map them to the 30 complications listed in Ref.(10). After ICD-10 mapping, we manually examine the unmapped ones and search for literature evidence of a possible match. For example, we do not automated have an ICD-10 match of Fallot’s tetralogy (ICD-10:Q21) to the Cardiovascular complication(ICD-10: I00-I99). Since it is a congenital heart disease, it is manually assigned to this complication. Manually mapped indications are ∼10% of the mapped ones. Thus, even if there are few errors, it will not significantly affect the final results. In practice, indications are mapped into 17 of the 30 COVID-19 complications (see Table 1). Our library of 3,608 indications(21) does not have any matches to 4/30 complications (Dialysis initiation, Intracranial hemorrhage, Hypertensive crisis, Cardiogenic shock). Effectively, the complication recall rate is 17/26∼65%. These mapped indications are then used to prioritize MOA proteins, pathways.

**Table 1.**
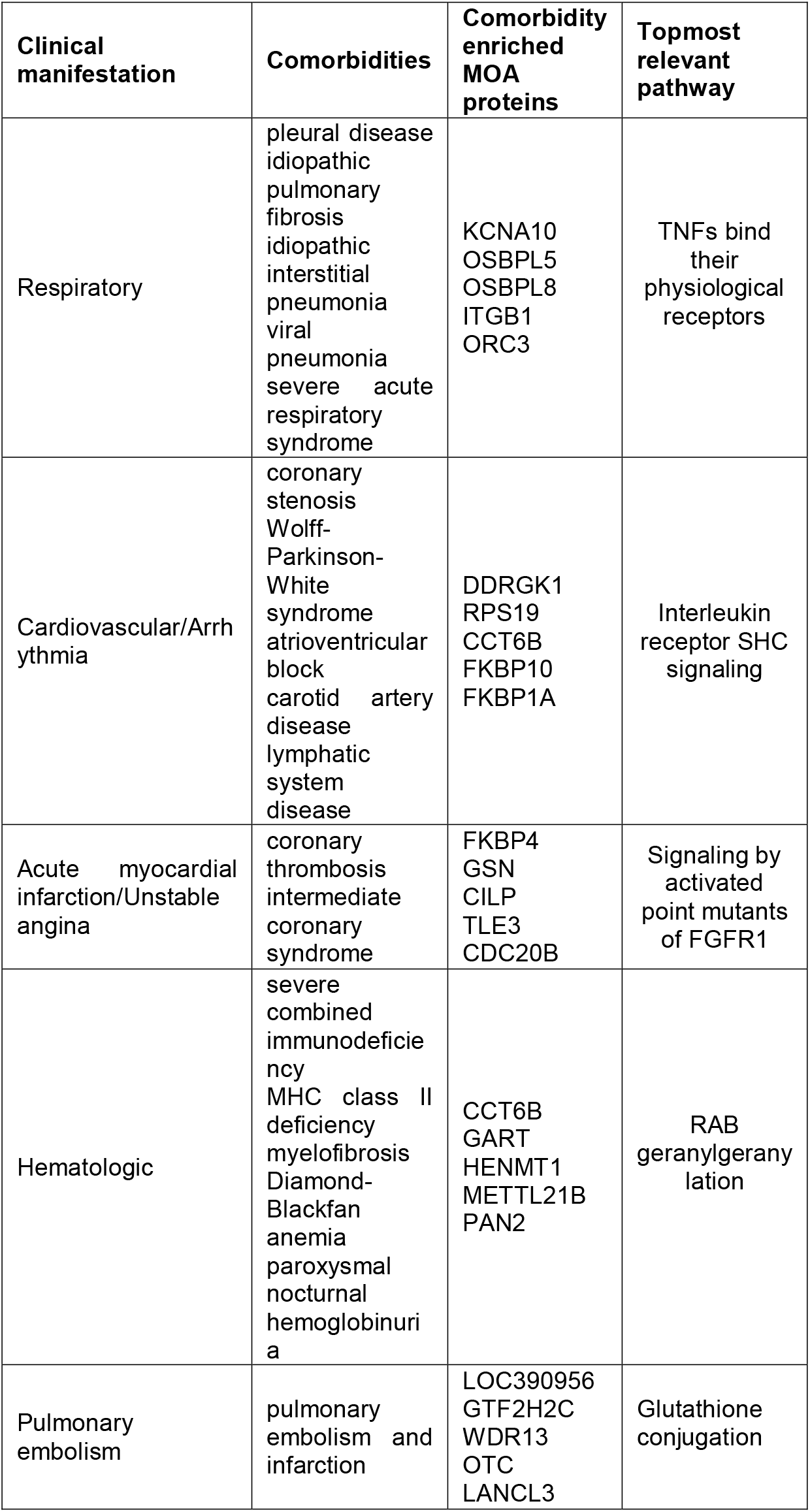
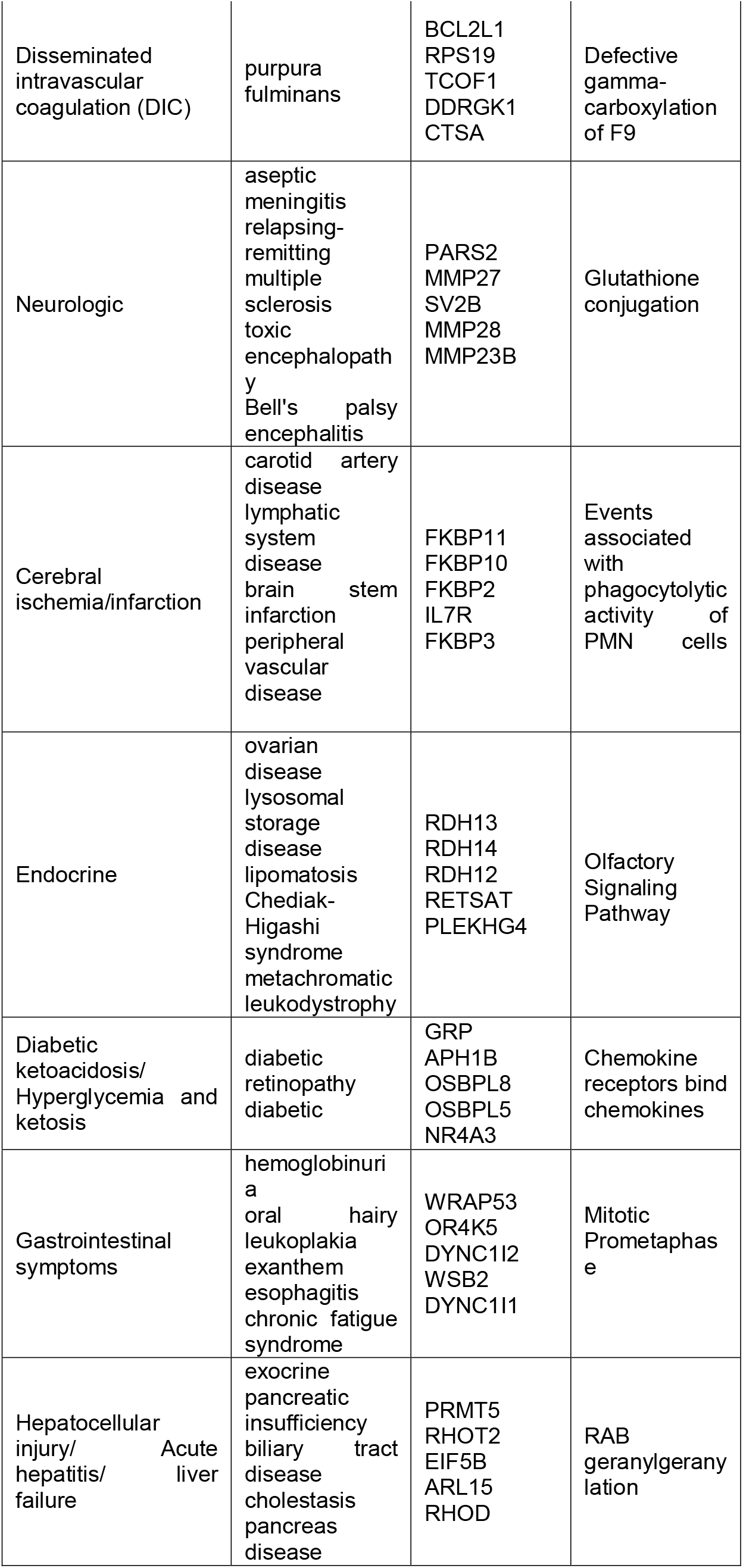
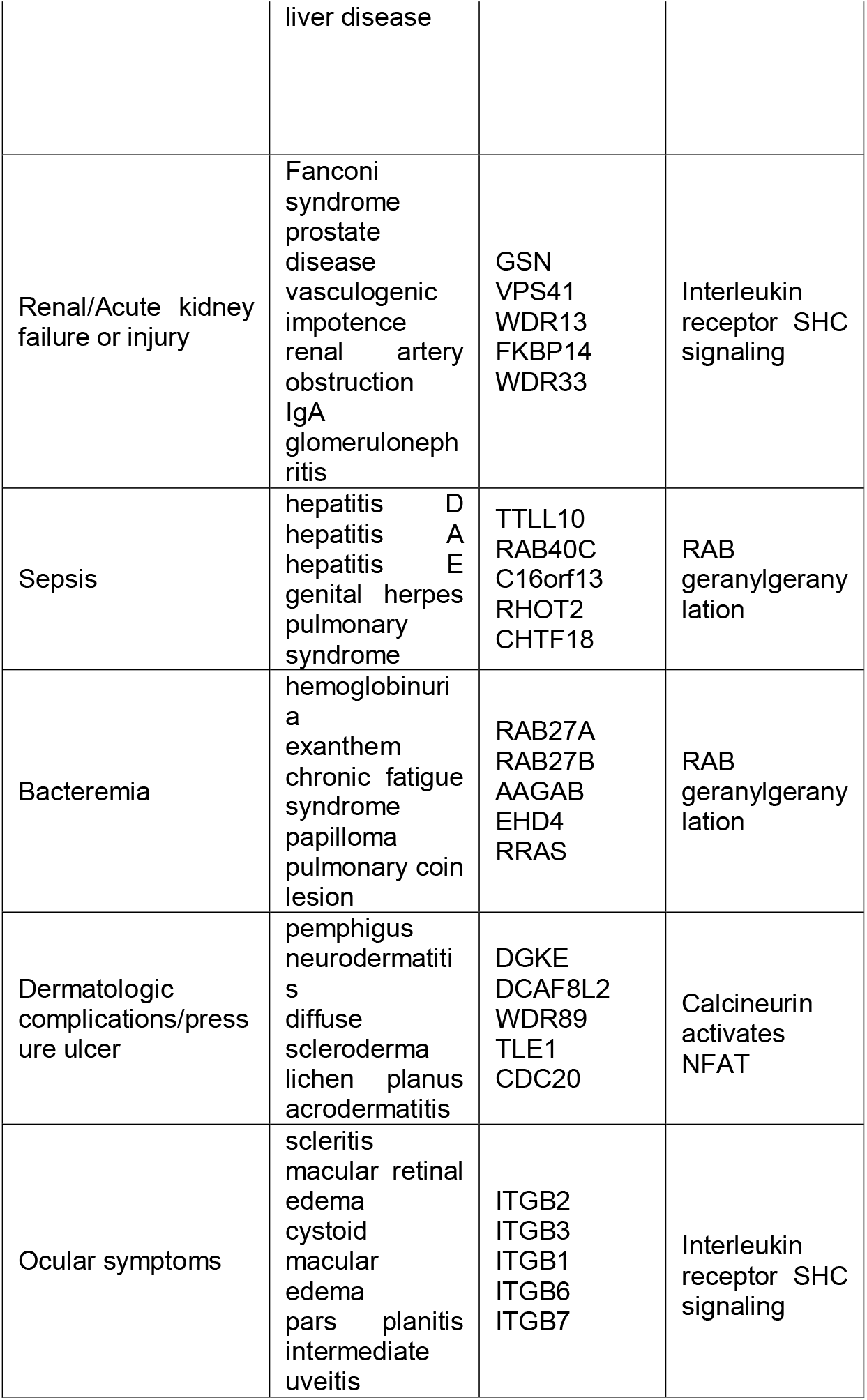
Top 5 comorbidities, top 5 comorbidity enriched MOA proteins, and topmost relevant pathway for each COVID-19 severe adverse clinical manifestations predicted using the SARS-CoV-2 interactome as input

The 6 human genes near the 3p21.31 locus of human genome identified in a GWAS study as being strongly associated with respiratory failure in COVID-19 patients(5) (odds ratio 1.77) were next used to predict comorbidities. We determined 380 significant comorbidities having a Z-score >1.65. Strikingly, as shown in Table 2, many severe clinical complications associated with COVID-19 are in the top-ranked comorbidity predictions including respiratory complications, myocardial infarction and cytokine storms. Table S2 provides an expanded list that includes myocardial infarction, stroke, neurological manifestations, hearing disorders, hypoxemia, lung, cardiovascular and diabetic risk factors(2, 20, 22). Excluding 4 indications not in our library, with GWAS risk gene input, the recall rate of the COVID-19 complications is 19/26∼73%. Since the 332 proteins of the SARS-CoV-2 human interactome and the 6 GWAS COVID-19 survival risk genes that do not overlap except for FYCO1, their comorbidity predictions are complementary. Combining both the human interactome and GWAS complications provides a prediction of 22/26 (84%) of COVID-19’s complications.

**Table 2.**
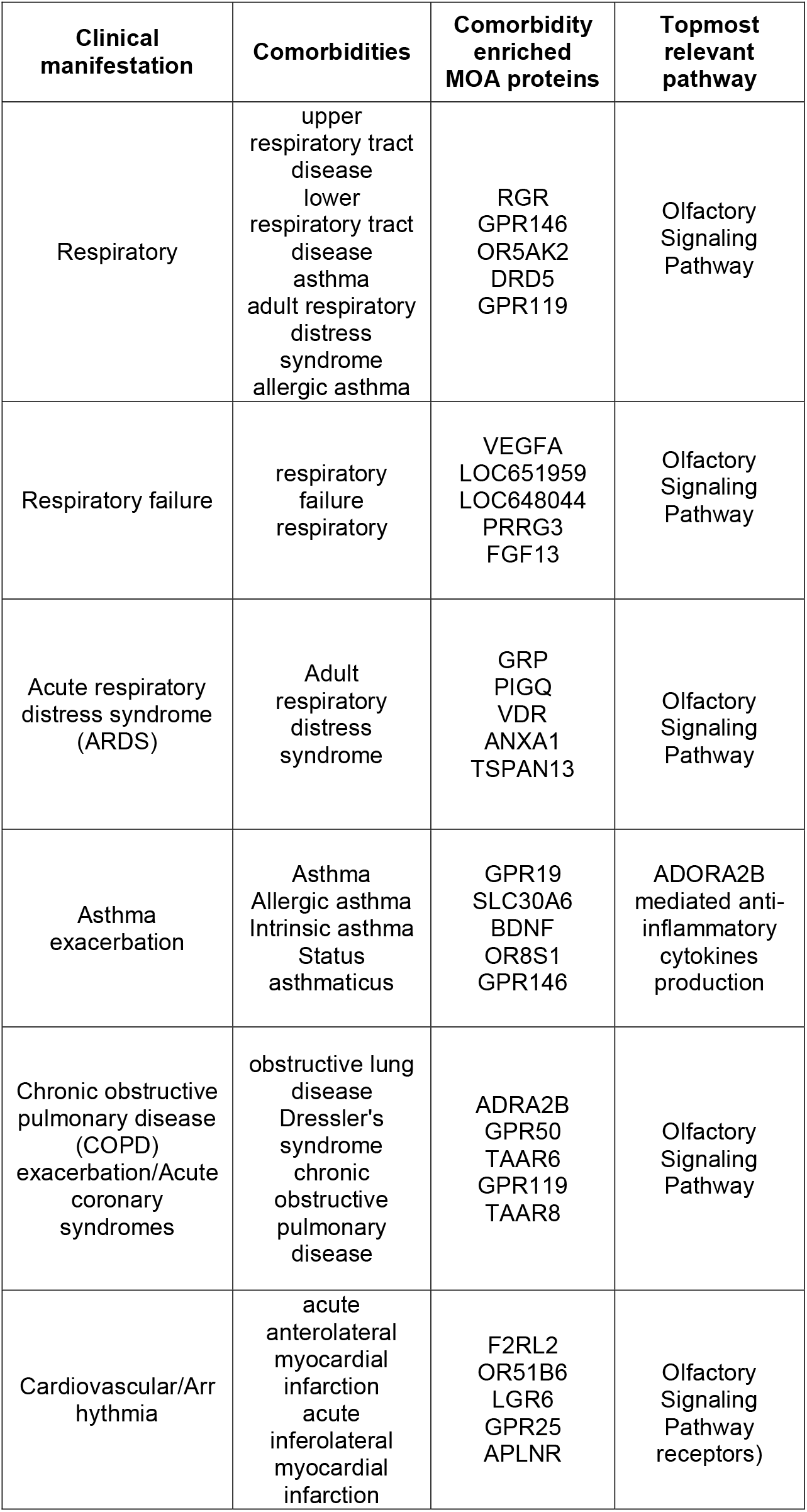
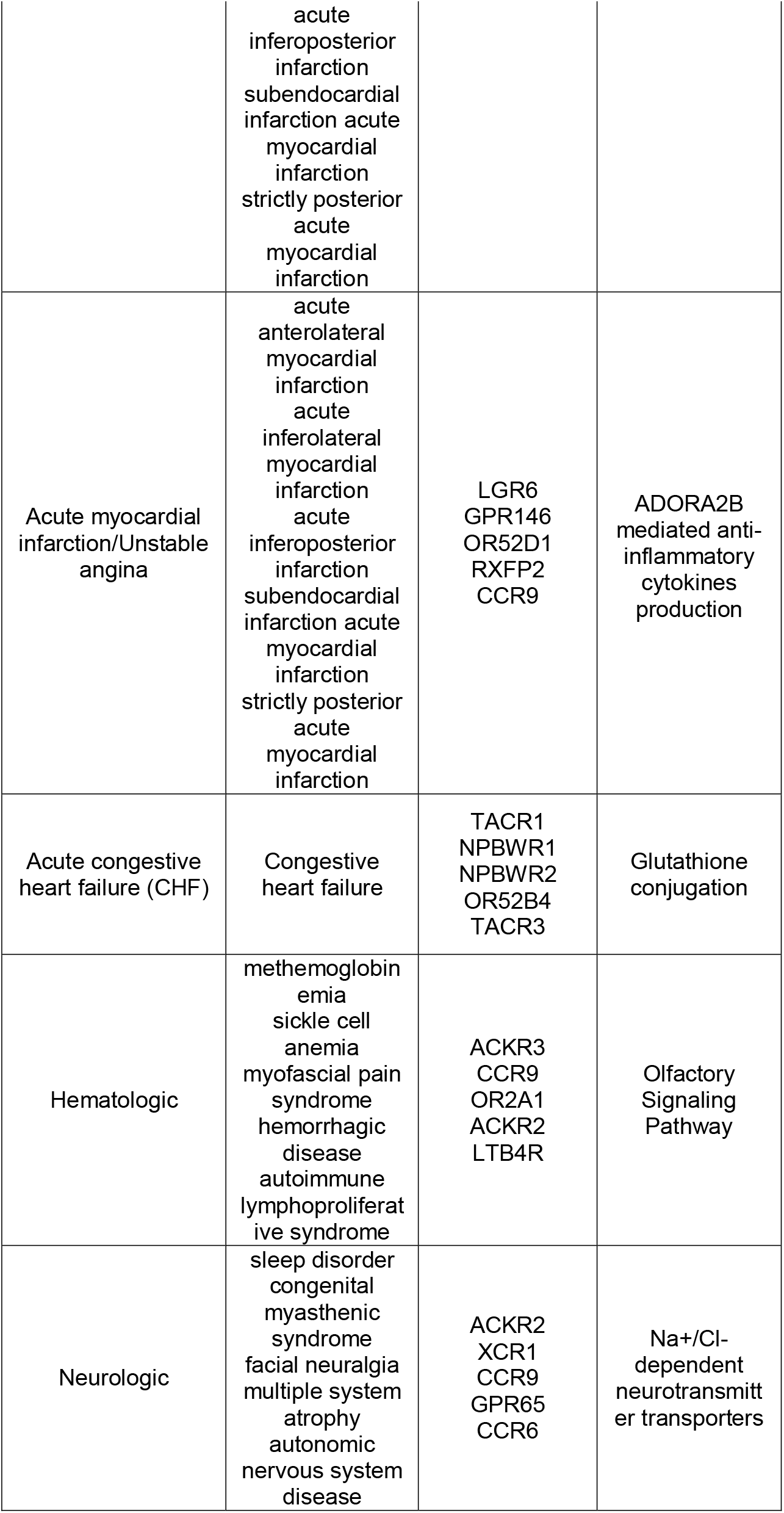
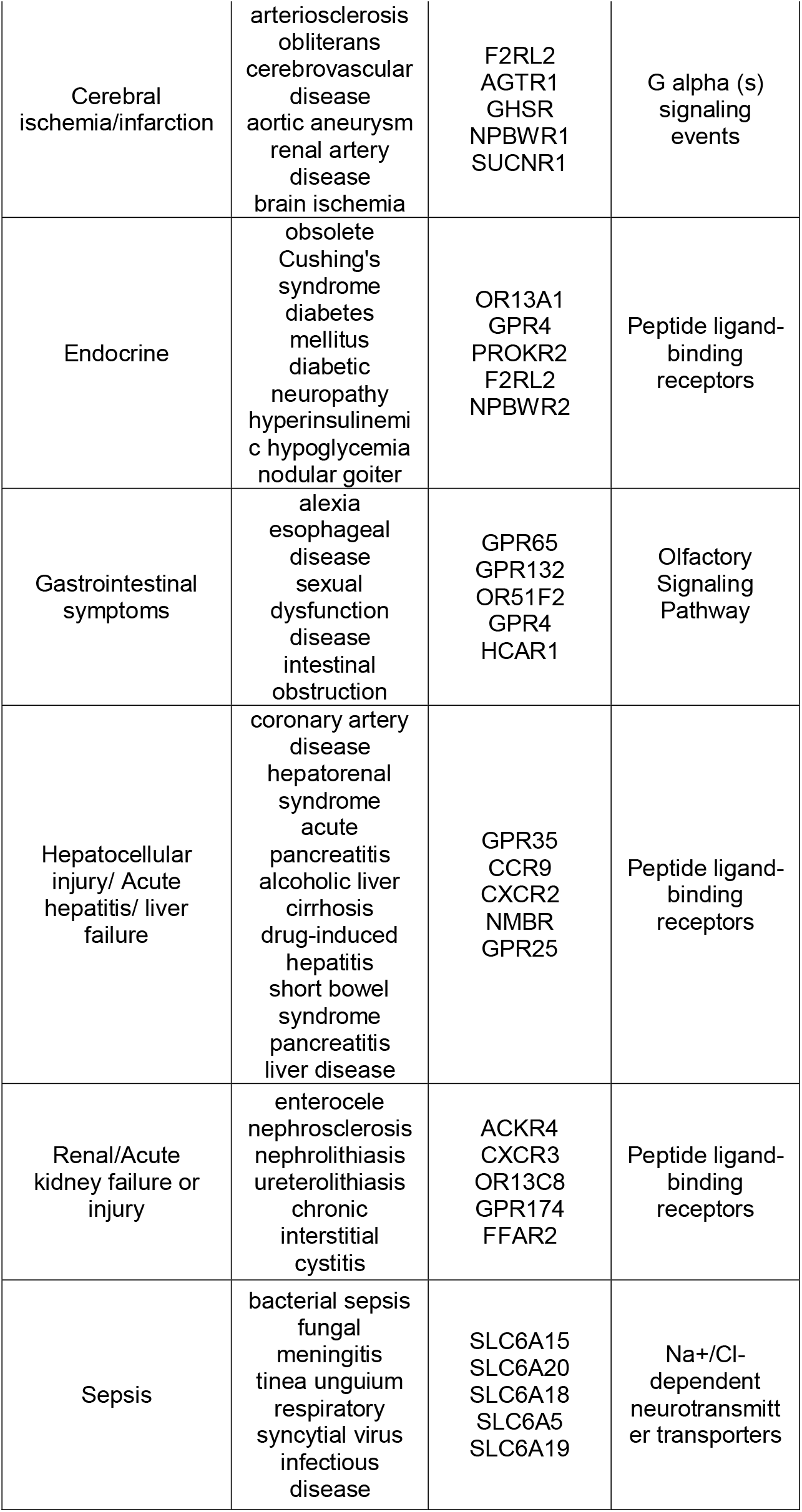
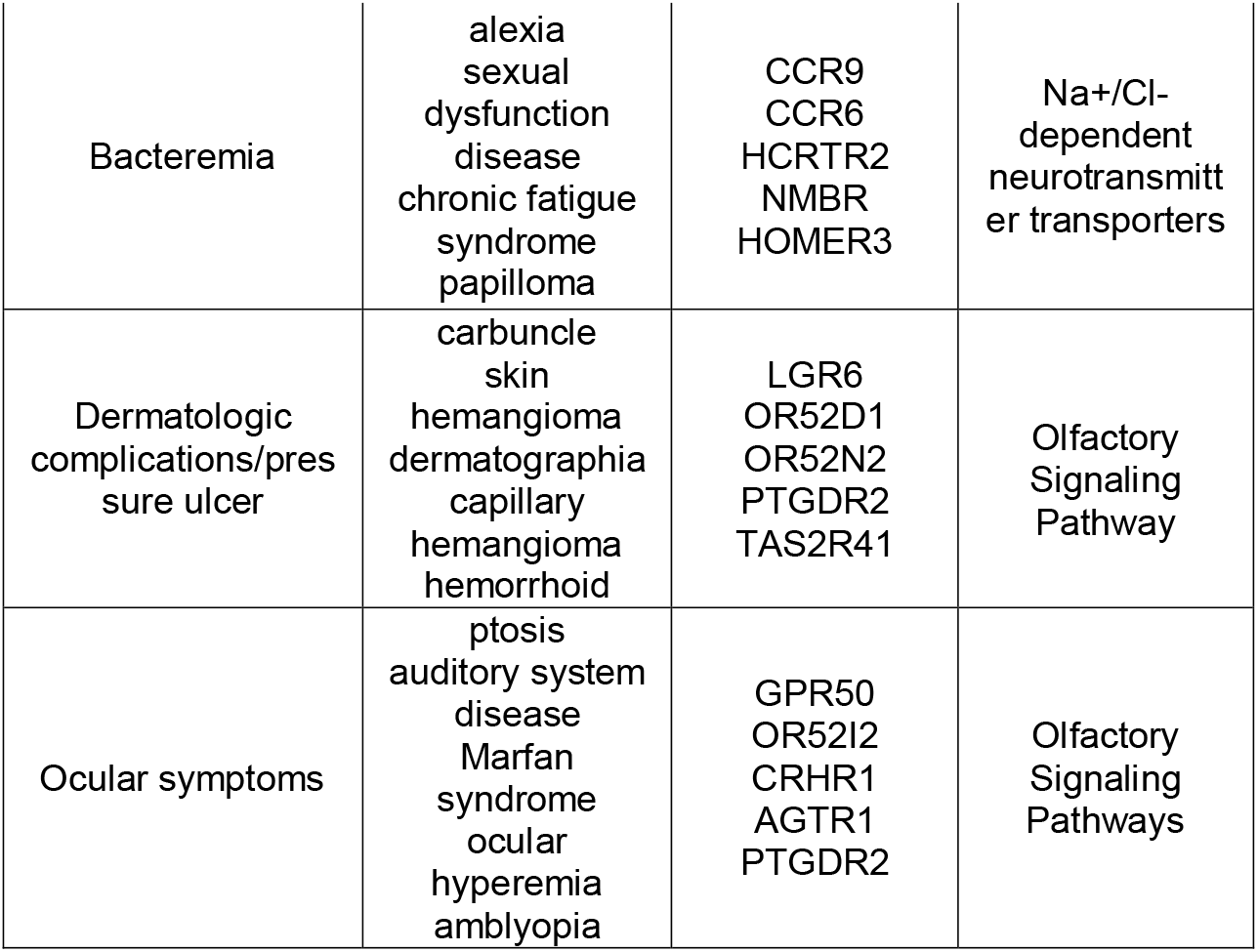
Top 5 comorbidities, top 5 comorbidity enriched MOA proteins, and topmost relevant pathway for each COVID-19 severe adverse clinical manifestations predicted using GWAS input

### Diseases comorbid with COVID-19

From the significant comorbidities found from the interactome data, there were 297 neurologic, 41 hematologic, 34 ocular, 26 endocrine, 14 gastrointestinal, 13 cardiovascular, 11 respiratory, 9 renal failure, 9 sepsis, 6 cerebral ischemia, 6 dermatological, 5 bacteremia, 5 hepatocellular, 2 acute myocardial infarction, 1 diabetic ketoacidosis, 1 Disseminated intravascular coagulation (DIC), and 1 pulmonary embolism diseases. From the GWAS data, there were 48 neurologic, 47 cardiovascular, 28 ocular, 27 respiratory, 26 endocrine, 17 gastrointestinal, 13 cerebral ischemia, 10 hepatocellular injury, 9 renal failure, 8 acute myocardial infarction, 7 dermatological, 6 bacteremia, 5 hematologic, 4 sepsis, 4 asthma exacerbation, 3 chronic obstructive pulmonary disease (COPD), 1 acute congestive heart failure (CHF), 1 acute respiratory distress syndrome (ARDS), and 1 respiratory failure diseases.

### Pathway analysis

The top 100 comorbidity enriched MOA proteins for each complication with n > 1 diseases were used as input into global pathway analysis. If there was only 1 disease in a complication, all of the **MEDICASCY** significantly predicted MOA proteins for that disease were used as inputs into the global pathway analysis. A top important pathway, the interactome and GWAS as input results are shown in Tables 1 and 2, with full lists in Tables S1 and S2 respectively.

The top 20 most frequent significant pathways across the clinical manifestations calculated from the interactome and GWAS inputs are shown in Tables 3 and 4, respectively. Combining this with the hierarchically ranked pathways for each clinical manifestation allowed us to further identify hallmark pathways attributed with unusual COVID-19 symptoms such as loss of sense of smell, cytokine storms, blood clots, and unusual neurological symptoms.

**Table 3.** Top 20 most frequent pathways across the interactome clinical manifestations.

| **Pathway\|Top pathway** | **Frequency of clinical manifestations** |
| --- | --- |
| Association of TriC/CCT with target proteins during biosynthesis\|Metabolism of proteins | 9 |
| Interleukin receptor SHC signaling\|Immune System | 7 |
| TGFBR1 LBD Mutants in Cancer\|Disease | 6 |
| Synthesis of very long-chain fatty acyl-CoAs\|Metabolism | 6 |
| RHO GTPases Activate Formins\|Signal Transduction | 6 |
| Nuclear Receptor transcription pathway\|Gene expression | 6 |
| Elastic fibre formation\|Extracellular matrix organization | 6 |
| RAB geranylgeranylation\|Metabolism of proteins | 5 |
| N-Glycan antennae elongation\|Metabolism of proteins | 5 |
| Molecules associated with elastic fibres\|Extracellular matrix organization | 5 |
| Keratan sulfate/keratin metabolism\|Metabolism | 5 |
| Keratan sulfate biosynthesis\|Metabolism | 5 |
| Interleukin-4 and Interleukin-13 signaling\|Immune System | 5 |
| HSP90 chaperone cycle for steroid hormone receptors (SHR)\|Cellular responses to external stimuli | 5 |
| HSF1-dependent transactivation\|Cellular responses to external stimuli | 5 |
| Attenuation phase\|Cellular responses to external stimuli | 5 |
| Acyl chain remodeling of PS\|Metabolism | 5 |
| The NLRP1 inflammasome\|Immune system | 4 |
| Sodium/Calcium exchangers\|Transport of small molecules | 4 |
| Reduction of cytosolic Ca<sup>++</sup> levels\|Hemostasis | 4 |

**Table 4.** Top 20 most frequent pathways across the GWAS clinical manifestations.

| **Pathway\|Top pathway** | **Frequency of clinical manifestations** |
| --- | --- |
| Signaling by GPCR\|Signal Transduction | 19 |
| Peptide ligand-binding receptors\|Signal Transduction | 19 |
| GPCR downstream signaling\|Signal Transduction | 19 |
| G alpha (s) signaling events\|Signal Transduction | 19 |
| Class A/1 (Rhodopsin-like receptors)\|Signal Transduction | 19 |
| Olfactory Signaling Pathway\|Signal Transduction | 18 |
| GPCR ligand binding\|Signal Transduction | 18 |
| ADORA2B mediated anti-inflammatory cytokines production\|Disease | 18 |
| G alpha (q) signaling events\|Signal Transduction | 16 |
| Signal Transduction\|Signal Transduction | 14 |
| Leishmania parasite growth and survival\|Disease | 14 |
| G alpha (i) signaling events\|Signal Transduction | 14 |
| Anti-inflammatory response favoring Leishmania parasite infection\|Disease | 14 |
| Leishmania infection\|Disease | 12 |
| Amine ligand-binding receptors\|Signal Transduction | 12 |
| Adrenoceptors\|Signal Transduction | 12 |
| Relaxin receptors\|Signal Transduction | 11 |
| Chemokine receptors bind chemokines\|Signal Transduction | 11 |
| Tachykinin receptors bind tachykinins\|Signal Transduction | 10 |
| Glucagon-type ligand receptors\|Signal Transduction | 10 |

#### Loss of sense of smell

The olfactory signaling pathway is associated with 18/19 clinical manifestation groups from the GWAS results. This pathway involves enzymatic activity that initiates the binding of an odorant molecule to a receptor resulting in an electrical signal that is transmitted to the brain(23). The olfactory signaling pathway was the top pathway for the asthma exacerbation, hematologic, respiratory failure, and gastrointestinal GWAS clinical manifestations. There is clinical evidence that some individuals infected with SARS-CoV-2 experience a loss of smell and taste(23). Furthermore, it has been found that olfactory receptors may be an alternative SARS-CoV-2 entry into the local host cells, which may lead to its spread into the central nervous system(24).

#### Neurological symptoms

There have been a number of unusual COVID-19-related neurological symptoms reported, such as stroke, confusion, and as previously mentioned, loss of sense of smell and taste(25). The most frequent clinical manifestation group for the interactome results was neurologic. The top neurologic pathway was glutathione conjugation associated with facilitating the metabolism of xenobiotics. Dysregulation of glutathione plays a role in a wide variety of diseases including neurodegenerative diseases and cancer(26, 27). Decreased glutathione levels can lead to oxidative stress which may result in Parkinson’s and Alzheimer’s disease. Moreover, an imbalance in glutathione levels can impact the immune system(27). A decreased glutathione concentration is highly associated with serious manifestations causing increased COVID-19 mortality, which may be due to the increased susceptibility to uncontrolled viral replication(28).

The most frequent clinical manifestation group from the GWAS results was also neurological. The top pathway was Na+/Cl-dependent neurotransmitter sodium symporters, which use sodium and chloride electrochemical gradients to import/export a number of substrates. They are associated with Parkinson’s disease, orthostatic intolerance, and depression(29). A meta-analysis found that low blood sodium increases the risk and severity of COVID-19(30). Thus, neurotransmitter transporters that depend on Na+ could be dysregulated due to decreased levels of blood sodium.

### Blood clotting

Increased blood clots have been seen in individuals infected with COVID-19. A meta-analysis found that individuals infected by COVID-19 with blood clots have increased mortality(31). The second most frequent clinical manifestation group for the interactome results was hematologic. The top hematologic pathway was RAB geranylgeranylation, the eighth most frequent pathway across all clinical manifestations from the interactome. RAB geranylgeranylation is a post-translational modification that allows RABs to connect with intracellular membranes where they regulate vesicle transport pathways(32). Dysregulation of RAB geranylgeranylation transferase function has been linked to abnormal blood clotting.

### Cytokine storm

There have been a number of COVID-19 patients that face ARDS, which may be due to a cytokine storm elicited the nuclear factor kappa B (NF-κB) pathway(33). The second most frequent clinical manifestation group for the GWAS results was cardiovascular. The top pathway was G alpha (s) signaling events, which covered all GWAS clinical manifestation groups. This pathway’s primary function is to activate adenylate cyclase producing cAMP. G-protein receptors are associated with heart disease. Among the proteins in this pathway, C5aR1 is a G-protein-coupled receptor. The C5a-C5aR1 complex is involved in COVID-19 progression and is part of a potential therapeutic strategy(34).This complex is associated with the innate immune response, with C5 a key driver in complement-mediated inflammation(34). Other cytokine storm-related pathways covering a majority of clinical manifestation groups for the interactome results include interleukin receptor SHC signaling, Interleukin-4 and Interleukin-13 signaling(35). ADORA2B mediated anti-inflammatory cytokines production and chemokine receptors bind chemokines are also prevalent pathways.

### Uncharacterized Manifestations

There was a total of 234 significant comorbidities that were not mapped to a known COVID-19 clinical manifestation group from the interactome results. To further understand their effects, we grouped them by their main ICD-10 classification and performed **CoPathway** analysis on the top two groups to further understand their molecular bases. The top two uncharacterized groups were Neoplasms and Congenital malformations, deformations, and chromosomal abnormalities. The comorbidities, comorbidity enriched MOA proteins and pathways for the interactome results are shown in SI, Table S3.

A total of 155 diseases were not mapped to a clinical manifestation group from the GWAS results. The comorbidities, comorbidity enriched MOA proteins and pathways for the GWAS results are shown in Table S4. Again, to further understand these potentially important COVID-19 adverse events, we grouped them by their main ICD-10 classification and performed **CoPathway** analysis on the top two groups. The top two uncharacterized groups were Neoplasms and Mental and behavioral disorders. There were 37 pathways with a p-value < 0.05. The top pathway was amine-ligand binding receptors, class A GPCRs that bind to biogenic amine ligands. These amines can act as neurotransmitters. A study found that specific trace amines are highly prevalent in patients with mental disorders(36). To the best of our knowledge, there has been no research investigating the relationship between amine-ligand binding receptors and COVID-19 as a suitable biomarker for therapeutic intervention; however, there are numerous COVID-19 associated mental and behavioral occurrences(37).

### Neoplasms

From interactome, there were 90 pathways with a p-value < 0.05 involving neoplasms, with many involving hormonal regulation. The top pathway involves activation of AMPK downstream of NMDARs, that is associated with the neuronal system. AMPK is an enzyme that regulates cellular energy and homeostasis via activating catabolic pathways while switching off cellular growth and proliferation(38). AMPK has been targeted for cancer treatment because its activation can reduce cancer incidence. NMDARs are vital in controlling synaptic plasticity and memory. Increased expression of NMDARs occurs in a variety of cancers such as neuroblastoma, breast, small-cell lung, and ovarian cancer(39). Anti-NMDAR encephalitis, characterized by abnormal neurological and behavioral symptoms, has been reported in both COVID-19 and HSV(40). Notably, HSV can lead to an increased cervical cancer risk(41, 42).

From GWAS, there were 29 pathways for Neoplasms with a p-value < 0.05. The top pathway was the nuclear receptor transcription pathway. Nuclear receptors are DNA-binding transcription factors capable of binding hormones, vitamins, small molecules, and other ligands. There are a number of underlying disease mechanisms associated with dysregulation of nuclear receptors. This can yield a wide range of conditions such as cancer, diabetes, and hormone-related conditions. Nuclear receptors have been targeted by cancer therapeutics as they are key players in gene regulatory networks(43). There has not been substantial research on the relationship between nuclear receptors and COVID-19; but we note that some viruses target nuclear receptors as part of the viral replication process(44).

### Is SARS-CoV-2 an oncovirus?

As indicated above, there were a large number of disease comorbidities associated with neoplasms from both the interactome and GWAS results. Neoplasms cause abnormal tissue growth, a significant cancer characteristic. Perhaps, SARS-CoV-2 hijacks the human host replication machinery or proliferation pathways(6). Indeed, viruses can initiate signal transduction pathways leading to cytokine and chemokine expression. They also dysregulate signaling pathways to promote viral infection and cellular transformations(45) that elicit a proinflammatory response similar to that in cancer(46). A salient example is Human Papillomavirus (HPV). Most cervical cancers(47) are caused by the cytokine flux associated with inflammation post-HPV infection(42). Furthermore, the second most significant COVID-19 comorbid disease T-cell leukemia is linked to the human T-cell lymphotropic virus (HTLV-I), an RNA retrovirus. More generally, a number of oncoviruses cause cancer(48). Certain viruses transform human cells causing loss of ability to regulate cell division.

Although we still do not know the long-term consequences post-COVID-19 infection, these results raise the possibility that SARS-CoV-2 is an oncovirus. To assess this potential relationship, we screened our comorbidity enriched MOA proteins associated with Neoplasms from both the interactome and GWAS sets against the COSMIC(49) database gene set containing 723 oncogenes. We found that 47% (n=340) and 29% (n=208) of the MOA proteins from the interactome and GWAS Neoplasms are oncogenes in the COSMIC database(42) respectively; see SI Tables S5-6. For further validation, we compared the differential gene expression set from Table S4 of (50), which demonstrated the differential gene expression analysis of COVID-19 patients (n=1,918 differentially expressed genes with an adjusted p-value < 0.05), to the COSMIC database(49). From this, 11% (n=82) of the genes overlapped with the oncogenes in the COSMIC database (see SI Table S7). We next performed a 3-way merge between the interactome/GWAS Neoplasms comorbidity enriched MOA proteins, the SARS-CoV-2 differentially expressed genes, and the COSMIC database oncogenes. We found 40 and 23 overlapping genes from the interactome and GWAS 3-way merge, respectively. The pathway analysis on the overlapping COVID-19 differentially expressed genes and the COSMIC database indicates homogeneity of viral replication mechanisms and oncogenesis. A number of the identified pathways such as interferon-gamma signaling(51, 52), immunoregulatory interactions between lymphoid and non-lymphoid cell(53), and antigen processing-cross presentation(54, 55) are related to viral replication and oncogenesis. Possibly the machinery of viral replication and oncogenesis are similar. Clearly, additional investigation is needed to explore the possibility that SARS-CoV-2 is an oncovirus.

### MOATAI-VIR Website

The resulting data from **MOATAI-VIR** is provided to academic users at: http://pwp.gatech.edu/cssb/MOATAI-VIR/. Provided on the webpage are downloadable files for the significant diseases, proteins and pathways for the highlighted symptoms (e.g., loss of sense of smell) and clinical manifestations for interactome and/or GWAS. Furthermore, the significant diseases, proteins, and pathways for the summary and uncharacterized manifestation results for interactome and/or GWAS are also downloadable.

## Discussion

**MOATAI-VIR** has been shown to identify possible molecular mechanisms responsible for COVID-19’s severe adverse consequences. Most of COVID-19’s severe symptoms successfully predicted. These predicted comorbidities and the human MOA proteins possibly responsible for COVID-19’s complications will be validated in future work in cell lines and animal models. They will also be combined with antiviral drugs that directly target SARS-CoV-2 proteins to kill the virus. The goal is to mitigate both COVID-19 infection and subsequent adverse complications to improve clinical outcomes. Thus, **MOATAI-VIR** provides a series of logical, systematic choices designed to suggest treatments to COVID-19’s adverse reactions. Equally important, **MOATAI-VIR** is a general methodology can be applied to understand the possible etiology of human host response new outbreaks of other novel viral infections as they emerge. Finally, we note that an extension of **MOATAI-VIR** can be used to suggest repurposed drugs that modulate the host response to COVID-19’s complications.

## Materials and Methods

Additional details concerning **MOATAI-VIR** are provided in SI. Here, we note that **CoPathway** determines significant pathways associated with the most frequent comorbidity enriched MOA proteins. It first assesses the frequency of MOA proteins across the comorbidities for a desired group and then processing a top number (typically 100) of those MOA proteins through the Reactome(56) for global pathway analysis. Pathways with a p-value < 0.05 are then extracted and deemed the significant pathways portrayed in LIST_CoPathway._

## Data Availability

Results of MOATAI-VIR are available for academic users at: http://pwp.gatech.edu/cssb/MOATAI-VIR/.

## Acknowledgments

This project was funded by R35GM118039 of the Division of General Medical Sciences of the NIH.

