## Supplemental Information for "MOATAI-VIR - an AI algorithm that predicts severe adverse events and molecular features for COVID-19’s complications"

Jeffrey Skolnick

**This PDF file includes:**

Supplementary text

Figures S1 to S4

Tables S0 to S7

SI References

**Supplementary Information Text**

**Supplementary Materials and Methods**

An overview of the **MOATAI-VIR** approach was shown in main text Figure 1, with a detailed flowchart shown in Figure S1. The goal of **MOATAI-VIR** is to identify severe adverse responses associated with SARS-CoV-2 and their corresponding human mode of action (MOA) proteins. To accomplish these objectives, we input either the experimentally determined human proteins from the human-SARS-CoV-2 interactome(1) or the COVID-19 GWAS survival associated risk genes(2, 3) as a set of MOA proteins in a SARS-CoV-2 MOA profile. We then employ our recently developed **LeMeDISCO** algorithm which compares these SARS-CoV-2 MOA proteins to the **MEDICASCY**(4) predicted MOAs of 3,608 diverse disease indications. **LeMeDISCO** calculates a Z-score that determines diseases significantly associated with either the human component of the SARS-CoV-2-human interactome or the GWAS-based SARS-CoV-2 MOA protein profiles(1-3). **LeMeDISCO** then outputs indications that likely cause the adverse events resulting from SARS-CoV-2 infection.

**Application of MEDICASCY for the large-scale prediction of MOA proteins for diseases**

While **MEDICASCY** has been published(4), a brief description is provided for the convenience of the reader. The flowchart in Figure S2 shows how indications are predicted by **MEDICASCY**. **MEDICASCY** requires the structures of all proteins in the human exome. These were modeled by a fast version of the **TASSER^VMT^** algorithm(5-7) applied to the entire human exome (<ftp://ftp.ncbi.nih.gov/genomes/H_sapiens/protein/)>. Then, two classes of features were computed for the machine learning of indications from the input drugs: (a) A 256 dimensional molecular fingerprint computed directly from the chemical structure that is converted to its MACCS fingerprint(8) using the Open Babel software (<http://openbabel.org/wiki/Main_Page)>. (b) The other feature is generated from a drug’s predicted human protein targets using the latest version of **FINDSITE^comb2.0^**’s(9) virtual ligand screening of the given drug against the 32,584 (97%) human proteins with pre-computed structures and ligand binding pockets(7). Protein targets are then filtered for possible disease associations by requiring that at least >5% of all possible missense or frameshift/stop mutations of a given protein are disease associated as assessed by **ENTPRISE/ENTPRISE-X**(10, 11). The idea is that if essentially all variations of the protein are neutral, then it is highly unlikely that it is a mode of action protein for any disease. **ENTPRISE/ENTPRISE-X** both have low false positive rates, which is essential considering the very large number of variations found in the human exome.

Having predicted that a given protein is associated with some disease, the next issue is to identify which disease. To accomplish this, each protein is mapped to a 960 dimensional disease profile that serves as input to **Know-GENE**(12) whose output predicts the diseases a protein could be associated with. In principle, these two steps could provide the mapping of proteins to indications. However, this mapping is limited to proteins interacting with those proteins having some (but not all) disease associations that are used for gene-disease association inference.

Next, we employ a more powerful approach that uses drugs as a probe to identify proteins associated with a given disease. To accomplish this objective, a newly developed, Boosted Random Forest (BRF) machine learning approach for multiple label regression is employed for learning and prediction(4). BRFs build on the Random Forest (RF) approach which is an ensemble learning method for classification and regression(13). However, the BRF gives identical results to the RF but is much faster with much smaller memory requirements. The BRF learns indications of a drug by its fingerprint feature from drugs of similar structure and by the disease profile feature from drugs of similar disease profiles derived from their human protein targets. These two features are independently used in both training and prediction. The final prediction score is the average of these two disparate prediction approaches.

In practice, **MEDICASCY** covers 3,608 indications defined in the Human Disease Ontology database(14). Those indications are collected from three different sources: (1) the approved drug subset from the Therapeutic Target Database (TTD version Sept 12, 2017)(15), (2) the SIDER4 indication set(16), and (3) all the clinical trial drug sets were collected from ClinicalTrials.gov and mapped to DrugBank(17) by Himmelstein *et al*(18). All disease indications are converted to Human Disease Ontology IDs(14) (releases/2018-12-17) and merged if the Tanimoto Coefficients (T_c_)(19) of the two drugs are one. The merged dataset has 2,059 drugs with 3,608 unique indication terms and 123,146 drug-indication pairs. These indications cover diverse disease classes.

**MEDICASCY** can also infer possible MOA proteins for a given indication. Figure S3 shows the flowchart for inferring MOA proteins for a given disease. **MEDICASCY** starts by mapping the probe drugs to their respective indications. Once the drug-protein mapping (as predicted by **FINDSITE^comb2.0^** VLS(9)) is known, we can infer protein-indication mapping. The goal is to infer the putative MOAs of a disease by calculating the enrichment of a protein target T for a given disease D. To achieve this, we use a set of drugs with possible diverse indications as probes to map their indications, and then, by combining this information with **FINDSITE^comb2.0^**’s predicted drug-protein mapping, we infer protein-indication relationships. Here, we choose the 2,095 FDA-approved drugs from the DrugBank (version 5.09) database(17) as the probe set.

**MEDICASCY** is applied in prediction mode (i.e., any training drugs having a Tanimoto-Coefficient =1 to a given input drug are excluded from training) to avoid a strong bias towards drugs in the training set. For each of the 3,608 indications, we rank the 2,095 probe drugs according to their Z-scores, Z_d_, defined using the raw score computed by **MEDICASCY** from:

$Z_{d}=(\frac{raw score - average raw score of 2,095 drugs}{standard deviation of 2095 raw scores} )$	(1)

To predict a drug as having the given indication, we applied a Z_d_ cutoff of 2 that approximately corresponds to a p-value of 0.02 for the upper tailed null hypotheses. Thus, for each indication D, the 2,095 probe drugs are separated into two groups: N1 are predicted to have indication D (Z_d_$\geq$2) and N2 (=2,095-N1) are not predicted to have indication D (Z_d_<2). We should point out that this is a very loose prediction of a drug’s indication. The advantage of using a Z_d_ cutoff for the prediction is that for a given indication, it always predicts some drugs having the indication with its expected statistical confidence.

We then examine each protein target, T, in the human proteome of our modeled 32,584 proteins. There are a subset of the drugs (or perhaps none) predicted by **FINDSITE^comb2.0^**(9) to bind T. For a given indication D and a human protein target T, we define the relative risk RR(D,T) of the given target T with respect to indication D as:

RR(D,T)=$\frac{fraction of drugs with indication D binding to the target T}{fraction of drugs without indication D binding to the target T}$ `	(2)

The numerator is the estimation of the probability of drugs having the predicted indication D (Z_d_$\geq$2) that bind to protein T and is calculated as F1=(# of drugs having Z_d_$\geq$2 and binding to T)/N1. The denominator is the probability of finding drugs that do not have the predicted indication D but which bind to protein T calculated as F2=(# of drugs having Z _d_$<$2 and binding to T)/(2,095-N1). This latter probability serves as the background probability that an arbitrary drug will bind to T. When no drug is predicted to bind to protein T, RR(D,T) is set to zero. RR(D,T)=F1/F2 > 1 means that a drug having indication D is more likely to bind to T than arbitrary drugs not having the predicted indication D will bind to T.

We then compute the statistical significance of RR(D,T) by calculating a p-value using the method described in (20). We define a protein target T as predicted to be a possible MOA target for indication D if its p-value<0.001 because it is more likely to be targeted by its efficacious drugs than an arbitrary drug. In the end, for each of the 3,608 indications, there is a list of predicted possible MOA proteins.

**Comorbidity predictions by LeMeDISCO**

A flowchart of the **LeMeDISCO** comorbidity prediction algorithm is shown in Figure S4. Using the input of two sets of putative MOA proteins from two diseases predicted by **MEDICASCY** or obtained from experimental data, one then calculates the Comorbidity Factor CF(D_1_,D_2_) of disease D_2_ to D_1_ defined below in eq. 3. We then calculate a Z-score, Z_Co_, based on the overlapped MOA proteins of the two diseases to predict the significance of CF(D_1_,D_2_) by

CF(D_1_,D_2_)= N_s_ /(N_D2_ ×(N_D1_/$N_{t}$))	(3a)

Z_Co_ =$(N_{s}-{(N}_{D2}\times\frac{N_{D1}}{N_{t}}))/(\frac{\sqrt{N_{D1}\times N_{D2}\times\left( N_{t}-N_{D1} \right)}}{N_{t}})$	(3b)

$N_{D1}$, $N_{D2}$ are the numbers of MOA proteins/genes of disease D_1_, D_2_$; N_{s}$ is the number of overlapped MOAs between D_1_, D_2_ and $N_{t}$ is the total human proteins/genes. Thus, CF is the ratio of the observed number of MOA protein overlaps over the expected random number of MOA proteins. Since both $N_{D1}$ and $N_{D2}$ < 0.1$\times$N_t_ for the majority of the indications, a normal distribution of the null hypothesis for $N_{s}$ holds(20). Eq. 3b is based on the normal distribution approximation of $N_{s}$ when generated by the null hypothesis (or random)(20). Again, Z_Co_ > 2 corresponds to a p-value of ~ 0.02 for the upper tailed null hypothesis. Thus, the higher Z_Co_, the more significant is the shared number of MOA proteins. We will use Z_Co_ for predicting comorbidity and compare it with observed comorbidity.

Common probable MOA targets between disease pairs can be derived from either experimental data or **MEDICASCY** predictions. The types of experimental data that can be used include, but are not limited to, differential gene expression (GE), Mendelian or somatic mutation profiles comparing disease vs. control normal samples, better vs. worse prognosis samples, or drug treated vs. control untreated samples(21). Similarly, the types of input MOA proteins for **LeMeDISCO** are flexible. Thus, **LeMeDISCO** has a unique advantage of being able to test diverse hypotheses relevant to drug repurposing and discovery. It also can prioritize probable MOA proteins for subsequent experimental target validation and has application to precision medicine or targeted therapies. For COVID-19, we use its human interacting proteins(1) and GWAS risk genes(2, 3) as input MOA protein sets.

**Validating the use of Z_Co_ for comorbidity prediction**

First, we validated **LeMeDISCO**’s Z_Co_ in eq. 3 by correlating it with the observed comorbidity as quantified by these two measures: (a) the logarithm of relative risk log(RR) score and (b) the φ**-**score (Pearson's correlation for binary variables)(22). The relative risk (RR) score is the probability that two diseases occur in a single individual relative to random occurrence, and scales exponentially with respect to the strength of two diseases interacting or influencing each other. Thus, instead of using the RR score, as done in previous studies(22-25), here we use the log(RR) score for correlation analysis. The log(RR) and φ**-**score are computed from US Medicare insurance claim data of approximately 13,038,014 individuals, who had the 32,341,347 inpatient hospital visits(22) using the following equations:

Log(RR) = log($\frac{n_{AB}/n_{tot}}{{(n}_{A}/n_{tot}){(n}_{B}/n_{tot})})$	(4a)

$\varphi\mathbf{-}score=(n_{AB}*n_{tot}-n_{A}*n_{B})/\sqrt{n_{A}*n_{B}*\left( n_{tot}-n_{A} \right)*\left( n_{tot}-n_{B} \right)}$	(4b)

where n_tot_ = total number of patients in the data set; n_A_, n_B_ = number of patients diagnosed with disease A and B, respectively; n_AB_ = number of patients diagnosed with both diseases A and B.

We also compared Z_Co_ to other scores such as (A) the XD score from known disease-gene associations and protein-protein network propagation(23), (B) NG, is the number of shared genes between disease pairs(23), (C) the *S_AB_* score, a protein-protein network-based separation of a disease pair calculated from known disease-gene associations, defined as S_AB_=<d_AB_>-(<d_AA_>+<d_BB_>)/2, where S_AB_ compares the shortest distances between proteins within each disease A & B(25), <d_AA_> and <d_BB_>, to the shortest distances <d_AB_> between A-B protein pairs(25), and (D) the symptom similarity score obtained from text mining(24).

In addition to the correlation analysis, we also calculated the recall rate of each prediction score. This is important in that conclusions drawn from a small fraction of true comorbid diseases might not be true in general. We define a positive comorbidity pair when their log(RR) > 0 or RR>1 and a comorbidity prediction by Z_Co_ > 1.65 (corresponding p-value<0.05), XD score > 0, *S_AB_* score < 0, or the symptom similarity score > 0.1(19). The recall rate is defined as

$recall=\frac{Number of correctly predicted comorbidity pairs}{Total number of true comorbidity pairs}$	(5)

Table S0 summarizes the results of testing **LeMeDISCO**’s Z_Co_ and the comparison to the XD score(23), NG(23), the *S_AB_* score(25) and the symptom similarity(24) for correlations with comorbidity quantified by the log(RR) score and φ**-**score. Note that Z_Co_, XD score, NG and symptom similarity are expected to have positive correlations and the *S_AB_* score (a distance measure on interactome between two diseases) is expected to have negative correlations with the log(RR) score and φ**-**score. If the results have opposite correlations, no p-values are provided. Here, we consider the symptom similarity score of ref.(24) as a score for predicting comorbidity rather than a measure of likely comorbidity since it is not computed directly from the observed disease frequencies.

Mapping the DOIDs from the Human Disease Ontology database to the ICD-9 IDs of Ref.(22), we obtain 198,149 disease pairs for use in large scale testing of **LeMeDISCO**. To compare **LeMeDISCO**’s results using Z_Co_ with the XD score, which is the closest to our shared MOA protein approach, we mapped their ICD-9 disease code to the DOIDs and obtained a subset of 29,783 pairs from their dataset of 97,665 pairs(23). For comparison to *S_AB_* score(25), the MeSH(26) disease names(27) were mapped to DOIDs for consistency. A consensus set of 947 disease pairs from their dataset and our dataset of 198,149 was obtained. A similar dataset of 2,630 disease pairs was obtained for comparison with the symptom similarity score (consensus of Supplementary dataset 4 of (24) with the above set of 198,149 pairs). In addition to calculating the Pearson’s correlation of all data points, we also divided the data points into 10 bins according to the respective score range of Z_Co_, XD, *S_AB_* and symptom similarity score. The corresponding log(RR) and φ**-**scores are averaged in each bin. For example, if *S_AB_* is in the range *S_AB_min_* to *S_AB_max_*, a bin size of (*S_AB_max_* - *S_AB_min_*)/10 is used to partition the data points (disease pairs) into 10 bins according to each datum point’s *S_AB_* value. Inside each bin, the log(RR) or φ**-**scores are averaged over the data points; then, the center value of the bin represents the prediction score *S_AB_*. This gives equal weight to the rare prediction scores in the correlation analysis.

For the large set of 198,149 disease pairs, all correlations of Z_Co_ with log(RR) score and φ**-**score are statistically significant (p-value < 0.05). We also tested the correlations of the number of shared MOA proteins (NP) between two diseases by **LeMeDISCO**. They are all worse than **LeMeDISCO**’s Z_Co_ and have an insignificant (p-value > 0.05) when data binning is applied. For the 29,783 disease pairs used for the XD score comparison, **LeMeDISCO**’s Z_Co_ is obviously better than the XD score. Their NG score essentially has no significant correlation with log(RR) and only shows correlation with the φ**-**score for unbinned data. When the data are binned, both the XD score and NG score have no significant correlations. This demonstrates that the shared number of genes derived from known disease-gene associations is not good enough for interpreting comorbidity.

Compared to the *S_AB_* score(25) on the 947 disease pairs, **LeMeDISCO**’s Z_Co_ is slightly worse than the *S_AB_* score for unbinned data but better than the *S_AB_* score for binned data. The reason for this difference is that the correlation for unbinned data is dominated by those data points where the prediction scores are concentrated. For *S_AB_* scores, 874/947 are > 0, and thus, *S_AB_* score’s correlation for unbinned data is dominated by those points with *S_AB_* score > 0. In this region, no comorbidity is predicted, whereas for binned data, the rare predictions with *S_AB_* score < 0 (where comorbidity is predicted) will have equal effects as those whose *S_AB_* score > 0 on the correlation. We note that the sparse predictions of *S_AB_* score < 0 means that its recall rate is very low if a *S_AB_* score < 0 is considered to be a positive comorbidity prediction.

The symptom similarity score has better correlations than **LeMeDISCO**. However, it only explains the relationship of one phenotype (symptom) to another phenotype (disease) and lacks the ability to identify the underlying molecular mechanism responsible for this comorbidity. The recall for the symptom similarity score is 100% because it already discards non-significant scores (< 0.1). Nevertheless, all correlations of Z_Co_ are statistically significant, and its recall rate is close to 70% for these 2,630 disease pairs. The advantage of Z_Co_ over the symptom similarity score is that it has clear molecular interpretations as it provides the overlapping MOA proteins. Moreover, **LeMeDISCO** does not rely on prior knowledge or symptomatic information for each disease. Hence, it provides a much larger coverage of comorbidity predictions of the 198,149 disease pairs each ranked by its Z_Co_ score to reflect the corresponding expected statistical confidence.

**Predicting COVID-19’s clinical manifestations**

We first predict the severe adverse consequences of COVID-19 using **LeMeDISCO**. **LeMeDISCO** predicts comorbid diseases by scanning the putative MOAs of input disease against those of the 3,608 library diseases. The input MOAs of COVID-19 are from either the experimentally determined human-SARS-CoV-2 interactome(1) or COVID-19 GWAS survival associated risk genes(2, 3). **LeMeDISCO** then calculates the Z_Co_ of the input proteins to the MOAs of the 3,608 library diseases taken from **MEDICASCY’s** predictions using eq. 3. Diseases are ranked by their respective Z_Co_ and are predicted as severe adverse consequences of COVID-19.

**Mapping the COVID-19 severe adverse events to clinical/uncharacterized manifestation groups**

The clinical manifestations from were first mapped to their corresponding ICD-10 code(s)(28). Then, the predicted COVID-19 severe adverse events were placed in their corresponding clinical manifestation group via overlapping ICD-10 code(s)(28). Some severe adverse events fell into multiple clinical manifestation groups as they may have more than one ICD-10 code(28). To further analyze the predicted comorbidities that did not map to a clinical manifestation group, the remaining diseases were grouped by their main ICD-10(28) classification for the subsequent analysis.

**CoPathway**

The objective of the **CoPathway** method is to establish lists of significant pathways associated with each clinical/uncharacterized manifestation. If there are n > 1 comorbidities in a clinical/uncharacterized manifestation group, then the frequency of indications for each MOA proteins were determined and hierarchically ranked. The top 100 MOA proteins for a given clinical manifestation was used for global pathway analysis. If there was only n = 1 comorbidity for a manifestation group, then the significant MOA proteins determined by MEDICASCY were used for the global pathway analysis. The Reactome(29) global pathway analysis tool was used to determine the most enriched and significant pathways associated with the inputted set of proteins for each manifestation group. We then extracted only the pathways with a p-value < 0.05 and further mapped the pathways to their top pathway classification for further insight and prospective.

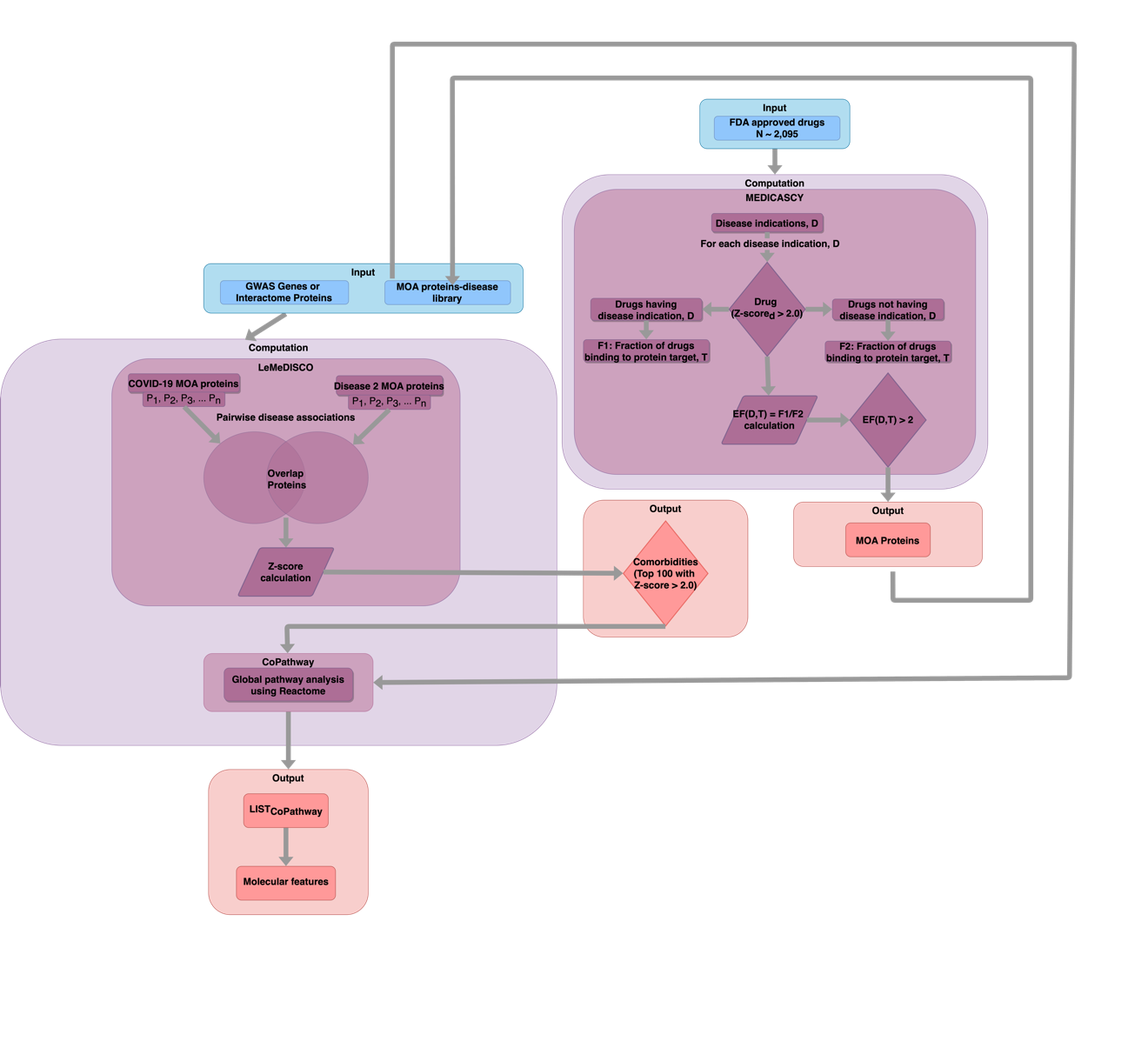
Fig. S1. Flowchart of MOATAI-VIR.

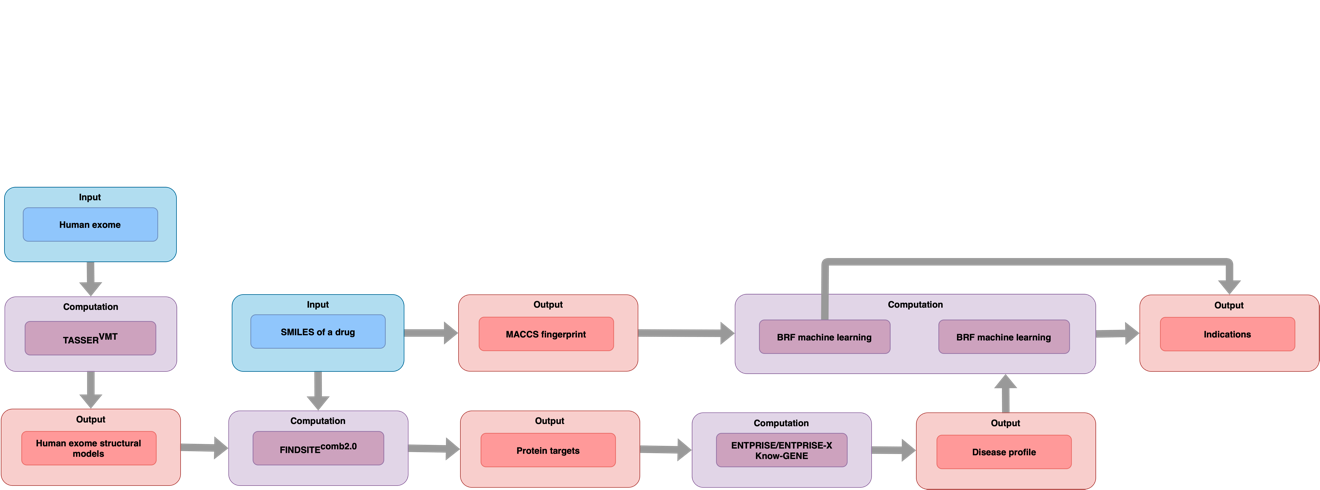

Fig. S2. Flowchart showing how indications are predicted by MEDICASCY.

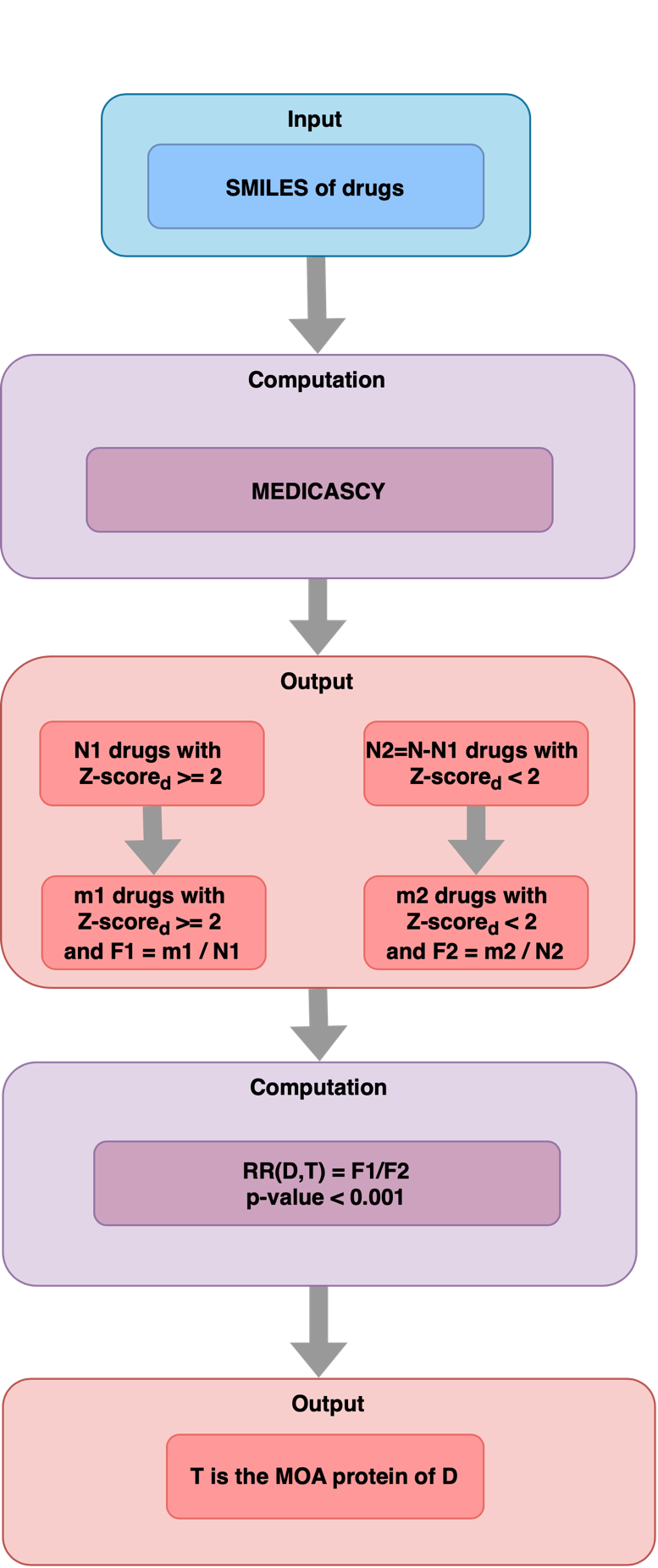

Fig. S3. Flowchart showing how MEDICASCY predicts the MOA proteins associated with a given disease.

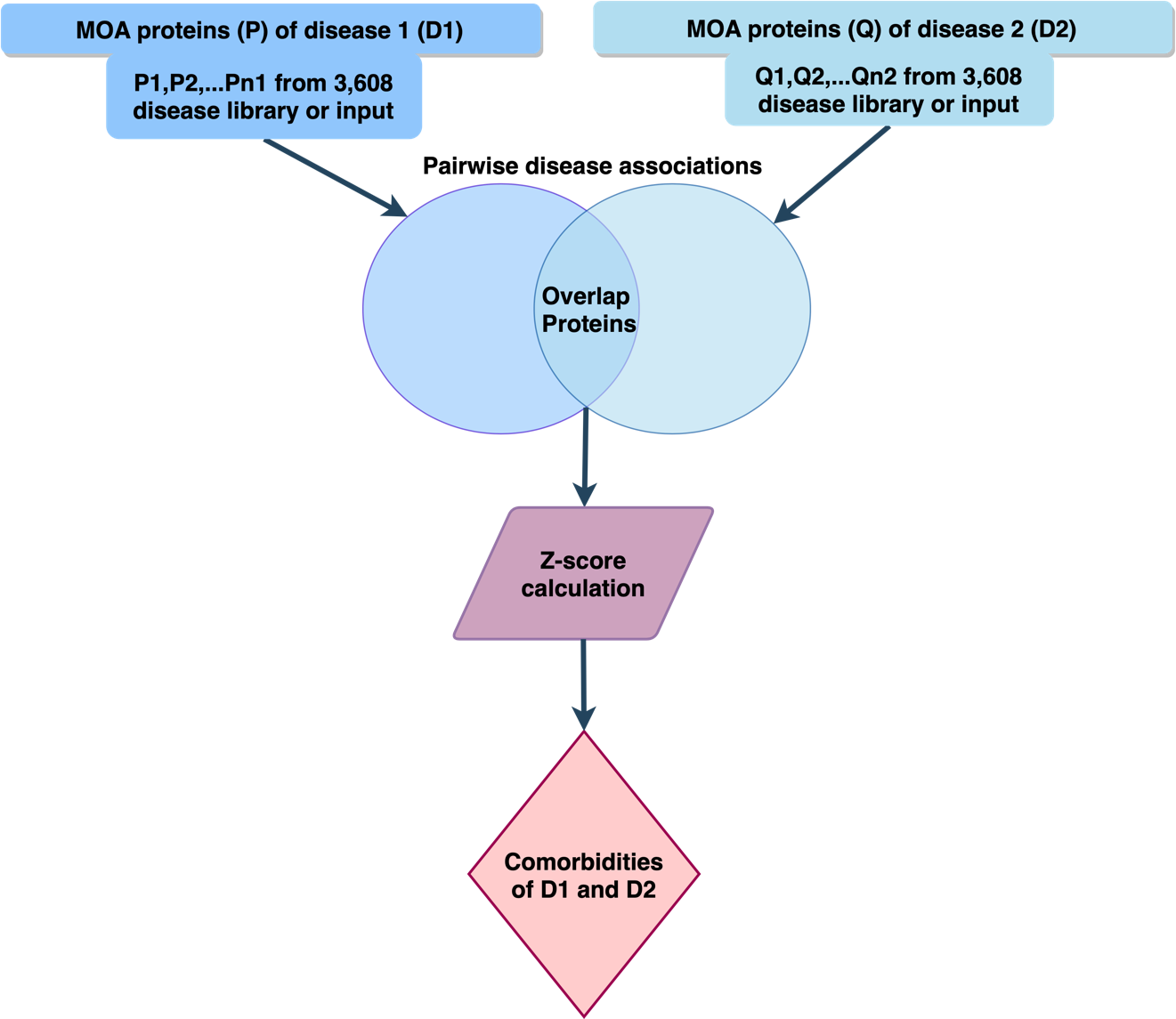

Fig. S4. LeMeDISCO Z-score (see eq. 3) based protocol for determining the comorbidity of a given pair of diseases.

**Table S0**. Comparison of **LeMeDISCO**’s Z_Co_ with the XD score, NG, *S_AB_* score and symptom

similarity for correlations with comorbidity quantified by the log(RR) score, φ**-**score and recall^a^.

|  | Unbinned^b^ | | 10 bins^b^ | | Recall |
| --- | --- | --- | --- | --- | --- |
|  | Log(RR) score | φ**-**score | Log(RR) score | φ**-**score |  |
| 198,149 pairs^c^ | | | | | |
| LeMeDISCO | **0.284(0.0)** | **0.194(0.0)** | **0.778(8.1E-3)** | **0.798(5.6E-3)** | **45.6%** |
| 29,783 pairs^d^ | | | | | |
| LeMeDISCO | **0.148(0.0)** | **0.106(0.0)** | **0.868(1.1E-3)** | **0.844(2.1E-3)** | **53.4%** |
| XD score(23) | 0.050(5.9E-18) | 0.082(0.0) | 0.445(0.20) | 0.252(0.48) | 6.5% |
| NG^e^ | 0.008(0.17) | 0.058(1.3E-23) | -0.436 | -0.175 | - |
| 947 pairs^f^ | | | | | |
| LeMeDISCO | 0.148(4.8E-6) | 0.172(1.0E-7) | **0.706(0.023)** | **0.670(0.034)** | **75.0%** |
| *S_AB_* score(25) | **-0.188(5.5E-9)** | **-0.218(1.2E-11)** | -0.671(0.034) | -0.473(0.17) | 8.5% |
| 2,630 pairs^g^ | | | | | |
| LeMeDISCO | 0.180(1.4E -20) | 0.172(6.5E-19) | 0.776(8.3E-3) | 0.663(0.037) | 67.8% |
| Symptom similarity(24) | **0.337(0.0)** | **0.197(1.6E-24)** | **0.950(2.6E-5)** | **0.960(1.1E-5)** | **100%** |

^a^ Numbers in parenthesis are the p-values of the corresponding correlation. Bold indicates the best results for the given data set.

^b^ Unbinned means raw data; each pair is a data point. 10 bins: partitioning the prediction scores into 10 equal size bins. In each bin, the Log(RR) & φ**-**score are averaged over data points in the bin. This gives equal weight to the rare prediction scores in the correlation analysis.

^c^ Mapping the DOID IDs from the human DO database to ICD9 IDs of Ref.(22), gives a set of 198,149 disease pairs

^d^ Mapped the ICD9 disease code to our DOID of DO and obtained a consensus subset of 29,783 pairs from Table S0 dataset of 97,665 pairs in Ref.(23).

^e^ NG is the number of shared genes between disease pairs in Ref.(23).

^f^ Consensus set of 947 disease pairs from the dataset of Ref.(25) and our dataset of 198,149.

^g^ A consensus dataset of 2,630 disease pairs was obtained from their Supplementary dataset 4 of Ref.(24) compared to our set of 198,149 pairs.

**Table S1.** Hierarchically ranked comorbidities, comorbidity enriched MOA proteins and pathways for each COVID-19 clinical manifestation from the SARS-CoV-2 interactome(1) as input results.

| **Clinical**  **manifestation** | **Comorbidities** | **Comorbidity**  **enriched**  **MOA proteins** | **Pathways**  **\| Top Pathways** |
| --- | --- | --- | --- |
| Respiratory | bronchial disease idiopathic pulmonary fibrosis pleural disease idiopathic interstitial pneumonia severe acute respiratory syndrome interstitial lung disease lymphangioleiomyomatosis pulmonary fibrosis Loeffler syndrome berylliosis viral pneumonia | KCNA10 OSBPL5 OSBPL8 ITGB1 ORC3 ELOVL1 C5orf46 SQRDL LXN CHDC2 ELOVL4 COX7A2 ITGB2 CCDC34 PRELID1 COX7A1 ITGB3 ELOVL7 PPP2R1B ITGB6 PPP2R1A RNF139 NR4A3 PHTF2 ANO6 ARFGEF2 ITGB5 PIGQ SF3B1 PITPNM2 ELOVL3 TOMM5 TMEM86B BTAF1 PITPNA NR0B1 NUP107 ICMT GRP AR TCP11 APH1A TSPAN13 OR9A2 FAM160A1 ITGB7 CHCHD7 COX7A2L TMEM185A SLC8A3 PGR PITPNB DGKE PITPNC1 ARFGEF1 PITPNM1 NR3C2 FASLG IL22RA2 TNFSF13 PRKCD DCAF8L2 CSF2RA FKBP11 PMP22 C1QL3 PSD FKBP14 GBF1 TNFSF12-TNFSF13 SCIN IL2RA FKBP9 FKBP2 TNFSF18 CD70 LTA TNF FKBP5 FKBP1B TNFSF14 OR1S2 CYB561A3 LTB SLC8A1 C2orf48 CCT6B FKBP1A PSD2 NR3C1 VPS41 PSD4 IARS2 FKBP10 FKBP7 EMP3 SLC8A2 FKBP4 WDR33 MKL1 | Nuclear Receptor transcription pathway \| Gene expression (Transcription) TNFs bind their physiological receptors \| Immune system Molecules associated with elastic fibres \| Extracellular matrix organization HSP90 chaperone cycle for steroid hormone receptors (SHR) \| Cellular responses to external stimuli TNFR2 non-canonical NF-kB pathway \| Immune system TNF receptor superfamily (TNFSF) members mediating non-canonical NF-kB pathway \| Immune system Elastic fibre formation \| Extracellular matrix organization Reduction of cytosolic Ca++ levels \| Hemostasis Sodium/Calcium exchangers \| Transport of small molecules PP2A-mediated dephosphorylation of key metabolic factors \| Metabolism Synthesis of very long-chain fatty acyl-CoAs \| Metabolism E2F mediated regulation of DNA replication \| Cell cycle RHO GTPases Activate Formins \| Signal transduction Association of TriC/CCT with target proteins during biosynthesis \| Metabolism of proteins Signaling by GSK3beta mutants \| Disease ERKs are inactivated \| Signal transduction MASTL Facilitates Mitotic Progression \| Cell cycle Signaling by CTNNB1 phospho-site mutants \| Disease S33 mutants of beta-catenin aren't phosphorylated \| Disease T41 mutants of beta-catenin aren't phosphorylated \| Disease S45 mutants of beta-catenin aren't phosphorylated \| Disease S37 mutants of beta-catenin aren't phosphorylated \| Disease Synthesis of PI \| Metabolism Interleukin-4 and Interleukin-13 signaling \| Immune system Platelet calcium homeostasis \| Hemostasis "Regulation of glycolysis by fructose 2 \| 6-bisphosphate metabolism" Beta-catenin phosphorylation cascade \| Signal transduction Fatty acyl-CoA biosynthesis \| Metabolism ECM proteoglycans \| Extracellular matrix organization Inhibition of replication initiation of damaged DNA by RB1/E2F1 \| Cell cycle Interleukin receptor SHC signaling \| Immune system Acyl chain remodeling of PS \| Metabolism TGFBR1 LBD Mutants in Cancer \| Disease PI and PC transport between ER and Golgi membranes \| Metabolism Adrenaline signaling through Alpha-2 adrenergic receptor \| Hemostasis Ion homeostasis \| Muscle contraction Linoleic acid (LA) metabolism \| Metabolism |
| Cardiovascular/Arrhythmia | coronary thrombosis vascular disease atherosclerosis carotid artery disease peripheral vascular disease brain stem infarction Wolff-Parkinson-White syndrome coronary restenosis coronary stenosis lymphatic system disease pulmonary embolism and infarction intermediate coronary syndrome atrioventricular block | DDRGK1 RPS19 CCT6B FKBP10 FKBP1A FKBP1B FKBP2 FKBP7 FKBP3 IL7R FKBP11 C2orf48 FKBP15 FKBP9 PACRGL FKBP14 AWAT1 GSN CDC20B WDR13 TCOF1 TLE3 EEF1D PPP3R2 TLE1 WDR33 IL21R BCL2L1 CDC20 DCAF8L2 IL4R CLSTN3 DCAF4L2 FKBP6 VPS41 EML4 CORO1C SCIN PAN2 IL2RA IL9R FKBP5 EIF2A MKL1 FKBP4 WDR89 PGLYRP4 IL2RB RPTOR TMEM81 PHTF2 AIP POLR3F IL2RG NAGK SQLE NOMO2 B4GALT4 METTL20 NAGPA RAB27A PXDNL TOMM5 PGLYRP3 TPO B4GALT2 CD248 PPARA AAGAB PGLYRP2 AGPAT9 EPX LPO PXDN RAB27B B4GALT3 MPO TOR4A ABCG1 SUV39H2 MRPS2 MKKS EXOC4 SSBP1 TMEM213 RAB19 DIP2C ABCF2 PRKAG2 GTPBP4 TUBAL3 SETD4 GIMAP4 GIMAP8 EZH2 CCDC3 HSPA14 ERG CASP2 ABCB8 | Keratan sulfate biosynthesis \| Metabolism N-Glycan antennae elongation \| Metabolism of proteins Keratan sulfate/keratin metabolism \| Metabolism Interleukin receptor SHC signaling \| Immune system Interleukin-21 signaling \| Immune system Events associated with phagocytolytic activity of PMN cells \| Immune system HSF1-dependent transactivation \| Cellular responses to external stimuli N-glycan antennae elongation in the medial/trans-Golgi \| Metabolism of proteins Glycosaminoglycan metabolism \| Metabolism RAB geranylgeranylation \| Metabolism of proteins TGFBR1 LBD Mutants in Cancer \| Disease Interleukin-2 signaling \| Immune system Defective Inhibition of DNA Recombination at Telomere Due to DAXX Mutations \| Disease Association of TriC/CCT with target proteins during biosynthesis \| Metabolism of proteins Attenuation phase \| Cellular responses to external stimuli The NLRP1 inflammasome \| Immune system Loss of Function of TGFBR1 in Cancer \| Disease |
| Acute myocardial infarction/  Unstable angina | coronary thrombosis intermediate coronary syndrome | FKBP4 GSN CILP TLE3 CDC20B PGLYRP1 DCAF8L2 LRRTM2 FKBP15 CD22 IL7R FKBP1A DDRGK1 IL2RB SGOL1 FGF14 FGF8 B4GALT4 EIF2A SI FOXA1 FGF12 FKBP3 WDR89 PACRGL AGPAT9 FGF2 IL2RA PGLYRP2 PPP3R2 SCIN NAGLU FKBP14 FKBP9 VPS41 MRPL32 FKBP10 FKBP6 PHTF2 NPTX2 AZGP1 FGF11 CCT6B LRRC17 FGF20 DCAF4L2 TLE1 IL21R MRPL4 PTX4 FAM8A1 NAGPA NOMO2 EEF1D FKBP5 IL4R MKL1 CLSTN2 WDR13 C2orf48 TARBP2 CD248 PRICKLE3 IL9R FKBP1B ZNF513 FKBP2 EML4 NRXN1 PAPPA2 B4GALT3 CRP APCS CLSTN3 FKBP11 CDC20 B4GALT2 PGLYRP3 PGLYRP4 RENBP FKBP7 WDR33 CORO1C PRKRA NAGK AWAT1 FGF13 IL2RG PITPNM2 PARP4 LRRC29 KRT7 FGF9 KRT75 WRAP53 ATMIN LTB4R COCH SLITRK1 PITPNA | Signaling by activated point mutants of FGFR1 \| Disease FGFR2c ligand binding and activation \| Signal transduction FGFR4 ligand binding and activation \| Signal transduction Signaling by activated point mutants of FGFR3 \| Disease FGFR1c ligand binding and activation \| Signal transduction FGFR3c ligand binding and activation \| Signal transduction FGFR3 ligand binding and activation \| Signal transduction FGFR1 ligand binding and activation \| Signal transduction FGFR2 ligand binding and activation \| Signal transduction FGFR3 mutant receptor activation \| Disease Activated point mutants of FGFR2 \| Disease FGFR3b ligand binding and activation \| Signal transduction Keratan sulfate biosynthesis \| Metabolism Phase 0 - rapid depolarisation \| Muscle contraction N-Glycan antennae elongation \| Metabolism of proteins Keratan sulfate/keratin metabolism \| Metabolism FGFR2 mutant receptor activation \| Disease SHC-mediated cascade:FGFR3 \| Signal transduction SHC-mediated cascade:FGFR4 \| Signal transduction SHC-mediated cascade:FGFR1 \| Signal transduction SHC-mediated cascade:FGFR2 \| Signal transduction Interleukin receptor SHC signaling \| Immune system Small interfering RNA (siRNA) biogenesis \| Gene expression (Transcription) Phospholipase C-mediated cascade; FGFR3 \| Signal transduction Phospholipase C-mediated cascade; FGFR4 \| Signal transduction Association of TriC/CCT with target proteins during biosynthesis \| Metabolism of proteins Phospholipase C-mediated cascade: FGFR1 \| Signal transduction Phospholipase C-mediated cascade; FGFR2 \| Signal transduction Interleukin-21 signaling \| Immune system FGFRL1 modulation of FGFR1 signaling \| Signal transduction N-glycan antennae elongation in the medial/trans-Golgi \| Metabolism of proteins Glycosaminoglycan metabolism \| Metabolism Signaling by Type 1 Insulin-like Growth Factor 1 Receptor (IGF1R) \| Signal transduction Digestion of dietary carbohydrate \| Digestion and absorption Synthesis of UDP-N-acetyl-glucosamine \| Metabolism of proteins Cardiac conduction \| Muscle contraction Antimicrobial peptides \| Immune system MicroRNA (miRNA) biogenesis \| Gene expression (Transcription) Interleukin-2 signaling \| Immune system PI3K Cascade \| Signal transduction MPS IIIB - Sanfilippo syndrome B \| Disease TGFBR1 LBD Mutants in Cancer \| Disease PI-3K cascade:FGFR3 \| Signal transduction PI-3K cascade:FGFR4 \| Signal transduction PI-3K cascade:FGFR1 \| Signal transduction PI-3K cascade:FGFR2 \| Signal transduction Constitutive Signaling by Aberrant PI3K in Cancer \| Disease Attenuation phase \| Cellular responses to external stimuli FRS-mediated FGFR3 signaling \| Signal transduction FRS-mediated FGFR4 signaling \| Signal transduction FRS-mediated FGFR1 signaling \| Signal transduction Negative regulation of FGFR3 signaling \| Signal transduction Negative regulation of FGFR4 signaling \| Signal transduction FRS-mediated FGFR2 signaling \| Signal transduction Protein-protein interactions at synapses \| Neuronal system IRS-mediated signalling \| Signal transduction Loss of Function of TGFBR1 in Cancer \| Disease Signaling by FGFR3 point mutants in cancer \| Disease Signaling by FGFR3 in disease \| Disease Negative regulation of FGFR2 signaling \| Signal transduction RUNX1 and FOXP3 control the development of regulatory T lymphocytes (Tregs) \| Gene expression (Transcription) Negative regulation of FGFR1 signaling \| Signal transduction FGFR1 mutant receptor activation \| Disease IRS-related events triggered by IGF1R \| Signal transduction Intestinal saccharidase deficiencies \| Disease Neurexins and neuroligins \| Neuronal system IGF1R signaling cascade \| Signal transduction |
| Hematologic | hemophagocytic lymphohistiocytosis paroxysmal nocturnal hemoglobinuria common variable immunodeficiency beta thalassemia neutropenia aplastic anemia pancytopenia purpura fulminans Diamond-Blackfan anemia alpha-2-plasmin inhibitor deficiency Fanconi anemia X-linked agammaglobulinemia thrombocytopenia factor VII deficiency factor V deficiency anemia protein S deficiency hemoglobin C disease hemoglobinopathy Chediak-Higashi syndrome obsolete leukemoid reaction chronic granulomatous disease thymoma histiocytosis mastocytosis protein C deficiency bone marrow disease myelofibrosis MHC class II deficiency hemolytic anemia congenital hemolytic anemia primary immunodeficiency disease severe combined immunodeficiency autoimmune hemolytic anemia hematopoietic system disease polycythemia vasculitis autoimmune thrombocytopenic purpura Wiskott-Aldrich syndrome hypereosinophilic syndrome thalassemia | CCT6B GART HENMT1 METTL21B PAN2 METTL21A TFB1M METTL9 ATIC AIFM1 SEPT10 SQRDL MTHFS MTHFSD METTL17 VCPKMT ABCC9 APAF1 ARL17B SEPT5 AK4 MRAS SPAG16 EEF1A2 USB1 FAM153A NSUN2 NME9 SEPT6 TREX2 ATL1 PCMTD2 MSH3 METTL3 BCS1L MEPCE IARS GDPGP1 AMPD3 GUF1 EHD3 HRAS RBKS GTPBP8 TIMM8A GPN1 RRM1 TRMT2B PRKAR1B ERI3 NLRP10 METTL4 RNMT RAB22A TOP3B RAP1B ERI1 CNNM2 ORC1 ABCB6 NDUFAF7 RAB3B ADPRH RAB19 SRL SEPT12 VWA8 TMEM5 C9orf41 CMTM1 MTG1 AP1S3 CMPK1 IDNK NUBP1 RAB33A AMPD2 GMPR2 OPA1 EHD2 SEPT9 RAP2C RALB POLE2 ARL5A ABCB9 ERMN MTIF3 RUVBL1 NRAS RAB31 RUVBL2 IRGM RAB1A GTPBP10 TRMT44 HSPA6 RAP1A NPIPB3 HSPA5 | RAB geranylgeranylation \| Metabolism of proteins Signaling by RAF1 mutants \| Disease RAS GTPase cycle mutants \| Disease Signaling by high-kinase activity BRAF mutants \| Disease MAP2K and MAPK activation \| Signal transduction Oncogenic MAPK signaling \| Disease Signaling by RAS mutants \| Disease Signaling by moderate kinase activity BRAF mutants \| Disease Signaling downstream of RAS mutants \| Disease Paradoxical activation of RAF signaling by kinase inactive BRAF \| Disease Signaling by BRAF and RAF fusions \| Disease RAS signaling downstream of NF1 loss-of-function variants \| Disease Metabolism of nucleotides \| Metabolism of RNA Purine salvage \| Metabolism Neutrophil degranulation \| Immune system Signalling to ERKs \| Signal transduction p38MAPK events \| Signal transduction RAF activation \| Signal transduction Signalling to RAS \| Signal transduction Nucleotide salvage \| Metabolism Estrogen-stimulated signaling through PRKCZ \| Signal transduction "Defective ABCC9 causes CMD10 \| ATFB12 and Cantu syndrome" CREB1 phosphorylation through NMDA receptor-mediated activation of RAS signaling \| Neuronal system Inwardly rectifying K+ channels \| Neuronal system Defective ABCB6 causes MCOPCB7 \| Disease GRB2:SOS provides linkage to MAPK signaling for Integrins \| Hemostasis SOS-mediated signalling \| Signal transduction Activation of RAS in B cells \| Immune system Activated NTRK3 signals through RAS \| Signal transduction Activated NTRK2 signals through RAS \| Signal transduction ABC-family proteins mediated transport \| Transport of small molecules ATP sensitive Potassium channels \| Neuronal system SHC-related events triggered by IGF1R \| Signal transduction MET activates RAP1 and RAC1 \| Signal transduction Defective Mismatch Repair Associated With MSH2 \| Disease SMAC(DIABLO)-mediated dissociation of IAP:caspase complexes \| Programmed cell death GRB2 events in ERBB2 signaling \| Signal transduction Interconversion of nucleotide di- and triphosphates \| Metabolism |
| Pulmonary embolism | pulmonary embolism and infarction | LOC390956 GTF2H2C WDR13 OTC LANCL3 RBBP7 CTPS2 TBL1X GGT1 GGT5 GSTT1 GSTT2 PPIL2 GGT2 GNB1L IL17RA RAE1 CSTF1 GGT7 TASP1 SSC5D GRWD1 STRN4 PRODH2 FXYD3 WDR83 FZR1 SEH1L PYCR1 RPTOR LGALS3BP EXOC7 LLGL2 DCAF7 UTP18 RUNDC3A CCL8 CORO6 WSB1 GGT6 PAFAH1B1 WDR81 FBXO31 WDR59 KATNB1 CORO1A THOC6 WDR24 SV2B PPIB GNB5 GATM WDR76 ZNF106 WDR20 WDR25 GLRX5 ATXN3 GSTZ1 JDP2 ACYP1 SMOC1 DCAF5 L3HYPDH STRN3 ALOX5AP WSB2 PWP1 NUP37 NEDD1 CCDC184 MGST1 MFAP5 CD163L1 GNB3 CD3E FAM76B EED CORO1B ASRGL1 CD6 SLC43A1 DDB2 TALDO1 DPYSL4 PPP2R2D BUB3 GSTO2 GSTO1 BTRC SEC31B PPIF WDR5 PTGES WDR34 WDR31 CORO2A TLE4 DNAI1 SMU1 TBC1D31 PROSC NRG1 DPYSL2 PPP2R2A LOXL2 TNFRSF10A MSR1 GNB2 MBLAC1 DYNC1I1 SRCRB4D TBL2 PPIA MIOS WIPI2 NUP43 PPIL4 PEX7 CDC40 PPIL6 GSTA4 GSTA5 PPIL1 CLIC1 DDAH2 GPX5 PAK1IP1 GNB2L1 LTC4S THOC3 FBXW11 C5orf52 DPYSL3 PPP2R2B WDR55 C5orf63 PPIC GLRX MTX3 SV2C PPWD1 CWC27 ERCC8 GPX8 CDC20B WDR70 AGA PLRG1 MGST2 ENPEP SEC31A CXCL10 PPP2R2C CRMP1 GNB4 SLC25A20 NKTR SEC13 DNPEP SPAG16 WDR12 PPIL3 DYNC1I2 PPIG WDSUB1 CIAO1 PLGLB2 LOXL3 WDR92 ACYP2 STRN DPYSL5 LOC650157 TSSC1 PYCR2 WDR26 RBBP5 RNPEP GLRX2 RFWD2 DCAF6 MGST3 TMEM79 SV2A PPIAL4B PPIAL4D PPIAL4G WDR77 GSTM3 GSTM5 WDR47 DDAH1 DEPDC1 PARS2 GPX7 TXNDC12 PPIH CTPS1 PPIE CSF3R RBBP4 EIF3I CLIC4 PADI6 PADI3 PADI1 PADI2 FBXO44 GNB1 PTH2 OAZ2 SLC25A15 EEF1G DPYS GDAP1 SLC25A2 PPAT OAZ3 CLTCL1 MRPL42 CTSA FARS2 FARSB | Glutathione conjugation \| Metabolism Phase II - Conjugation of compounds \| Metabolism Aflatoxin activation and detoxification \| Metabolism Neddylation \| Metabolism of proteins Defective TPR may confer susceptibility towards thyroid papillary carcinoma (TPC) \| Disease Presynaptic function of Kainate receptors \| Neuronal system Nuclear Pore Complex (NPC) Disassembly \| Cell cycle Regulation of Glucokinase by Glucokinase Regulatory Protein \| Metabolism Glutathione synthesis and recycling \| Metabolism Activation of G protein gated Potassium channels \| Neuronal system G protein gated Potassium channels \| Neuronal system Inhibition of voltage gated Ca2+ channels via Gbeta/gamma subunits \| Neuronal system Inwardly rectifying K+ channels \| Neuronal system Activation of kainate receptors upon glutamate binding \| Neuronal system Vpr-mediated nuclear import of PICs \| Disease NEP/NS2 Interacts with the Cellular Export Machinery \| Disease Interactions of Vpr with host cellular proteins \| Disease Cooperation of PDCL (PhLP1) and TRiC/CCT in G-protein beta folding \| Metabolism of proteins Activation of GABAB receptors \| Neuronal system GABA B receptor activation \| Neuronal system tRNA processing in the nucleus \| Metabolism of RNA Viral Messenger RNA Synthesis \| Disease Glucagon-type ligand receptors \| Signal transduction NS1 Mediated Effects on Host Pathways \| Disease Prostacyclin signalling through prostacyclin receptor \| Hemostasis Metabolism of non-coding RNA \| Metabolism of RNA snRNP Assembly \| Metabolism of RNA Glycolysis \| Metabolism G-protein activation \| Signal transduction ADP signalling through P2Y purinoceptor 12 \| Hemostasis Toxicity of botulinum toxin type A (botA) \| Disease "Adrenaline \| noradrenaline inhibits insulin secretion" CRMPs in Sema3A signaling \| Developmental biology G beta:gamma signalling through BTK \| Signal transduction Thromboxane signalling through TP receptor \| Hemostasis Export of Viral Ribonucleoproteins from Nucleus \| Disease GABA receptor activation \| Neuronal system Amplification of signal from unattached kinetochores via a MAD2 inhibitory signal \| Cell cycle Amplification of signal from the kinetochores \| Cell cycle Transport of Ribonucleoproteins into the Host Nucleus \| Disease Transcriptional regulation by small RNAs \| Gene expression (Transcription) Separation of Sister Chromatids \| Cell cycle Biological oxidations \| Metabolism Amino acids regulate mTORC1 \| Cellular responses to external stimuli HCMV Early Events \| Disease Crosslinking of collagen fibrils \| Extracellular matrix organization Rev-mediated nuclear export of HIV RNA \| Disease EML4 and NUDC in mitotic spindle formation \| Cell cycle Resolution of Sister Chromatid Cohesion \| Cell cycle RHO GTPases Activate Formins \| Signal transduction ADP signalling through P2Y purinoceptor 1 \| Hemostasis Mitotic Spindle Checkpoint \| Cell cycle G beta:gamma signalling through PLC beta \| Signal transduction Nuclear Envelope Breakdown \| Cell cycle Class B/2 (Secretin family receptors) \| Signal transduction Synthesis of Leukotrienes (LT) and Eoxins (EX) \| Metabolism Toxicity of botulinum toxin type E (botE) \| Disease Urea cycle \| Metabolism Elastic fibre formation \| Extracellular matrix organization Glucagon-like Peptide-1 (GLP1) regulates insulin secretion \| Metabolism Vasopressin regulates renal water homeostasis via Aquaporins \| Transport of small molecules RNA Polymerase II Transcription Termination \| Gene expression (Transcription) LTC4-CYSLTR mediated IL4 production \| Disease Chaperonin-mediated protein folding \| Metabolism of proteins Toxicity of botulinum toxin type F (botF) \| Disease Toxicity of botulinum toxin type D (botD) \| Disease mRNA 3'-end processing \| Metabolism of RNA G alpha (z) signalling events \| Signal transduction Signal amplification \| Hemostasis Nuclear import of Rev protein \| Disease Gene Silencing by RNA \| Gene expression (Transcription) Proline catabolism \| Metabolism Mitotic Prometaphase \| Cell cycle Transport of Mature mRNA derived from an Intron-Containing Transcript \| Metabolism of RNA Transport of the SLBP independent Mature mRNA \| Metabolism of RNA Aquaporin-mediated transport \| Transport of small molecules Glucose metabolism \| Metabolism Protein folding \| Metabolism of proteins Thrombin signalling through proteinase activated receptors (PARs) \| Hemostasis PRC2 methylates histones and DNA \| Gene expression (Transcription) Transport of Mature mRNA Derived from an Intronless Transcript \| Metabolism of RNA Potassium Channels \| Neuronal system G beta:gamma signalling through CDC42 \| Signal transduction "TALDO1 deficiency: failed conversion of Fru(6)P \| E4P to SH7P "TALDO1 deficiency: failed conversion of SH7P \| GA3P to Fru(6)P Antigen processing: Ubiquitination & Proteasome degradation \| Immune system Glucagon signaling in metabolic regulation \| Metabolism PKMTs methylate histone lysines \| Chromatin organization SCF-beta-TrCP mediated degradation of Emi1 \| Cell cycle HCMV Infection \| Disease Interactions of Rev with host cellular proteins \| Disease Regulation of ornithine decarboxylase (ODC) \| Metabolism |
| Disseminated intravascular coagulation (DIC) | purpura fulminans | BCL2L1 RPS19 TCOF1 DDRGK1 CTSA CPVL PCDHGA11 MRPL55 BPY2B NCOA5 ABHD1 GGCX PCSK2 PCSK6 NR2F2 PCSK1 MTTP CAPN11 | BH3-only proteins associate with and inactivate anti-apoptotic BCL-2 members \| Programmed cell death Defective gamma-carboxylation of F9 \| Disease The NLRP1 inflammasome \| Immune system STAT5 activation downstream of FLT3 ITD mutants \| Disease Defective NEU1 causes sialidosis \| Disease Activation of NOXA and translocation to mitochondria \| Programmed cell death NGF processing \| Signal transduction Expression and Processing of Neurotrophins \| Signal transduction Activation of PUMA and translocation to mitochondria \| Programmed cell death Interleukin-38 signaling \| Immune system "Activation \| myristolyation of BID and translocation to mitochondria" Defective factor IX causes hemophilia B \| Disease Degradation of the extracellular matrix \| Extracellular matrix organization Gamma-carboxylation of protein precursors \| Metabolism of proteins Defects of contact activation system (CAS) and kallikrein/kinin system (KKS) \| Disease Diseases of hemostasis \| Disease "Activation \| translocation and oligomerization of BAX" Maturation of protein M \| Disease Peptide hormone biosynthesis \| Metabolism of proteins SMAD2/3 MH2 Domain Mutants in Cancer \| Disease DSCAM interactions \| Developmental biology Activation of BIM and translocation to mitochondria \| Programmed cell death Activation and oligomerization of BAK protein \| Programmed cell death Activation of BAD and translocation to mitochondria \| Programmed cell death Loss of Function of SMAD2/3 in Cancer \| Disease NTRK3 as a dependence receptor \| Signal transduction Insulin processing \| Metabolism of proteins Signaling by TGF-beta Receptor Complex in Cancer \| Disease Activation of BMF and translocation to mitochondria \| Programmed cell death "Gamma-carboxylation \| transport Virion Assembly and Release \| Disease "Synthesis \| secretion Diseases associated with glycosylation precursor biosynthesis \| Disease "Plasma lipoprotein assembly \| remodeling |
| Neurologic | familial hemiplegic migraine amyotrophic lateral sclerosis type 1 amyotrophic lateral sclerosis type 2 amyotrophic lateral sclerosis type 4 Balo concentric sclerosis brain compression streptococcal meningitis Mast syndrome oculopharyngeal muscular dystrophy distal myopathy photosensitive epilepsy visual epilepsy aseptic meningitis ulnar nerve lesion radial nerve lesion mitochondrial complex V (ATP synthase) deficiency nuclear type 2 Vagus nerve disease Bell's palsy plantar nerve lesion tarsal tunnel syndrome common peroneal nerve lesion lesion of sciatic nerve locked-in syndrome myoclonic cerebellar dyssynergia quadriplegia choreatic disease Miller Fisher syndrome SPOAN syndrome mitochondrial complex II deficiency Stiff-Person syndrome Grn-related frontotemporal lobar degeneration with Tdp43 inclusions secondary Parkinson disease post-vaccinal encephalitis hypoglossal nerve disease Cayman type cerebellar ataxia extrapyramidal and movement disease multiple cranial nerve palsy hyperekplexia hyperekplexia 1 facial paralysis hyperekplexia 2 hyperekplexia 3 alcoholic neuropathy polyneuropathy due to drug torsion dystonia 1 postencephalitic Parkinson disease congenital central hypoventilation syndrome obsolete Alpers syndrome glossopharyngeal neuralgia neuroleptic malignant syndrome Reye syndrome normal pressure hydrocephalus benign shuddering attacks Melkersson-Rosenthal syndrome hypomyelinating leukodystrophy 11 hypomyelinating leukodystrophy 5 hypomyelinating leukodystrophy 7 with or without oligodontia and-or hypogonadotropic hypogonadism hypomyelinating leukodystrophy 8 with or without oligodontia and-or hypogonadotropic hypogonadism syndromic X-linked intellectual disability type 10 relapsing-remitting multiple sclerosis chronic inflammatory demyelinating polyneuritis Landau-Kleffner syndrome thoracic outlet syndrome nemaline myopathy vascular myelopathy central nervous system lymphoma hereditary sensory and autonomic neuropathy type 7 temporal lobe epilepsy hereditary sensory and autonomic neuropathy type 6 hereditary sensory neuropathy type 1E glossopharyngeal nerve disease inclusion body myositis lateral medullary syndrome central core myopathy haemophilus meningitis toxic encephalopathy Leigh disease olfactory nerve disease agenesis of the corpus callosum with peripheral neuropathy Schwartz-Jampel syndrome 1 pantothenate kinase-associated neurodegeneration myoclonic dystonia 11 torsion dystonia 13 torsion dystonia 2 torsion dystonia 6 torsion dystonia 4 torsion dystonia 17 dystonia 5 neuromuscular disease dystonia 9 childhood onset GLUT1 deficiency syndrome 2 myotonic disease dystonia 21 paroxysmal nonkinesigenic dyskinesia 2 dystonia 16 paroxysmal nonkinesigenic dyskinesia 1 striatonigral degeneration dystonia 27 dystonia 23 juvenile myoclonic epilepsy dystonia 24 episodic kinesigenic dyskinesia 1 episodic kinesigenic dyskinesia 2 dystonia 25 dystonia 12 X-linked dystonia-parkinsonism torsion dystonia with onset in infancy blepharospasm cranial nerve disease median neuropathy Brown-Sequard syndrome myositis central pontine myelinolysis progressive bulbar palsy peroneal nerve paralysis chronic fatigue syndrome Huntington's disease-like 1 Huntington's disease-like 2 diplegia of upper limb neuropathy spinocerebellar ataxia type 1 with axonal neuropathy Alzheimer's disease 2 Alzheimer's disease 5 periodic limb movement disorder Alzheimer's disease 6 Alzheimer's disease 7 Alzheimer's disease 8 Alzheimer's disease 10 Alzheimer's disease 11 Alzheimer's disease 12 Alzheimer's disease 13 Alzheimer's disease 14 Alzheimer's disease 15 encephalitis Charcot-Marie-Tooth disease type 1A Charcot-Marie-Tooth disease type 1F Charcot-Marie-Tooth disease type 1D Charcot-Marie-Tooth disease type 1C Charcot-Marie-Tooth disease type 1B Charcot-Marie-Tooth disease type 1E Charcot-Marie-Tooth disease type 2A1 Charcot-Marie-Tooth disease type 2A2A Charcot-Marie-Tooth disease type 2B1 Charcot-Marie-Tooth disease type 2J Charcot-Marie-Tooth disease type 2I Charcot-Marie-Tooth disease type 2B Charcot-Marie-Tooth disease axonal type 2T Charcot-Marie-Tooth disease type 2R Charcot-Marie-Tooth disease axonal type 2F Charcot-Marie-Tooth disease type 2D Charcot-Marie-Tooth disease type 2E Charcot-Marie-Tooth disease axonal type 2H Charcot-Marie-Tooth disease axonal type 2K Charcot-Marie-Tooth disease type 2Y Charcot-Marie-Tooth disease axonal type 2P Charcot-Marie-Tooth disease axonal type 2Q Charcot-Marie-Tooth disease axonal type 2U Charcot-Marie-Tooth disease axonal type 2L Charcot-Marie-Tooth disease axonal type 2O Charcot-Marie-Tooth disease axonal type 2N Charcot-Marie-Tooth disease type 2B2 Charcot-Marie-Tooth disease axonal type 2C Charcot-Marie-Tooth disease type 4C Charcot-Marie-Tooth disease type 4J Charcot-Marie-Tooth disease type 4A Charcot-Marie-Tooth disease type 4D Charcot-Marie-Tooth disease type 4K Charcot-Marie-Tooth disease type 4B2 Charcot-Marie-Tooth disease type 4B1 Charcot-Marie-Tooth disease type 4H Charcot-Marie-Tooth disease type 4F Charcot-Marie-Tooth disease type 4B3 Charcot-Marie-Tooth disease type 4E Charcot-Marie-Tooth disease type 4G Charcot-Marie-Tooth disease dominant intermediate B Charcot-Marie-Tooth disease recessive intermediate C Charcot-Marie-Tooth disease dominant intermediate C Charcot-Marie-Tooth disease dominant intermediate D Charcot-Marie-Tooth disease dominant intermediate A Charcot-Marie-Tooth disease recessive intermediate D Charcot-Marie-Tooth disease recessive intermediate B Charcot-Marie-Tooth disease dominant intermediate E Charcot-Marie-Tooth disease dominant intermediate F Charcot-Marie-Tooth disease X-linked dominant 6 Charcot-Marie-Tooth disease X-linked recessive 2 Charcot-Marie-Tooth disease X-linked dominant 1 Charcot-Marie-Tooth disease X-linked recessive 5 Charcot-Marie-Tooth disease X-linked recessive 3 Charcot-Marie-Tooth disease X-linked recessive 4 autosomal dominant limb-girdle muscular dystrophy autosomal recessive limb-girdle muscular dystrophy autosomal recessive limb-girdle muscular dystrophy type 2A autosomal recessive limb-girdle muscular dystrophy type 2B autosomal recessive limb-girdle muscular dystrophy type 2C autosomal recessive limb-girdle muscular dystrophy type 2D autosomal recessive limb-girdle muscular dystrophy type 2E autosomal recessive limb-girdle muscular dystrophy type 2F autosomal recessive limb-girdle muscular dystrophy type 2G autosomal recessive limb-girdle muscular dystrophy type 2H autosomal recessive limb-girdle muscular dystrophy type 2J autosomal recessive limb-girdle muscular dystrophy type 2L autosomal recessive limb-girdle muscular dystrophy type 2Q obsolete autosomal recessive limb-girdle muscular dystrophy type 2R autosomal recessive limb-girdle muscular dystrophy type 2S autosomal recessive limb-girdle muscular dystrophy type 2Y autosomal recessive limb-girdle muscular dystrophy type 2O autosomal recessive limb-girdle muscular dystrophy type 2P autosomal recessive limb-girdle muscular dystrophy type 2T autosomal recessive limb-girdle muscular dystrophy type 2U autosomal recessive limb-girdle muscular dystrophy type 2M autosomal recessive limb-girdle muscular dystrophy type 2K autosomal recessive limb-girdle muscular dystrophy type 2N autosomal recessive limb-girdle muscular dystrophy type 2I obsolete autosomal dominant limb-girdle muscular dystrophy type 1A obsolete autosomal dominant limb-girdle muscular dystrophy type 1B obsolete autosomal dominant limb-girdle muscular dystrophy type 1C autosomal dominant limb-girdle muscular dystrophy type 1H autosomal dominant limb-girdle muscular dystrophy type 2 autosomal dominant limb-girdle muscular dystrophy type 1 autosomal dominant limb-girdle muscular dystrophy type 3 muscular dystrophy-dystroglycanopathy megaconial type congenital muscular dystrophy rigid spine muscular dystrophy 1 congenital muscular dystrophy 1B muscular dystrophy-dystroglycanopathy type B5 muscular dystrophy-dystroglycanopathy type B6 congenital muscular dystrophy due to integrin alpha-7 deficiency congenital muscular dystrophy due to LMNA mutation Aicardi-Goutieres syndrome alternating hemiplegia of childhood neurodegeneration with brain iron accumulation 2a neurodegeneration with brain iron accumulation 2b neurodegeneration with brain iron accumulation 4 neurodegeneration with brain iron accumulation 5 neurodegeneration with brain iron accumulation 6 hereditary spastic paraplegia 10 hereditary spastic paraplegia 11 hereditary spastic paraplegia 12 hereditary spastic paraplegia 13 hereditary spastic paraplegia 14 hereditary spastic paraplegia 15 hereditary spastic paraplegia 16 hereditary spastic paraplegia 17 hereditary spastic paraplegia 18 hereditary spastic paraplegia 19 hereditary spastic paraplegia 2 hereditary spastic paraplegia 23 hereditary spastic paraplegia 24 hereditary spastic paraplegia 25 hereditary spastic paraplegia 26 hereditary spastic paraplegia 27 hereditary spastic paraplegia 28 hereditary spastic paraplegia 29 hereditary spastic paraplegia 30 hereditary spastic paraplegia 31 hereditary spastic paraplegia 32 hereditary spastic paraplegia 34 hereditary spastic paraplegia 35 hereditary spastic paraplegia 36 hereditary spastic paraplegia 37 hereditary spastic paraplegia 38 hereditary spastic paraplegia 39 hereditary spastic paraplegia 3A hereditary spastic paraplegia 4 hereditary spastic paraplegia 41 hereditary spastic paraplegia 42 hereditary spastic paraplegia 43 hereditary spastic paraplegia 44 hereditary spastic paraplegia 45 hereditary spastic paraplegia 46 hereditary spastic paraplegia 48 hereditary spastic paraplegia 49 hereditary spastic paraplegia 53 hereditary spastic paraplegia 54 hereditary spastic paraplegia 55 hereditary spastic paraplegia 56 hereditary spastic paraplegia 57 hereditary spastic paraplegia 5A hereditary spastic paraplegia 6 hereditary spastic paraplegia 61 hereditary spastic paraplegia 62 hereditary spastic paraplegia 63 hereditary spastic paraplegia 64 hereditary spastic paraplegia 7 hereditary spastic paraplegia 72 hereditary spastic paraplegia 73 hereditary spastic paraplegia 75 hereditary spastic paraplegia 77 hereditary spastic paraplegia 8 hereditary spastic paraplegia 9A hereditary spastic paraplegia 9B myotonic dystrophy type 2 distal spinal muscular atrophy 1 Troyer syndrome chronic meningitis generalized epilepsy with febrile seizures plus Timothy syndrome | PARS2 MMP27 SV2B MMP28 MMP23B MMP25 MMP21 MMP19 SV2A MMP26 RNPEP CCL8 L3HYPDH C5orf52 CD3E MMP16 LGALS3BP PADI2 FBXO31 CD6 PLGLB2 FBXO44 C5orf63 CD163L1 SRCRB4D PADI6 PADI3 PADI1 CD5L CCDC184 SSC5D LOXL3 LOXL2 MSR1 CLIC1 DDAH2 HSBP1 GPX7 RPL35 DDAH1 GSTT2 GOSR2 MGST1 GLRX SLC43A1 TXNDC12 GSTO1 PTGES LEKR1 GGT5 FAM13C GSTO2 DMBT1 DEPDC1 FAM167B GSTT1 GGT6 CPLX3 SSNA1 CSF3R LTC4S SMOC1 GGT7 GSTM5 GSTM3 ACYP1 GGT2 JDP2 IFT81 GSTA5 SYCE3 MBLAC1 MGST2 ALOX5AP GPX5 LANCL3 GSTA4 GATM ACYP2 PPIL2 GLRX2 MGST3 IL17RA DTNA ATXN3 GPX8 TMEM79 TALDO1 DNPEP NRG1 CRMP1 FAM175A AARS KIAA1598 SCARA5 DPYSL2 DPYSL4 RASAL1 DPYSL5 HHIPL1 | Glutathione conjugation \| Metabolism Phase II - Conjugation of compounds \| Metabolism Aflatoxin activation and detoxification \| Metabolism Biological oxidations \| Metabolism Glutathione synthesis and recycling \| Metabolism CRMPs in Sema3A signaling \| Developmental biology Toxicity of botulinum toxin type E (botE) \| Disease Toxicity of botulinum toxin type A (botA) \| Disease Synthesis of Leukotrienes (LT) and Eoxins (EX) \| Metabolism Crosslinking of collagen fibrils \| Extracellular matrix organization GRB7 events in ERBB2 signaling \| Signal transduction Synthesis of Lipoxins (LX) \| Metabolism Synthesis of 5-eicosatetraenoic acids \| Metabolism Vitamin C (ascorbate) metabolism \| Metabolism "TALDO1 deficiency: failed conversion of SH7P \| GA3P to Fru(6)P "TALDO1 deficiency: failed conversion of Fru(6)P \| E4P to SH7P ERBB2 Regulates Cell Motility \| Signal transduction Toxicity of botulinum toxin type F (botF) \| Disease Toxicity of botulinum toxin type D (botD) \| Disease Arachidonic acid metabolism \| Metabolism ERBB2 Activates PTK6 Signaling \| Signal transduction Attenuation phase \| Cellular responses to external stimuli Scavenging by Class A Receptors \| Vesicle-mediated transport Semaphorin interactions \| Developmental biology Downregulation of ERBB2:ERBB3 signaling \| Signal transduction Essential pentosuria \| Disease HSF1-dependent transactivation \| Cellular responses to external stimuli Type II Na+/Pi cotransporters \| Transport of small molecules Elastic fibre formation \| Extracellular matrix organization |
| Cerebral ischemia/infarction | vascular disease atherosclerosis carotid artery disease peripheral vascular disease brain stem infarction lymphatic system disease | FKBP11 FKBP10 FKBP2 IL7R FKBP3 FKBP7 PACRGL FKBP15 FKBP14 FKBP1B FKBP1A FKBP9 C2orf48 RPS19 AWAT1 TOMM5 CCT6B PAN2 PHTF2 BCL2L1 FKBP4 NR5A2 IL21R FKBP6 CORO1C DCAF4L2 VPS41 CDC20 TMEM81 GSN NR5A1 TCOF1 WDR33 EML4 FKBP5 WDR89 AIP PIGQ PPP3R2 IL9R EEF1D CDC20B DCAF8L2 TLE3 OSBPL8 DDRGK1 CLSTN3 PPARA SCIN IL2RA WDR13 IL4R EIF2A RPTOR IARS2 GPR123 MKL1 TLE1 PGLYRP4 PKP3 METTL20 PPP1R21 LPO CARD11 TMEM185A ARFGEF1 CCHCR1 NAGPA SRBD1 PXDNL GPRC5A OR11A1 DGKE PGLYRP3 EPX MPO B4GALT3 PPP3R1 SPEG OR1K1 GUK1 NR0B1 OR1C1 OR14A16 OR11L1 CILP P2RY8 B4GALT4 TMEM215 PXDN TPO ORC3 OR1L4 EFR3A OR1N1 TCP11 PTGS1 PPARD PTGS2 RARRES1 | Nuclear Receptor transcription pathway \| Gene expression (Transcription) Keratan sulfate biosynthesis \| Metabolism Keratan sulfate/keratin metabolism \| Metabolism Events associated with phagocytolytic activity of PMN cells \| Immune system N-Glycan antennae elongation \| Metabolism of proteins HSF1-dependent transactivation \| Cellular responses to external stimuli Calcineurin activates NFAT \| Immune system Regulation of gene expression in early pancreatic precursor cells \| Developmental biology TGFBR1 LBD Mutants in Cancer \| Disease Association of TriC/CCT with target proteins during biosynthesis \| Metabolism of proteins Olfactory Signaling Pathway \| Signal transduction Interleukin-4 and Interleukin-13 signaling \| Immune system Attenuation phase \| Cellular responses to external stimuli The NLRP1 inflammasome \| Immune system Loss of Function of TGFBR1 in Cancer \| Disease |
| Endocrine | ovarian disease secondary hyperparathyroidism mucopolysaccharidosis III hypobetalipoproteinemia Wolman disease argininosuccinic aciduria calcinosis gangliosidosis bilirubin metabolic disorder glycogen storage disease II Chediak-Higashi syndrome lipid metabolism disorder lipomatosis lysosomal storage disease Tay-Sachs disease GM1 gangliosidosis Sandhoff disease insulinoma glucose metabolism disease thyroid gland disease pellagra diabetic retinopathy peroxisomal disease amyloidosis metachromatic leukodystrophy Krabbe disease | RDH13 RDH14 RDH12 RETSAT PLEKHG4 FAM84A HRASLS5 DHRS7 RLBP1 RDH8 RBP3 PLA2G16 FAM84B RDH11 OR1N1 HRASLS2 RARRES3 OR4X2 NR1I3 ELOVL7 KCNA10 OR11L1 COX7A1 COX7A2L RARRES1 KALRN OR2G6 ELOVL1 ELOVL4 COX7A2 SESTD1 AZI2 LRAT OR7D2 SAE1 MTPAP CCT6A AIFM1 FHIT TGDS CBWD1 ACTR8 CCT6B OR2T2 OSBPL8 CCT3 SLC8A3 IVD GALE OSBPL5 CHST5 LXN NUDT15 CDIPT PPIP5K1 ATL1 DDX5 FAM122C ATP5B DDX4 RBP4 NMRK1 OR1E2 IDNK GTPBP10 DHX58 AK6 TTL ARL17B LOC100294341 ADK DDX23 KIF11 WSCD1 RFC2 OR2G3 ATP5A1 ACSL4 NUDCD2 OR1C1 CBWD5 GNL2 MORC1 OSBPL9 NR4A3 NMNAT3 ADORA3 OR4C3 HINT3 PSMC4 DDX49 VDR DDX20 ERCC6 ZCCHC6 UXS1 KIF2A MAT1A SEPT6 UST | The canonical retinoid cycle in rods (twilight vision) \| Signal transduction Diseases associated with visual transduction \| Disease Diseases of the neuronal system \| Disease Retinoid cycle disease events \| Disease Olfactory Signaling Pathway \| Signal transduction Acyl chain remodelling of PE \| Metabolism Nuclear Receptor transcription pathway \| Gene expression (Transcription) Prefoldin mediated transfer of substrate to CCT/TriC \| Metabolism of proteins Visual phototransduction \| Signal transduction Fatty acyl-CoA biosynthesis \| Metabolism Synthesis of very long-chain fatty acyl-CoAs \| Metabolism Acyl chain remodelling of PS \| Metabolism RA biosynthesis pathway \| Signal transduction Association of TriC/CCT with target proteins during biosynthesis \| Metabolism of proteins Cooperation of PDCL (PhLP1) and TRiC/CCT in G-protein beta folding \| Metabolism of proteins Leading Strand Synthesis \| DNA replication Polymerase switching \| Cell cycle Folding of actin by CCT/TriC \| Metabolism of proteins Glycerophospholipid biosynthesis \| Metabolism Formation of tubulin folding intermediates by CCT/TriC \| Metabolism of proteins Gap-filling DNA repair synthesis and ligation in TC-NER \| DNA repair Translesion synthesis by POLI \| DNA repair Translesion synthesis by POLK \| DNA repair Formation of ATP by chemiosmotic coupling \| Metabolism Retinoid metabolism and transport \| Metabolism Gap-filling DNA repair synthesis and ligation in GG-NER \| DNA repair Retinoid metabolism disease events \| Disease G alpha (s) signalling events \| Signal transduction Signaling by Retinoic Acid \| Signal transduction Lagging Strand Synthesis \| DNA replication Dual incision in TC-NER \| DNA repair Defective GALE can cause Epimerase-deficiency galactosemia (EDG) \| Disease PCNA-Dependent Long Patch Base Excision Repair \| DNA repair Recognition of DNA damage by PCNA-containing replication complex \| DNA repair The retinoid cycle in cones (daylight vision) \| Signal transduction Adenosine P1 receptors \| Signal transduction Cooperation of Prefoldin and TriC/CCT in actin and tubulin folding \| Metabolism of proteins NTF3 activates NTRK3 signaling \| Signal transduction Defective MAT1A causes Methionine adenosyltransferase deficiency (MATD) \| Disease Translesion Synthesis by POLH \| DNA repair Resolution of AP sites via the multiple-nucleotide patch replacement pathway \| DNA repair NTF3 activates NTRK2 (TRKB) signaling \| Signal transduction Termination of translesion DNA synthesis \| DNA repair |
| Diabetic ketoacidosis/ Hyperglycemia and ketosis | diabetic retinopathy | GRP APH1B OSBPL8 OSBPL5 NR4A3 ANXA1 TSPAN13 NR3C1 SLC8A1 AR TMEM86B PRLR NR3C2 APH1A ITGB7 ITGB1 ITGB5 ITGB6 ITGB2 NUP107 KCNA10 ICMT ITGB3 PGR COX7A1 COX7A2 COX7A2L SLC8A2 SLC8A3 ELOVL3 ELOVL4 ELOVL7 ELOVL1 IL22RA2 AIFM1 CSF2RA SQRDL PPP2R1B PPP2R1A PSD2 FBXO8 FAM160A1 GBF1 BTAF1 ARFGEF1 ARFGEF2 PSD3 SF3B1 CHDC2 TRPC4 PSD UGGT1 PSD4 TMEM62 TCP11 PHTF2 TRPV1 TOMM5 IL21R ORC3 IL7R PRKCD IL9R IL4R PIGQ NR0B1 PPP3CC PPP6C LXN IARS2 IL2RB RARRES1 ERBB3 GPRC5A CYB561A3 THOC2 PLEKHG4 HRASLS2 HRASLS5 COX4I2 COX4I1 MRPL32 EMP3 AADAC RLBP1 PLA2G16 RARRES3 RBP3 FAM84A RDH8 FAM84B LRAT TMEM185A OSBPL9 VDR PACRGL PMP22 PTGR2 AWAT1 WDR13 DCAF8L2 MKL1 FKBP1A DGKE FKBP10 WDR89 FKBP3 CORO1C FKBP11 FKBP4 FKBP2 IL2RA GSN FKBP15 EEF1D DCAF4L2 FKBP9 FKBP14 SCIN FKBP5 CDC20B EIF2A FKBP7 WDR33 EML4 FKBP1B C2orf48 CDC20 TLE3 TLE1 FKBP6 VPS41 CYP11B1 TMED6 PPP3R2 ABCC8 BCMO1 PAN2 CDKN2B RPE65 RPTOR WRAP53 SLC39A8 TMEM81 AIP CLSTN3 CD248 OR9A2 EBLN2 RANBP1 OR4A16 WDR38 NR1I3 NOMO2 ARHGEF28 PGLYRP4 RASGRP2 PGLYRP3 OR5M11 B4GALT4 B4GALT3 B4GALT2 OR6K3 NAGLU GPR123 OSBPL3 DNAH12 NR5A1 NAGK SI UMODL1 THRA MC1R OSMR GPC3 OR10A7 OR13A1 CXCL2 CXCL3 CXCL5 PPBP PF4 CXCL1 PF4V1 CXCL6 CXCL8 OR52B4 GPR149 CCDC19 NR0B2 MAGEB1 CAPN3 PAH TPH2 NECAP1 TPH1 TH OR2F2 C20orf195 HOOK2 EPOR CEP112 DUOX1 GOLGA8R IKBIP C9orf117 SCAI OSR2 FGF20 CLIP2 ELMO1 TAX1BP1 IYD LRRFIP2 THRB TMEM177 ATRN PTH2 HAL YARS2 FANK1 AZGP1 GPR35 FARSB WARS2 YARS PPARA MRGPRX3 PAM JADE1 | Nuclear Receptor transcription pathway \| Gene expression (Transcription) Chemokine receptors bind chemokines \| Signal transduction N-Glycan antennae elongation \| Metabolism of proteins Keratan sulfate biosynthesis \| Metabolism Molecules associated with elastic fibres \| Extracellular matrix organization Metabolism of amine-derived hormones \| Metabolism PP2A-mediated dephosphorylation of key metabolic factors \| Metabolism Keratan sulfate/keratin metabolism \| Metabolism The canonical retinoid cycle in rods (twilight vision) \| Signal transduction Acyl chain remodelling of PE \| Metabolism Interleukin receptor SHC signaling \| Immune system Reduction of cytosolic Ca++ levels \| Hemostasis Sodium/Calcium exchangers \| Transport of small molecules RHO GTPases Activate Formins \| Signal transduction HSP90 chaperone cycle for steroid hormone receptors (SHR) \| Cellular responses to external stimuli Acyl chain remodelling of PS \| Metabolism Elastic fibre formation \| Extracellular matrix organization TRP channels \| Transport of small molecules Signaling by GSK3beta mutants \| Disease ERKs are inactivated \| Signal transduction MASTL Facilitates Mitotic Progression \| Cell cycle Signaling by CTNNB1 phospho-site mutants \| Disease S45 mutants of beta-catenin aren't phosphorylated \| Disease S37 mutants of beta-catenin aren't phosphorylated \| Disease S33 mutants of beta-catenin aren't phosphorylated \| Disease T41 mutants of beta-catenin aren't phosphorylated \| Disease NOTCH4 Activation and Transmission of Signal to the Nucleus \| Signal transduction Synthesis of very long-chain fatty acyl-CoAs \| Metabolism Defective F8 binding to von Willebrand factor \| Disease Association of TriC/CCT with target proteins during biosynthesis \| Metabolism of proteins "Regulation of glycolysis by fructose 2 \| 6-bisphosphate metabolism" Interleukin-4 and Interleukin-13 signaling \| Immune system Beta-catenin phosphorylation cascade \| Signal transduction E2F mediated regulation of DNA replication \| Cell cycle Effects of PIP2 hydrolysis \| Hemostasis |
| Gastrointestinal symptoms | esophagitis angular cheilitis oral hairy leukoplakia cheilitis gastroenteritis papilloma pericoronitis pulmonary coin lesion hemoglobinuria chronic fatigue syndrome stomatitis oral mucosa leukoplakia exanthem inflammatory bowel disease | WRAP53 OR4K5 DYNC1I2 WSB2 DYNC1I1 KATNB1 DCAF7 GNB2 GPR132 TSSC1 PPP2R2B RBBP4 UTP18 WDR55 PPP2R2D BUB3 SEC31A CSTF1 EIF3I CMKLR1 RFWD2 PWP1 STRN4 GRWD1 THOC3 PPP2R2C TBL2 TUBA4A DDB2 WDR76 ZNF106 RAE1 TBC1D31 WDR5 TOR1B WDR34 ERCC8 SMU1 WDR31 WDR13 DNAI1 TBL1X CDC20B WSB1 WDR47 TLE4 STRN3 WDR70 PLRG1 SEC13 GNB5 WDR20 WDR25 SEC31B TUBB1 WDR59 RRNAD1 CIAO1 DCAF5 MCM3AP WDR81 PPP2R2A PAFAH1B1 PARS2 GNB1L STRN RBBP7 NEDD1 CORO1B MIOS SPAG16 WIPI2 WDR24 PAK1IP1 NUP43 RPTOR RBBP5 WDR83 GNB3 CDC40 EED GNB1 PEX7 GNB4 THOC6 LLGL2 NUP37 GNB2L1 SEH1L WDR26 WDR12 NUDT5 CCR2 LIG4 EML4 SDCCAG3 SRPR RIMKLB DRG1 SYN3 | Mitotic Prometaphase \| Cell cycle EML4 and NUDC in mitotic spindle formation \| Cell cycle HCMV Early Events \| Disease Defective TPR may confer susceptibility towards thyroid papillary carcinoma (TPC) \| Disease Nuclear Pore Complex (NPC) Disassembly \| Cell cycle Regulation of Glucokinase by Glucokinase Regulatory Protein \| Metabolism Presynaptic function of Kainate receptors \| Neuronal system Resolution of Sister Chromatid Cohesion \| Cell cycle RHO GTPases Activate Formins \| Signal transduction Separation of Sister Chromatids \| Cell cycle G protein gated Potassium channels \| Neuronal system Inhibition of voltage gated Ca2+ channels via Gbeta/gamma subunits \| Neuronal system Activation of G protein gated Potassium channels \| Neuronal system Chaperonin-mediated protein folding \| Metabolism of proteins Vpr-mediated nuclear import of PICs \| Disease NEP/NS2 Interacts with the Cellular Export Machinery \| Disease HCMV Infection \| Disease Inwardly rectifying K+ channels \| Neuronal system Interactions of Vpr with host cellular proteins \| Disease Activation of kainate receptors upon glutamate binding \| Neuronal system tRNA processing in the nucleus \| Metabolism of RNA Protein folding \| Metabolism of proteins Viral Messenger RNA Synthesis \| Disease Amplification of signal from the kinetochores \| Cell cycle Amplification of signal from unattached kinetochores via a MAD2 inhibitory signal \| Cell cycle NS1 Mediated Effects on Host Pathways \| Disease Cooperation of PDCL (PhLP1) and TRiC/CCT in G-protein beta folding \| Metabolism of proteins Activation of GABAB receptors \| Neuronal system GABA B receptor activation \| Neuronal system Glucagon-type ligand receptors \| Signal transduction Prostacyclin signalling through prostacyclin receptor \| Hemostasis Export of Viral Ribonucleoproteins from Nucleus \| Disease Recruitment of NuMA to mitotic centrosomes \| Cell cycle M Phase \| Cell cycle Transport of Ribonucleoproteins into the Host Nucleus \| Disease Transcriptional regulation by small RNAs \| Gene expression (Transcription) G-protein activation \| Signal transduction ADP signalling through P2Y purinoceptor 12 \| Hemostasis Mitotic Spindle Checkpoint \| Cell cycle Neddylation \| Metabolism of proteins "Adrenaline \| noradrenaline inhibits insulin secretion" Amino acids regulate mTORC1 \| Cellular responses to external stimuli G beta:gamma signalling through BTK \| Signal transduction Thromboxane signalling through TP receptor \| Hemostasis GABA receptor activation \| Neuronal system Metabolism of non-coding RNA \| Metabolism of RNA snRNP Assembly \| Metabolism of RNA Rev-mediated nuclear export of HIV RNA \| Disease Transport of Mature mRNA derived from an Intron-Containing Transcript \| Metabolism of RNA Glycolysis \| Metabolism AURKA Activation by TPX2 \| Cell cycle Nuclear Envelope Breakdown \| Cell cycle ADP signalling through P2Y purinoceptor 1 \| Hemostasis G beta:gamma signalling through PLC beta \| Signal transduction Nuclear import of Rev protein \| Disease Gene Silencing by RNA \| Gene expression (Transcription) Mitotic Anaphase \| Cell cycle Glucagon-like Peptide-1 (GLP1) regulates insulin secretion \| Metabolism Vasopressin regulates renal water homeostasis via Aquaporins \| Transport of small molecules Transport of the SLBP independent Mature mRNA \| Metabolism of RNA G alpha (z) signalling events \| Signal transduction Signal amplification \| Hemostasis Transport of Mature mRNA Derived from an Intronless Transcript \| Metabolism of RNA Cellular response to heat stress \| Cellular responses to external stimuli RNA Polymerase II Transcription Termination \| Gene expression (Transcription) Aquaporin-mediated transport \| Transport of small molecules mRNA 3'-end processing \| Metabolism of RNA Thrombin signalling through proteinase activated receptors (PARs) \| Hemostasis Interactions of Rev with host cellular proteins \| Disease Mitotic Metaphase and Anaphase \| Cell cycle SUMOylation of SUMOylation proteins \| Metabolism of proteins COPI-independent Golgi-to-ER retrograde traffic \| Vesicle-mediated transport Potassium Channels \| Neuronal system G beta:gamma signalling through CDC42 \| Signal transduction Regulation of HSF1-mediated heat shock response \| Cellular responses to external stimuli Transport of the SLBP Dependant Mature mRNA \| Metabolism of RNA Class B/2 (Secretin family receptors) \| Signal transduction PKMTs methylate histone lysines \| Chromatin organization Transport of Mature mRNAs Derived from Intronless Transcripts \| Metabolism of RNA Recruitment of mitotic centrosome proteins and complexes \| Cell cycle Transport of Mature Transcript to Cytoplasm \| Metabolism of RNA Glucose metabolism \| Metabolism Loss of Nlp from mitotic centrosomes \| Cell cycle Loss of proteins required for interphase microtubule organization from the centrosome \| Cell cycle G alpha (12/13) signalling events \| Signal transduction Mitotic Prophase \| Cell cycle Glucagon signaling in metabolic regulation \| Metabolism Cell Cycle Checkpoints \| Cell cycle Centrosome maturation \| Cell cycle G beta:gamma signalling through PI3Kgamma \| Signal transduction PRC2 methylates histones and DNA \| Gene expression (Transcription) SUMOylation of RNA binding proteins \| Metabolism of proteins SLC transporter disorders \| Disease Nuclear Envelope (NE) Reassembly \| Cell cycle HCMV Late Events \| Disease SUMOylation of ubiquitinylation proteins \| Metabolism of proteins Microtubule-dependent trafficking of connexons from Golgi to the plasma membrane \| Vesicle-mediated transport tRNA processing \| Metabolism of RNA MHC class II antigen presentation \| Immune system Transport of connexons to the plasma membrane \| Vesicle-mediated transport Anchoring of the basal body to the plasma membrane \| Organelle biogenesis and maintenance HSP90 chaperone cycle for steroid hormone receptors (SHR) \| Cellular responses to external stimuli Influenza Viral RNA Transcription and Replication \| Disease Aggrephagy \| Autophagy SUMOylation of DNA replication proteins \| Metabolism of proteins HDACs deacetylate histones \| Chromatin organization Prefoldin mediated transfer of substrate to CCT/TriC \| Metabolism of proteins ADORA2B mediated anti-inflammatory cytokines production \| Disease Cellular responses to stress \| Cellular responses to external stimuli ISG15 antiviral mechanism \| Immune system HIV Life Cycle \| Disease Cellular responses to external stimuli \| Cellular responses to external stimuli Postmitotic nuclear pore complex (NPC) reformation \| Cell cycle Post-chaperonin tubulin folding pathway \| Metabolism of proteins Signaling by cytosolic PDGFRA and PDGFRB fusion proteins \| Disease Association of TriC/CCT with target proteins during biosynthesis \| Metabolism of proteins RHO GTPases activate IQGAPs \| Signal transduction Regulation of PTEN gene transcription \| Signal transduction Golgi-to-ER retrograde transport \| Vesicle-mediated transport G-protein beta:gamma signalling \| Signal transduction Activation of AMPK downstream of NMDARs \| Neuronal system |
| Hepatocellular injury/ Acute hepatitis/ liver failure | exocrine pancreatic insufficiency cholestasis pancreas disease liver disease biliary tract disease | PRMT5 RHOT2 EIF5B ARL15 RHOD ARL17B CNNM3 CNNM4 ARL13A C16orf13 MRPL35 MAT2A SEPT6 REV1 RAB1B NDUFS4 NME3 RAB9B NUBP2 RAB40A RAB40AL UXS1 TIMM8A TRMT2B SEPT10 RAB3C AK6 PPP1R21 MTIF2 AIFM1 NME4 RAB1A RAB40C TRIM23 POLE4 SMYD5 KIF2A FBXO41 DGUOK LOC100506422 NSF GALT DUT GNL3L NMRK1 FAM173B C9orf41 GSPT1 RRAGB CBWD2 ITPA OR4D10 RAB41 GSPT2 GNA14 GNAQ PAPD7 RUNDC3A ERAS NSUN2 RASEF IDNK ZCCHC6 ACSL1 TRAPPC11 SYN1 UPP2 TMEM252 METTL12 RABL2A ARL2 EHD1 RAB26 ATL3 SRL SEPT12 RALB EFTUD2 GNE RAB6C UPRT ARL5A ABCB7 TUT1 WTH3DI NUBP1 UGT3A1 CBWD5 CIITA TGM3 RND3 RRAGA RAB33A RHOQ ACSL6 LOC101929601 CHSY3 TBCEL HSPA8 AAGAB | RAB geranylgeranylation \| Metabolism of proteins Cytosolic iron-sulfur cluster assembly \| Metabolism Interconversion of nucleotide di- and triphosphates \| Metabolism Metabolism of nucleotides \| Metabolism of RNA COPI-dependent Golgi-to-ER retrograde traffic \| Vesicle-mediated transport Intra-Golgi and retrograde Golgi-to-ER traffic \| Vesicle-mediated transport Golgi-to-ER retrograde transport \| Vesicle-mediated transport Synthesis of very long-chain fatty acyl-CoAs \| Metabolism Rho GTPase cycle \| Signal transduction Chondroitin sulfate biosynthesis \| Metabolism RAB GEFs exchange GTP for GDP on RABs \| Vesicle-mediated transport Rab regulation of trafficking \| Vesicle-mediated transport Defective GALT can cause Galactosemia \| Disease RHO GTPases regulate CFTR trafficking \| Signal transduction "Defective GNE causes sialuria \| Nonaka myopathy and inclusion body myopathy 2" Diseases associated with glycosylation precursor biosynthesis \| Disease Eukaryotic Translation Termination \| Metabolism of proteins Nonsense Mediated Decay (NMD) independent of the Exon Junction Complex (EJC) \| Metabolism of RNA Fatty acyl-CoA biosynthesis \| Metabolism |
| Renal/Acute kidney failure or injury | membranous glomerulonephritis oligospermia female reproductive system disease renal artery obstruction IgA glomerulonephritis perineurioma prostate disease vasculogenic impotence Fanconi syndrome | GSN VPS41 WDR13 FKBP14 WDR33 FKBP7 SCIN FKBP11 WDR89 C2orf48 EML4 CDC20 FKBP10 FKBP1A FKBP3 DCAF8L2 CDC20B FKBP2 FKBP1B FKBP9 EIF2A CCT6B FKBP15 CORO1C EEF1D MKL1 PGLYRP3 PGLYRP4 AWAT1 PAN2 TLE1 PPP3R2 FKBP5 IL2RA TLE3 DCAF4L2 FKBP6 CD248 FKBP4 PACRGL RPTOR WRAP53 B4GALT2 IL9R B4GALT3 RENBP PGLYRP2 IL7R B4GALT4 TMEM81 AIP IL4R IL21R CLSTN3 NAGLU WDR38 NAGPA NOMO2 PGLYRP1 NEXN AZI2 AGPAT9 IL2RB NAGK NRG1 KCNA10 RETSAT KALRN KDELR3 LCNL1 RDH13 ITGB6 RANBP1 DHRS7 RDH8 NUP107 GC SPEG RBP4 PPP3R1 APH1A HRASLS5 CILP RDH11 PQLC3 TRPM8 MTTP TMEM150C RDH12 BTLA RDH14 RARRES1 RARRES3 HRASLS2 IL2RG FAM84B MAGEB5 ALPK3 ITGB1 SEPT10 | Keratan sulfate biosynthesis \| Metabolism Association of TriC/CCT with target proteins during biosynthesis \| Metabolism of proteins N-Glycan antennae elongation \| Metabolism of proteins Keratan sulfate/keratin metabolism \| Metabolism Interleukin receptor SHC signaling \| Immune system The canonical retinoid cycle in rods (twilight vision) \| Signal transduction RA biosynthesis pathway \| Signal transduction Interleukin-21 signaling \| Immune system Acyl chain remodelling of PE \| Metabolism HSF1-dependent transactivation \| Cellular responses to external stimuli N-glycan antennae elongation in the medial/trans-Golgi \| Metabolism of proteins RHO GTPases Activate Formins \| Signal transduction Retinoid metabolism disease events \| Disease GRB7 events in ERBB2 signaling \| Signal transduction Glycosaminoglycan metabolism \| Metabolism Calcineurin activates NFAT \| Immune system Signaling by Retinoic Acid \| Signal transduction Synthesis of UDP-N-acetyl-glucosamine \| Metabolism of proteins MPS IIIB - Sanfilippo syndrome B \| Disease TGFBR1 LBD Mutants in Cancer \| Disease Antimicrobial peptides \| Immune system ERBB2 Regulates Cell Motility \| Signal transduction Interleukin-2 signaling \| Immune system Molecules associated with elastic fibres \| Extracellular matrix organization Diseases of the neuronal system \| Disease Diseases associated with visual transduction \| Disease Retinoid cycle disease events \| Disease Chaperonin-mediated protein folding \| Metabolism of proteins Defective gamma-carboxylation of F9 \| Disease ERBB2 Activates PTK6 Signaling \| Signal transduction Attenuation phase \| Cellular responses to external stimuli Downregulation of ERBB2:ERBB3 signaling \| Signal transduction Loss of Function of TGFBR1 in Cancer \| Disease |
| Sepsis | hantavirus pulmonary syndrome hepatitis B hepatitis D hepatitis E X-linked hyper IgM syndrome genital herpes dientamoebiasis | TTLL10 RAB40C C16orf13 RHOT2 CHTF18 NME3 NUBP2 GNAI2 GNAT1 RAB26 SRL URGCP SEPT12 RHOA C3orf62 GNL3 GSPT1 UBA3 ISG20 KIF7 AP3S2 FHIT GDPGP1 HDDC3 ARF4 NME4 SPCS1 NUBP1 SEPT2 USB1 RAB5A TK2 DYNC1LI2 NAE1 RRAD NOL3 SYN2 ATG7 GHRL ARL8B VPS4A DTYMK NKIRAS1 KIFC3 DYNC1LI1 IP6K2 TREX1 NME6 KIF9 NPIPB3 TUFM KIF15 KIF22 PYCARD PAPD5 NOD2 GNAO1 ARL13B PRMT5 MRPL47 MFN1 SDR39U1 NUBPL ARHGAP5 POLE2 ARF6 RPL22L1 ATL1 RHOJ HSPA2 RAB15 ARL14 SMC4 ABCF3 REM2 RAB2B N4BP2L2 TRMT44 RFC3 VWA8 ARL11 TGDS ABCC4 RAP2A LIG4 RAB20 OSGEP LSG1 OPA1 EDDM3A GFM1 RAP2B ABCD4 EEFSEC RUVBL1 CKMT1A SPATA5L1 RABL3 ADPRH RAB8B | RAB geranylgeranylation \| Metabolism of proteins "Synthesis \| secretion RAB GEFs exchange GTP for GDP on RABs \| Vesicle-mediated transport Gap-filling DNA repair synthesis and ligation in GG-NER \| DNA repair Rab regulation of trafficking \| Vesicle-mediated transport PCNA-Dependent Long Patch Base Excision Repair \| DNA repair Recognition of DNA damage by PCNA-containing replication complex \| DNA repair Cooperation of PDCL (PhLP1) and TRiC/CCT in G-protein beta folding \| Metabolism of proteins Leading Strand Synthesis \| DNA replication Polymerase switching \| Cell cycle Resolution of AP sites via the multiple-nucleotide patch replacement pathway \| DNA repair Termination of translesion DNA synthesis \| DNA repair Dual Incision in GG-NER \| DNA repair G-protein activation \| Signal transduction Translesion synthesis by POLI \| DNA repair Gap-filling DNA repair synthesis and ligation in TC-NER \| DNA repair Polymerase switching on the C-strand of the telomere \| Cell cycle Translesion synthesis by POLK \| DNA repair Golgi-to-ER retrograde transport \| Vesicle-mediated transport Interconversion of nucleotide di- and triphosphates \| Metabolism Rho GTPase cycle \| Signal transduction Lagging Strand Synthesis \| DNA replication Defective LMBRD1 causes methylmalonic aciduria and homocystinuria type cblF \| Disease Dual incision in TC-NER \| DNA repair Cytosolic iron-sulfur cluster assembly \| Metabolism Defective ABCD4 causes MAHCJ \| Disease MHC class II antigen presentation \| Immune system Chaperonin-mediated protein folding \| Metabolism of proteins Defective CYP27A1 causes Cerebrotendinous xanthomatosis (CTX) \| Disease Eukaryotic Translation Termination \| Metabolism of proteins Neutrophil degranulation \| Immune system The NLRP1 inflammasome \| Immune system Translesion Synthesis by POLH \| DNA repair TBC/RABGAPs \| Vesicle-mediated transport Nonsense Mediated Decay (NMD) independent of the Exon Junction Complex (EJC) \| Metabolism of RNA |
| Bacteremia | papilloma pulmonary coin lesion hemoglobinuria chronic fatigue syndrome exanthem | RAB27A RAB27B AAGAB EHD4 RRAS RHOV ARL5B PRTFDC1 DLG1 POLD1 YME1L1 RHEBL1 TRMT44 IVD LSG1 OPA1 RAB18 MPP7 TRDMT1 RAB28 HAUS2 CKM SPATA5L1 GTPBP4 TUBAL3 RASL11B ARHGAP35 NUDT5 CKMT1A GUF1 RHOH EHD2 HSPA14 SUV39H2 RUVBL2 NOA1 BMS1 ABCF3 RAB6B SRPRB UBA5 ITPK1 RAB43 RAB7A EEFSEC RUVBL1 MKKS NDUFAF5 KIF16B PSMC1 RABL3 ADPRH GTPBP8 ITPA SAR1A NME9 RHOBTB1 MRPL47 MFN1 ADSSL1 KIF26A RPL22L1 TRMT61A CKB ARL14 GFM1 MRAS RAP2B HLTF SETD3 AK7 TGM3 ARL9 IRGC ACSL1 DNM2 HYKK NSUN2 RAB3D NDUFS6 SWSAP1 TRIP13 ASNA1 TRAPPC11 GCDH SCRG1 TRMT1 RAB8A MMAA FAM173B KIF2A LONP1 KIF7 RAB3C SMC2 PNPLA6 ARL15 RAB11B MRPL46 OR7G3 | RAB geranylgeranylation \| Metabolism of proteins RAB GEFs exchange GTP for GDP on RABs \| Vesicle-mediated transport Rab regulation of trafficking \| Vesicle-mediated transport Golgi-to-ER retrograde transport \| Vesicle-mediated transport Kinesins \| Hemostasis Creatine metabolism \| Metabolism tRNA modification in the nucleus and cytosol \| Metabolism of RNA COPI-dependent Golgi-to-ER retrograde traffic \| Vesicle-mediated transport Sema4D mediated inhibition of cell attachment and migration \| Developmental biology Factors involved in megakaryocyte development and platelet production \| Hemostasis Intra-Golgi and retrograde Golgi-to-ER traffic \| Vesicle-mediated transport tRNA processing \| Metabolism of RNA MHC class II antigen presentation \| Immune system TBC/RABGAPs \| Vesicle-mediated transport Negative regulation of NMDA receptor-mediated neuronal transmission \| Neuronal system CREB1 phosphorylation through NMDA receptor-mediated activation of RAS signaling \| Neuronal system "Unblocking of NMDA receptors \| glutamate binding and activation" Defective MMAA causes methylmalonic aciduria type cblA \| Disease Defective MUT causes methylmalonic aciduria mut type \| Disease Mitotic Prometaphase \| Cell cycle Separation of Sister Chromatids \| Cell cycle Rho GTPase cycle \| Signal transduction COPI-independent Golgi-to-ER retrograde traffic \| Vesicle-mediated transport Pyrophosphate hydrolysis \| Metabolism |
| Dermatologic complications/pressure ulcer | diffuse scleroderma acrodermatitis neurodermatitis hair disease pemphigus lichen planus | DGKE DCAF8L2 WDR89 TLE1 CDC20 ANXA1 MRPL32 MKL1 EIF2A TOMM5 FKBP9 GRP FKBP14 KCNA10 WDR33 FKBP3 IARS2 PACRGL AIFM1 IL2RA FKBP11 EEF1D FKBP1A SLC8A3 IL7R FKBP5 FKBP6 EML4 SLC8A1 OSBPL5 WDR13 SLC8A2 GSN FKBP1B FKBP15 TRPV1 FKBP4 AWAT1 CDC20B TCP11 C2orf48 PPP3CC BTAF1 DCAF4L2 FKBP2 FKBP10 FAM160A1 CORO1C TLE3 ORC3 PRKCD PIGQ OSBPL8 PPP3CB SCIN FKBP7 SQRDL IL4R PPP3CA ELOVL7 IL22RA2 NR3C2 AR PPP6C WDR38 PLA2G4C MRPL4 IL2RG IL21R ELOVL4 APH1A ELOVL3 PSD COX4I1 TSPAN13 RPTOR THOC2 TMEM81 TRPC4 TMEM185A ITGB7 PITPNM2 FBXO8 VPS41 ITGB1 PRLR IL9R PITPNA ERBB3 COX7A2 GBF1 PAN2 COX7A1 NR3C1 COX7A2L UGGT1 ITGB5 PITPNB ITGB3 CCT6B | Nuclear Receptor transcription pathway \| Gene expression (Transcription) Calcineurin activates NFAT \| Immune system Reduction of cytosolic Ca++ levels \| Hemostasis Sodium/Calcium exchangers \| Transport of small molecules HSP90 chaperone cycle for steroid hormone receptors (SHR) \| Cellular responses to external stimuli TRP channels \| Transport of small molecules Molecules associated with elastic fibres \| Extracellular matrix organization Interleukin-21 signaling \| Immune system Platelet calcium homeostasis \| Hemostasis HSF1-dependent transactivation \| Cellular responses to external stimuli Elastic fibre formation \| Extracellular matrix organization CLEC7A (Dectin-1) induces NFAT activation \| Immune system Synthesis of very long-chain fatty acyl-CoAs \| Metabolism Interleukin-4 and Interleukin-13 signaling \| Immune system Interleukin receptor SHC signaling \| Immune system Stimuli-sensing channels \| Transport of small molecules Acyl chain remodelling of PS \| Metabolism TGFBR1 LBD Mutants in Cancer \| Disease PI and PC transport between ER and Golgi membranes \| Metabolism Adrenaline signalling through Alpha-2 adrenergic receptor \| Hemostasis Ion homeostasis \| Muscle contraction Association of TriC/CCT with target proteins during biosynthesis \| Metabolism of proteins GABA synthesis \| Neuronal system |
| Ocular symptoms | age related macular degeneration non-suppurative otitis media serous glue ear oculomotor nerve paralysis acute serous otitis media hypertensive retinopathy presbyopia panuveitis granular corneal dystrophy posterior uveitis pars planitis intermediate uveitis uveitis scleritis excessive tearing chronic purulent otitis media chronic atticoantral disease chronic tubotympanic suppurative otitis media macular corneal dystrophy corneal dystrophy retinitis acute transudative otitis media external ear disease cystoid macular edema macular degeneration macular retinal edema keratitis ophthalmoplegia retinal disease ocular hypotension dacryoadenitis hyperopia corneal disease neuroretinitis | ITGB2 ITGB3 ITGB1 ITGB6 ITGB7 ITGB5 AR NR3C2 OSBPL8 NUP107 TMEM86B OSBPL5 AIFM1 PGR TSPAN13 ANXA1 SQRDL TRPC4 NR3C1 ORC3 KCNA10 NR4A3 GRP APH1A TRPV1 SLC8A1 TCP11 ICMT ARFGEF2 GBF1 FBXO8 PSD2 PTGR2 CYB561A3 PRLR ARFGEF1 OSBPL9 PSD3 PSD NR2E3 FAM26E OSBPL3 TMEM43 FAR1 FAR2 PSD4 CYP11B1 CLDN7 CDKN2B CHDC2 GPR97 LXN ELOVL7 APH1B NR2C1 ELOVL4 COPS5 ELOVL3 SLC8A3 ELOVL1 SLC8A2 COX7A2 RARRES1 COX7A1 CELA2B PPP2R1A PIGQ PPP2R1B OSBPL6 PMP22 IL22RA2 COX7A2L NR1I2 GPRC5A FAM160A1 SHBG GAS6 CCDC93 NR0B1 EGFLAM CSF2RA GAL3ST2 BTAF1 SF3B1 SRD5A2 IL2RG IL7R VANGL2 IL9R HSD3B7 IL2RB UGGT1 TLE3 WDR13 FKBP4 TMEM62 DCAF8L2 TECRL GSN DCAF4L2 | Nuclear Receptor transcription pathway \| Gene expression (Transcription) Synthesis of very long-chain fatty acyl-CoAs \| Metabolism Molecules associated with elastic fibres \| Extracellular matrix organization Elastic fibre formation \| Extracellular matrix organization Interleukin receptor SHC signaling \| Immune system Reduction of cytosolic Ca++ levels \| Hemostasis PP2A-mediated dephosphorylation of key metabolic factors \| Metabolism Sodium/Calcium exchangers \| Transport of small molecules Fatty acyl-CoA biosynthesis \| Metabolism HSP90 chaperone cycle for steroid hormone receptors (SHR) \| Cellular responses to external stimuli TRP channels \| Transport of small molecules E2F mediated regulation of DNA replication \| Cell cycle Signaling by GSK3beta mutants \| Disease ERKs are inactivated \| Signal transduction MASTL Facilitates Mitotic Progression \| Cell cycle Signaling by CTNNB1 phospho-site mutants \| Disease S37 mutants of beta-catenin aren't phosphorylated \| Disease S45 mutants of beta-catenin aren't phosphorylated \| Disease S33 mutants of beta-catenin aren't phosphorylated \| Disease T41 mutants of beta-catenin aren't phosphorylated \| Disease NOTCH4 Activation and Transmission of Signal to the Nucleus \| Signal transduction Platelet calcium homeostasis \| Hemostasis "Regulation of glycolysis by fructose 2 \| 6-bisphosphate metabolism" Beta-catenin phosphorylation cascade \| Signal transduction G1 Phase \| Cell cycle Cyclin D associated events in G1 \| Cell cycle Wax biosynthesis \| Metabolism ECM proteoglycans \| Extracellular matrix organization Inhibition of replication initiation of damaged DNA by RB1/E2F1 \| Cell cycle RHO GTPases Activate Formins \| Signal transduction Interleukin-4 and Interleukin-13 signaling \| Immune system Wax and plasmalogen biosynthesis \| Metabolism NOTCH2 Activation and Transmission of Signal to the Nucleus \| Signal transduction Acyl chain remodelling of PS \| Metabolism Defective CYP11B1 causes Adrenal hyperplasia 4 (AH4) \| Disease Adrenaline signalling through Alpha-2 adrenergic receptor \| Hemostasis Linoleic acid (LA) metabolism \| Metabolism |

**Table S2.** Hierarchically ranked comorbidities, comorbidity enriched MOA proteins and pathways for each COVID-19 clinical manifestation from the GWAS risk genes(2) as input results.

| **Complication** | **Comorbidities** | **Comorbidity enriched MOA proteins** | **Pathways \| Top Pathways** |
| --- | --- | --- | --- |
| Respiratory | respiratory failure ventilation pneumonitis adult respiratory distress syndrome pulmonary edema bagassosis bird fancier's lung farmer's lung malt worker's lung lower respiratory tract disease obstructive lung disease mushroom workers' lung asthma chronic obstructive pulmonary disease rhinitis vasomotor rhinitis chronic rhinitis cork-handlers' disease maple bark strippers' lung lung disease intrinsic asthma status asthmaticus allergic asthma pulmonary emphysema upper respiratory tract disease tonsillitis common cold nasopharyngitis | RGR GPR146 OR5AK2 DRD5 GPR119 OR8S1 CRHR1 OR5A1 OR51L1 OR52N2 AGTR1 PIK3R3 SUCNR1 F2RL2 OR2V2 CXCR6 NTF3 BDNF XCR1 NGF TACR3 VIPR1 OR5M10 TACR1 OR52I2 GPR25 NTF4 F2R OR51B4 OR52D1 RXFP2 OR5M11 GALR1 GLP1R CNR2 NPY2R GPR21 TAAR6 PRLHR GPR88 BRS3 TACR2 ANXA1 APLNR DRD1 OR5M1 ADRB2 G0S2 ADRB3 OR2T6 CTXN3 ADRB1 GIPR OR51A2 RXFP1 TAAR1 GPR6 TAAR9 OR10A7 GPR149 OR7C1 NR3C1 NR3C2 OR2B6 OR10G4 OR10G9 OR10G8 OR8G1 OR51A4 OR1E2 OR3A3 OR3A2 OR1M1 OR1D5 OR52A5 OR1C1 OR7C2 TAS2R14 TAS2R31 GCH1 OR4K15 PNP GPR148 OR51B6 OR2G3 OR4X2 OR14A16 HTR4 OR10Q1 GPBAR1 OR8U1 OR11L1 OR2W3 OR12D3 OR2T33 OR2T2 OR2G6 OR14I1 OR6C4 TAS2R41 | G alpha (s) signalling events \| Signal transduction Olfactory Signaling Pathway \| Signal transduction GPCR ligand binding \| Signal transduction Signaling by GPCR \| Signal transduction GPCR downstream signalling \| Signal transduction ADORA2B mediated anti-inflammatory cytokines production \| Disease Class A/1 (Rhodopsin-like receptors) \| Signal transduction Nuclear Receptor transcription pathway \| Gene expression (Transcription) Signal Transduction \| Signal transduction Anti-inflammatory response favouring Leishmania parasite infection \| Disease Leishmania parasite growth and survival \| Disease Peptide ligand-binding receptors \| Signal transduction Amine ligand-binding receptors \| Signal transduction Tachykinin receptors bind tachykinins \| Signal transduction Leishmania infection \| Disease Relaxin receptors \| Signal transduction Dopamine receptors \| Signal transduction Glucagon-type ligand receptors \| Signal transduction Class B/2 (Secretin family receptors) \| Signal transduction MECP2 regulates transcription of neuronal ligands \| Gene expression (Transcription) Adrenoceptors \| Signal transduction G alpha (q) signalling events \| Signal transduction Class C/3 (Metabotropic glutamate/pheromone receptors) \| Signal transduction Loss of MECP2 binding ability to 5mC-DNA \| Disease Activated NTRK2 signals through PLCG1 \| Signal transduction Activated NTRK2 signals through RAS \| Signal transduction NTF4 activates NTRK2 (TRKB) signaling \| Signal transduction NTF3 activates NTRK3 signaling \| Signal transduction Transfer of LPS from LBP carrier to CD14 \| Immune system NTF3 activates NTRK2 (TRKB) signaling \| Signal transduction BDNF activates NTRK2 (TRKB) signaling \| Signal transduction |
| Respiratory failure | respiratory failure | VEGFA LOC651959 LOC648044 PRRG3 FGF13 MBNL3 CT47A5 IL13RA2 TCEAL4 FGF16 BEND2 CSF2RA NPTXR LGALS1 LGALS2 EYA2 WFDC6 FLRT3 GNRH2 LOC100287534 ZNF837 CNOT3 CLEC11A FGF21 LGALS14 LGALS13 LGALS4 LGALS7B ZNF566 ETV2 LSM14A CALR3 CALR AP1M2 ZNF57 FGF22 SERPINB11 SIGLEC15 SLC14A2 GATA6 VAPA PRR29 AKAP1 COX11 CHAD LRRC59 STAT5A KRTAP2-4 LGALS9 LGALS9B LGALS9C FGF11 ASGR1 ZNF232 CAMTA2 VMO1 FOXF1 PKD1L3 CDH16 GP2 AMDHD2 CRTC3 FAM154B PAQR5 SCG3 FGF7 JAG2 SERPINA9 LGALS3 ZNF219 FGF14 PDX1 FGF9 ELK3 PLXNC1 RAB3IP IFNG WIF1 NFE2 KLRC1 KLRC2 KLRC4-KLRK1 KLRD1 OLR1 CLEC7A CLEC9A CLEC12B CLEC1B CLEC2A CLEC2B KLRF1 CD69 CLEC2D KLRB1 KLRG1 CLEC6A C11orf97 NEU3 FGF3 FGF4 | Olfactory Signaling Pathway \| Signal transduction G alpha (s) signalling events \| Signal transduction GPCR downstream signalling \| Signal transduction FGFR2 ligand binding and activation \| Signal transduction Activated point mutants of FGFR2 \| Disease FGFR4 ligand binding and activation \| Signal transduction FGFR2c ligand binding and activation \| Signal transduction Nuclear Receptor transcription pathway \| Gene expression (Transcription) Passive transport by Aquaporins \| Transport of small molecules TRAF6 mediated IRF7 activation \| Immune system Signaling by activated point mutants of FGFR3 \| Disease FGFR1 ligand binding and activation \| Signal transduction FGFRL1 modulation of FGFR1 signaling \| Signal transduction Signaling by activated point mutants of FGFR1 \| Disease FGFR3c ligand binding and activation \| Signal transduction FGFR3 ligand binding and activation \| Signal transduction FGFR1c ligand binding and activation \| Signal transduction FGFR2 mutant receptor activation \| Disease Neutrophil degranulation \| Immune system FGFR3 mutant receptor activation \| Disease Keratan sulfate degradation \| Metabolism Rhesus blood group biosynthesis \| Metabolism Peptide ligand-binding receptors \| Signal transduction Defective B4GALT1 causes B4GALT1-CDG (CDG-2d) \| Disease Defective ST3GAL3 causes MCT12 and EIEE15 \| Disease Defective CHST3 causes SEDCJD \| Disease "Defective CHST14 causes EDS \| musculocontractural type" Defective CHST6 causes MCDC1 \| Disease FGFR3b ligand binding and activation \| Signal transduction Dermatan sulfate biosynthesis \| Metabolism SHC-mediated cascade:FGFR2 \| Signal transduction Diseases associated with glycosaminoglycan metabolism \| Disease Defective CHSY1 causes TPBS \| Disease O-glycosylation of TSR domain-containing proteins \| Metabolism of proteins Nucleotide-like (purinergic) receptors \| Signal transduction Class A/1 (Rhodopsin-like receptors) \| Signal transduction FGFR1b ligand binding and activation \| Signal transduction Defective B3GAT3 causes JDSSDHD \| Disease Glycogen storage diseases \| Disease FGFR2b ligand binding and activation \| Signal transduction Defective B3GALTL causes Peters-plus syndrome (PpS) \| Disease Cellular hexose transport \| Transport of small molecules P2Y receptors \| Signal transduction Prostanoid ligand receptors \| Signal transduction SHC-mediated cascade:FGFR1 \| Signal transduction "Defective B4GALT7 causes EDS \| progeroid type" Defective B3GALT6 causes EDSP2 and SEMDJL1 \| Disease Signaling by GPCR \| Signal transduction Molecules associated with elastic fibres \| Extracellular matrix organization IRAK4 deficiency (TLR2/4) \| Disease SHC-mediated cascade:FGFR4 \| Signal transduction Transport of glycerol from adipocytes to the liver by Aquaporins \| Transport of small molecules Hydroxycarboxylic acid-binding receptors \| Signal transduction Diseases of glycosylation \| Disease RHO GTPases activate KTN1 \| Signal transduction CS/DS degradation \| Metabolism |
| Acute respiratory distress syndrome (ARDS) | Adult respiratory distress syndrome | GRP PIGQ VDR ANXA1 TSPAN13 NR3C1 NR4A3 APH1B NR3C2 SLC8A1 SLC8A2 SLC8A3 PGR ELOVL3 ELOVL7 PRLR KCNA10 ELOVL1 OSBPL8 OSBPL5 ELOVL4 ARFGEF1 ORC3 TMEM86B IL22RA2 ITGB6 ARFGEF2 ITGB3 ITGB1 PPP2R1A PPP2R1B CELA2B AR COX7A1 PLEKHG4 RLBP1 ITGB7 PLA2G16 HRASLS2 RARRES3 HRASLS5 BTAF1 RBP3 COX7A2 NUDCD2 FAM160A1 LXN ITGB5 COX7A2L FAM84A ITGB2 RDH13 RDH8 RDH12 RDH11 DHRS7 GPRC5A PPP6C FAM84B LRAT RARRES1 RDH14 OR1S2 OR1J1 KALRN TRPV1 SF3B1 RETSAT OSBPL9 NR0B1 EMP3 HRH4 OR1N1 TCP11 OR11L1 PTGER1 CRABP2 CRABP1 TMEM185A PSD2 FBXO8 OR14A16 APH1A CSF2RA NUP107 GBF1 RBP5 FABP4 FABP9 FABP5 FABP7 FABP2 RBP1 RBP2 FABP3 RBP7 ICMT AIFM1 CDC45 CYB561A3 | G alpha (s) signalling events \| Signal transduction Class A/1 (Rhodopsin-like receptors) \| Signal transduction GPCR ligand binding \| Signal transduction Signaling by GPCR \| Signal transduction Olfactory Signaling Pathway \| Signal transduction GPCR downstream signalling \| Signal transduction ADORA2B mediated anti-inflammatory cytokines production \| Disease Peptide ligand-binding receptors \| Signal transduction Nuclear Receptor transcription pathway \| Gene expression (Transcription) Amine ligand-binding receptors \| Signal transduction Na+/Cl- dependent neurotransmitter transporters \| Transport of small molecules Prostanoid ligand receptors \| Signal transduction Activation of Ca-permeable Kainate Receptor \| Neuronal system Ionotropic activity of kainate receptors \| Neuronal system Potassium Channels \| Neuronal system Voltage gated Potassium channels \| Neuronal system "Unblocking of NMDA receptors \| glutamate binding and activation" Chemokine receptors bind chemokines \| Signal transduction Activation of kainate receptors upon glutamate binding \| Neuronal system Adenosine P1 receptors \| Signal transduction Adrenaline signalling through Alpha-2 adrenergic receptor \| Hemostasis Histamine receptors \| Signal transduction Opsins \| Signal transduction G alpha (i) signalling events \| Signal transduction Eicosanoid ligand-binding receptors \| Signal transduction Reuptake of GABA \| Neuronal system The canonical retinoid cycle in rods (twilight vision) \| Signal transduction Activation of AMPA receptors \| Neuronal system TWIK-releated acid-sensitive K+ channel (TASK) \| Neuronal system E2F-enabled inhibition of pre-replication complex formation \| Cell cycle Leishmania parasite growth and survival \| Disease Anti-inflammatory response favouring Leishmania parasite infection \| Disease Hormone ligand-binding receptors \| Signal transduction Tachykinin receptors bind tachykinins \| Signal transduction Relaxin receptors \| Signal transduction NrCAM interactions \| Developmental biology Diseases associated with visual transduction \| Disease Diseases of the neuronal system \| Disease Retinoid cycle disease events \| Disease G alpha (q) signalling events \| Signal transduction RA biosynthesis pathway \| Signal transduction Adrenoceptors \| Signal transduction Activation of Na-permeable kainate receptors \| Neuronal system Tandem pore domain potassium channels \| Neuronal system ABC-family proteins mediated transport \| Transport of small molecules Orexin and neuropeptides FF and QRFP bind to their respective receptors \| Signal transduction ATP sensitive Potassium channels \| Neuronal system Muscarinic acetylcholine receptors \| Signal transduction Activation of ATR in response to replication stress \| Cell cycle TRP channels \| Transport of small molecules Serotonin receptors \| Signal transduction CDC6 association with the ORC:origin complex \| DNA replication Hydroxycarboxylic acid-binding receptors \| Signal transduction |
| Asthma exacerbation | asthma  allergic asthma intrinsic asthma  status asthmaticus | GPR19 SLC30A6 BDNF OR8S1 GPR146 RGR PSD2 FSHR TAS2R14 NR3C1 OR2T27 KCNK2 F2RL2 AGTR1 OR10A5 DRD1 OR5M11 OR10A7 F2R NTF4 TPH1 OR2T4 OR2T6 TACR1 GLP1R GPR151 OR8U1 HTR4 GPBAR1 NTF3 VIPR1 ADRB2 OR2V2 CXCR6 GPR6 ADRA2B PRKCD UGGT1 GPR148 TAAR1 TAAR5 TAAR6 CLDN6 TAAR9 OR5M10 GPR88 RXFP1 FBXO8 ITGB3 ANXA1 APLNR TH TSHR OR52I2 OR51A4 NGF OR51L1 RXFP2 CRHR1 PIK3R3 SH2B1 NR3C2 ADRB1 GPR21 CNR2 SLC18A2 DRD5 OR8U9 OR5A1 GPR78 ARFGEF2 OR51A2 GIPR GPR25 OR52N2 AIFM1 OR5M1 SH2D1B PIK3R1 LGR6 OR56A1 OR52J3 SUCNR1 GPR149 SQRDL OR51B6 OR51B4 SH2B3 TAS2R41 PAH ADRB3 GPR119 TPH2 LGR5 OR10V1 ADRA1B PDE6A PDE5A CTXN3 PDE8B | Olfactory Signaling Pathway \| Signal transduction Signaling by GPCR \| Signal transduction GPCR downstream signalling \| Signal transduction ADORA2B mediated anti-inflammatory cytokines production \| Disease Nuclear Receptor transcription pathway \| Gene expression (Transcription) GPCR ligand binding \| Signal transduction Amine ligand-binding receptors \| Signal transduction Leishmania parasite growth and survival \| Disease Anti-inflammatory response favouring Leishmania parasite infection \| Disease Class A/1 (Rhodopsin-like receptors) \| Signal transduction Leishmania infection \| Disease Adrenoceptors \| Signal transduction Relaxin receptors \| Signal transduction Dopamine receptors \| Signal transduction Signal Transduction \| Signal transduction Glucagon-type ligand receptors \| Signal transduction Hormone ligand-binding receptors \| Signal transduction Peptide ligand-binding receptors \| Signal transduction Metabolism of amine-derived hormones \| Metabolism MECP2 regulates transcription of neuronal ligands \| Gene expression (Transcription) Class B/2 (Secretin family receptors) \| Signal transduction Serotonin and melatonin biosynthesis \| Metabolism Interleukin receptor SHC signaling \| Immune system Regulation of FZD by ubiquitination \| Signal transduction TWIK related potassium channel (TREK) \| Neuronal system Phenylketonuria \| Disease Adrenaline signalling through Alpha-2 adrenergic receptor \| Hemostasis Loss of MECP2 binding ability to 5mC-DNA \| Disease Activated NTRK2 signals through PI3K \| Signal transduction Activated NTRK2 signals through PLCG1 \| Signal transduction NTF4 activates NTRK2 (TRKB) signaling \| Signal transduction NTF3 activates NTRK3 signaling \| Signal transduction Transfer of LPS from LBP carrier to CD14 \| Immune system UNC93B1 deficiency - HSE \| Disease Activated NTRK2 signals through RAS \| Signal transduction NTF3 activates NTRK2 (TRKB) signaling \| Signal transduction BDNF activates NTRK2 (TRKB) signaling \| Signal transduction SMAC(DIABLO)-mediated dissociation of IAP:caspase complexes \| Programmed cell death |
| Chronic obstructive pulmonary disease (COPD) exacerbation/Acute coronary syndromes | obstructive lung disease chronic obstructive pulmonary disease Dressler's syndrome | ADRA2B GPR50 TAAR6 GPR119 TAAR8 TAAR9 LGR6 OR51L1 OR8U1 GPR149 OR5M8 OR51A4 OR5M10 OR5M1 KCNK2 GPR6 TACR1 GLP1R TAS2R41 DRD5 CCKAR NGF OR56A1 VIPR1 OR12D2 TAAR5 OR5V1 BDNF NTSR2 CCKBR OR52N2 OR10A2 MC5R OR2T6 OPN3 OR51B6 PCSK1N TAAR1 MTNR1B ADRB3 TACR2 ADRB2 HTR4 RPS19 RXFP1 NTF4 PIK3R3 NTF3 TAS2R14 GPR19 RXFP2 SH2B3 CNR2 HTR6 LGR5 PIK3R1 OR10A7 PCSK1 GPR21 GIPR OR56A4 TACR3 OR8U9 RRH DRD1 ADRB1 RGR CD300LG OR1A2 OR1D5 NR4A3 OR13F1 NPBWR1 OR4D1 SLC35G5 CRHR1 GLP2R MC1R OR1A1 OR13D1 PDE1C OR1J1 OR1J4 PDE6G OR1E2 OR3A3 CD300LF GPR142 TAS2R5 PILRB OR9A4 MTAP OR6V1 THRA OR1E1 ANXA1 OR3A2 OR4D2 FZD1 PDE7A | G alpha (s) signalling events \| Signal transduction Olfactory Signaling Pathway \| Signal transduction GPCR ligand binding \| Signal transduction Signaling by GPCR \| Signal transduction GPCR downstream signalling \| Signal transduction ADORA2B mediated anti-inflammatory cytokines production \| Disease Class A/1 (Rhodopsin-like receptors) \| Signal transduction Amine ligand-binding receptors \| Signal transduction Anti-inflammatory response favouring Leishmania parasite infection \| Disease Leishmania parasite growth and survival \| Disease Signal Transduction \| Signal transduction Leishmania infection \| Disease Tachykinin receptors bind tachykinins \| Signal transduction Opsins \| Signal transduction Peptide ligand-binding receptors \| Signal transduction Class B/2 (Secretin family receptors) \| Signal transduction Glucagon-type ligand receptors \| Signal transduction Adrenoceptors \| Signal transduction Relaxin receptors \| Signal transduction Dopamine receptors \| Signal transduction Serotonin receptors \| Signal transduction Cam-PDE 1 activation \| Signal transduction MECP2 regulates transcription of neuronal ligands \| Gene expression (Transcription) G alpha (i) signalling events \| Signal transduction Interleukin receptor SHC signaling \| Immune system Regulation of FZD by ubiquitination \| Signal transduction TWIK related potassium channel (TREK) \| Neuronal system Activated NTRK2 signals through PI3K \| Signal transduction Class C/3 (Metabotropic glutamate/pheromone receptors) \| Signal transduction Adrenaline signalling through Alpha-2 adrenergic receptor \| Hemostasis Loss of MECP2 binding ability to 5mC-DNA \| Disease Activated NTRK2 signals through PLCG1 \| Signal transduction Nuclear Receptor transcription pathway \| Gene expression (Transcription) G alpha (q) signalling events \| Signal transduction Activated NTRK2 signals through RAS \| Signal transduction NTF4 activates NTRK2 (TRKB) signaling \| Signal transduction NTF3 activates NTRK3 signaling \| Signal transduction Transfer of LPS from LBP carrier to CD14 \| Immune system UNC93B1 deficiency - HSE \| Disease BDNF activates NTRK2 (TRKB) signaling \| Signal transduction NTF3 activates NTRK2 (TRKB) signaling \| Signal transduction |
| Cardiovascular/Arrhythmia | essential hypertension secondary hypertension heart disease hypertensive heart disease atrial fibrillation nephrosclerosis cardiac arrest cardiovascular system disease dilated cardiomyopathy carotid stenosis thoracic aortic aneurysm renovascular hypertension aortic valve stenosis mitral valve stenosis brain ischemia renal artery disease long QT syndrome ischemia coronary artery disease cerebral arterial disease aortic aneurysm Wolff-Parkinson-White syndrome heart valve disease lymphedema arteriosclerosis obliterans aortic disease myocardial infarction congestive heart failure tetralogy of Fallot cerebrovascular disease abdominal aortic aneurysm varicose veins intermediate coronary syndrome arrhythmogenic right ventricular cardiomyopathy strictly posterior acute myocardial infarction myocardial stunning cardiomyopathy CADASIL 1 CADASIL 2 atrioventricular block subendocardial infarction acute myocardial infarction left bundle branch hemiblock Dressler's syndrome acute inferoposterior infarction acute inferolateral myocardial infarction acute anterolateral myocardial infarction hypertension | F2RL2 OR51B6 LGR6 GPR25 APLNR XCR1 PTGDR2 CXCR6 RXFP2 SUCNR1 KISS1R GALR3 NPBWR1 ADRB2 OR52B4 PRLHR ADRB3 TAAR9 RXFP1 NPFFR1 CRHR1 GNRHR ADRB1 CNR2 OR5M1 GPR6 AGTR1 TACR3 TACR2 CCKBR OR52I2 TAS2R14 GPR21 OR5M10 ADRA2B GALR1 OR5H1 F2R OR12D2 TAAR6 TAAR5 TACR1 OR52N2 OR13A1 OR5V1 OR51L1 GIPR GPR149 GPR151 MTNR1B GHSR RRH NPY2R OR8U1 TAAR1 AVPR2 HTR4 GLP1R TAS2R41 NPBWR2 BRS3 GPR50 CLDN6 LGR5 GPR19 OR5M8 OR2T6 DRD5 GPR119 HTR6 OR8U9 PIK3R3 RGR HCRTR1 OR51A2 NTSR2 GALR2 OR52D1 GPR150 DRD1 OR51A4 OR5M11 MC5R GPR146 OR56A1 TNNT1 TNNT3 OR10A7 CCKAR GPR88 MC1R OR10A5 TNNT2 OR5AK2 OR10A2 GPBAR1 OR8S1 GPR148 PPAN-P2RY11 OR52J3 | G alpha (s) signalling events \| Signal transduction Olfactory Signaling Pathway \| Signal transduction Peptide ligand-binding receptors \| Signal transduction Class A/1 (Rhodopsin-like receptors) \| Signal transduction GPCR ligand binding \| Signal transduction Signaling by GPCR \| Signal transduction GPCR downstream signalling \| Signal transduction ADORA2B mediated anti-inflammatory cytokines production \| Disease Anti-inflammatory response favouring Leishmania parasite infection \| Disease Leishmania parasite growth and survival \| Disease Amine ligand-binding receptors \| Signal transduction Signal Transduction \| Signal transduction Leishmania infection \| Disease G alpha (q) signalling events \| Signal transduction G alpha (i) signalling events \| Signal transduction Tachykinin receptors bind tachykinins \| Signal transduction Orexin and neuropeptides FF and QRFP bind to their respective receptors \| Signal transduction Serotonin receptors \| Signal transduction Adrenoceptors \| Signal transduction Netrin mediated repulsion signals \| Developmental biology Striated Muscle Contraction \| Muscle contraction Relaxin receptors \| Signal transduction Opsins \| Signal transduction InlA-mediated entry of Listeria monocytogenes into host cells \| Disease Activated NTRK3 signals through PI3K \| Signal transduction Thrombin signalling through proteinase activated receptors (PARs) \| Hemostasis Regulation of commissural axon pathfinding by SLIT and ROBO \| Developmental biology Downregulation of ERBB4 signaling \| Signal transduction Dopamine receptors \| Signal transduction Long-term potentiation \| Neuronal system Regulation of RUNX1 Expression and Activity \| Gene expression (Transcription) Vasopressin-like receptors \| Signal transduction Listeria monocytogenes entry into host cells \| Disease MET activates PTK2 signaling \| Signal transduction MET promotes cell motility \| Signal transduction Downstream signal transduction \| Signal transduction Cargo recognition for clathrin-mediated endocytosis \| Vesicle-mediated transport Adrenaline signalling through Alpha-2 adrenergic receptor \| Hemostasis Signaling by PDGF \| Signal transduction Regulation of FZD by ubiquitination \| Signal transduction Signaling by NTRK3 (TRKC) \| Signal transduction Class C/3 (Metabotropic glutamate/pheromone receptors) \| Signal transduction Defective AVP does not bind AVPR2 and causes neurohypophyseal diabetes insipidus (NDI) \| Disease Arachidonate production from DAG \| Hemostasis Regulation of RUNX3 expression and activity \| Gene expression (Transcription) Glucagon-type ligand receptors \| Signal transduction Clathrin-mediated endocytosis \| Vesicle-mediated transport Class B/2 (Secretin family receptors) \| Signal transduction |
| Acute myocardial infarction/Unstable angina | coronary artery disease myocardial infarction intermediate coronary syndrome strictly posterior acute myocardial infarction subendocardial infarction acute myocardial infarction acute inferoposterior infarction acute inferolateral myocardial infarction acute anterolateral myocardial infarction | LGR6 GPR146 OR52D1 RXFP2 CCR9 ACKR3 GPR55 OR52B4 GPR4 F2RL2 PIK3R1 HTR4 F2R GPR150 OR10H5 PCSK1 PTGER1 GPR151 TAAR9 GPR6 MC5R GLP1R KISS1R OR12D2 OR5V1 OR7E24 PPAN-P2RY11 OR2V2 DRD1 ADRB2 GIPR RXFP1 GALR1 NPY2R AGTR1 SLC30A6 TACR1 ADRA2B GPBAR1 CXCR6 XCR1 NPBWR2 OR5H1 RRH PROKR2 SUCNR1 GPR149 GHSR TNNT1 GPR78 DRD5 GNRHR VN1R4 TAAR6 TACR3 TAAR8 GALR2 TAAR5 OR10A5 OR8U1 LTB4R OR4A5 TPH1 CCKBR OR5M11 OR56A1 OR52N2 OR51B6 OR52J3 OR51L1 OR51A2 OR5M8 OR5M10 TAAR1 PAH GPR19 TAS2R14 GPR84 OR10A7 LGR5 TPH2 MTNR1B OR5M1 SH2B3 PTGDR2 OR1S1 APLNR OR5AK2 OR51A4 OR52I2 OR2F2 CLDN6 NPBWR1 ADRB3 MC1R OR2A1 OR2F1 TAS2R41 TH ADORA2B | G alpha (s) signalling events \| Signal transduction Olfactory Signaling Pathway \| Signal transduction ADORA2B mediated anti-inflammatory cytokines production \| Disease Class A/1 (Rhodopsin-like receptors) \| Signal transduction GPCR ligand binding \| Signal transduction Signaling by GPCR \| Signal transduction GPCR downstream signalling \| Signal transduction Peptide ligand-binding receptors \| Signal transduction Signal Transduction \| Signal transduction Leishmania parasite growth and survival \| Disease Anti-inflammatory response favouring Leishmania parasite infection \| Disease Amine ligand-binding receptors \| Signal transduction Leishmania infection \| Disease G alpha (q) signalling events \| Signal transduction G alpha (i) signalling events \| Signal transduction Tachykinin receptors bind tachykinins \| Signal transduction Relaxin receptors \| Signal transduction Dopamine receptors \| Signal transduction Chemokine receptors bind chemokines \| Signal transduction Prostanoid ligand receptors \| Signal transduction Metabolism of amine-derived hormones \| Metabolism Adrenoceptors \| Signal transduction Eicosanoid ligand-binding receptors \| Signal transduction Serotonin and melatonin biosynthesis \| Metabolism Regulation of FZD by ubiquitination \| Signal transduction Phenylketonuria \| Disease Adrenaline signalling through Alpha-2 adrenergic receptor \| Hemostasis Adenosine P1 receptors \| Signal transduction Glucagon-type ligand receptors \| Signal transduction UNC93B1 deficiency - HSE \| Disease SMAC(DIABLO)-mediated dissociation of IAP:caspase complexes \| Programmed cell death |
| Acute congestive heart failure (CHF) | Congestive heart failure | TACR1 NPBWR1 NPBWR2 OR52B4 TACR3 KISS1R PTGDR2 CCKBR NPFFR1 OR13A1 F2RL2 GALR1 OR52I2 GPR25 GALR3 MTNR1B APLNR TACR2 GPR151 NPY2R GNRHR LGR6 CNR2 OR8U9 GIPR CLDN6 OR5M1 OR52J3 PRLHR ADRB1 RGR GPR21 RXFP1 AVPR2 GPR50 TAS2R14 OR5AK2 OR8U1 OR10A5 OR52N2 TAAR5 GLP1R ADRB2 HTR4 GPR78 SUCNR1 AGTR1 OR5H1 XCR1 ADRA2B OR2T6 HTR6 BRS3 MC5R MC1R OR51B6 ADRB3 GPR146 TAAR6 GPR6 OR5V1 F2R GPR149 CXCR6 GPR88 PIK3R3 CRHR1 RXFP2 OR8S1 OR51L1 OR51A4 GPR150 GPBAR1 HCRTR1 GALR2 OR5M10 OR5M8 OR56A1 OR52D1 OR51A2 TAAR1 TAAR9 OR12D2 NTSR2 GPR119 RRH DRD5 GPR148 TAS2R41 DRD1 PCSK1N OR10A7 OR5M11 GHSR OPN3 GPR19 PCSK1 LGR5 OR1S1 OR56A4 | GPCR ligand binding \| Signal transduction G alpha (s) signalling events \| Signal transduction Class A/1 (Rhodopsin-like receptors) \| Signal transduction Glutathione conjugation \| Metabolism Reversible hydration of carbon dioxide \| Metabolism PD-1 signaling \| Immune system Translocation of ZAP-70 to Immunological synapse \| Immune system Amine ligand-binding receptors \| Signal transduction GPCR downstream signalling \| Signal transduction Cross-presentation of soluble exogenous antigens (endosomes) \| Immune system ADORA2B mediated anti-inflammatory cytokines production \| Disease SCF-beta-TrCP mediated degradation of Emi1 \| Cell cycle Regulation of ornithine decarboxylase (ODC) \| Metabolism Vif-mediated degradation of APOBEC3G \| Disease Phosphorylation of CD3 and TCR zeta chains \| Immune system Regulation of RAS by GAPs \| Signal transduction Hh mutants are degraded by ERAD \| Disease Olfactory Signaling Pathway \| Signal transduction CDT1 association with the CDC6:ORC:origin complex \| DNA replication Hh mutants abrogate ligand secretion \| Disease Degradation of AXIN \| Signal transduction APC/C:Cdc20 mediated degradation of Securin \| Cell cycle Autodegradation of Cdh1 by Cdh1:APC/C \| Cell cycle Regulation of activated PAK-2p34 by proteasome mediated degradation \| Programmed cell death Orc1 removal from chromatin \| Cell cycle MHC class II antigen presentation \| Immune system CDK-mediated phosphorylation and removal of Cdc6 \| Cell cycle Asymmetric localization of PCP proteins \| Signal transduction Downstream TCR signaling \| Immune system Regulation of Apoptosis \| Programmed cell death Autodegradation of the E3 ubiquitin ligase COP1 \| Cell cycle Assembly of the pre-replicative complex \| DNA replication Ubiquitin Mediated Degradation of Phosphorylated Cdc25A \| Cell cycle p53-Independent DNA Damage Response \| Cell cycle p53-Independent G1/S DNA damage checkpoint \| Cell cycle Signaling by GPCR \| Signal transduction Cdc20:Phospho-APC/C mediated degradation of Cyclin A \| Cell cycle Collagen degradation \| Extracellular matrix organization Assembly of collagen fibrils and other multimeric structures \| Extracellular matrix organization Degradation of the extracellular matrix \| Extracellular matrix organization Peptide ligand-binding receptors \| Signal transduction Activation of NF-kappaB in B cells \| Immune system APC:Cdc20 mediated degradation of cell cycle proteins prior to satisfation of the cell cycle checkpoint \| Cell cycle NIK-->noncanonical NF-kB signaling \| Immune system Oxygen-dependent proline hydroxylation of Hypoxia-inducible Factor Alpha \| Cellular responses to external stimuli APC/C:Cdc20 mediated degradation of mitotic proteins \| Cell cycle Ubiquitin-dependent degradation of Cyclin D \| Cell cycle DNA Replication Pre-Initiation \| DNA replication Dectin-1 mediated noncanonical NF-kB signaling \| Immune system Switching of origins to a post-replicative state \| Cell cycle Presynaptic depolarization and calcium channel opening \| Neuronal system Defective CFTR causes cystic fibrosis \| Disease Degradation of GLI1 by the proteasome \| Signal transduction Cation-coupled Chloride cotransporters \| Transport of small molecules G2/M Checkpoints \| Cell cycle Metabolism of polyamines \| Metabolism Hedgehog ligand biogenesis \| Signal transduction APC/C:Cdh1 mediated degradation of Cdc20 and other APC/C:Cdh1 targeted proteins in late mitosis/early G1 \| Cell cycle Aflatoxin activation and detoxification \| Metabolism Degradation of DVL \| Signal transduction Negative regulation of NOTCH4 signaling \| Signal transduction SCF(Skp2)-mediated degradation of p27/p21 \| Cell cycle Serotonin receptors \| Signal transduction Collagen formation \| Extracellular matrix organization Degradation of GLI2 by the proteasome \| Signal transduction ABC transporter disorders \| Disease GLI3 is processed to GLI3R by the proteasome \| Signal transduction TCR signaling \| Immune system Downstream signaling events of B Cell Receptor (BCR) \| Immune system Opsins \| Signal transduction Adrenaline signalling through Alpha-2 adrenergic receptor \| Hemostasis Activation of APC/C and APC/C:Cdc20 mediated degradation of mitotic proteins \| Cell cycle Stabilization of p53 \| Cell cycle Synthesis of DNA \| DNA replication \| Cell cycle Cellular response to hypoxia \| Cellular responses to external stimuli Interleukin-33 signaling \| Immune system Collagen chain trimerization \| Extracellular matrix organization UCH proteinases \| Metabolism of proteins DNA Replication \| DNA replication Regulation of APC/C activators between G1/S and early anaphase \| Cell cycle Separation of Sister Chromatids \| Cell cycle Phase II - Conjugation of compounds \| Metabolism Anti-inflammatory response favouring Leishmania parasite infection \| Disease Leishmania parasite growth and survival \| Disease FBXL7 down-regulates AURKA during mitotic entry and in early mitosis \| Cell cycle Adrenoceptors \| Signal transduction ABC-family proteins mediated transport \| Transport of small molecules Surfactant metabolism \| Metabolism of proteins The role of GTSE1 in G2/M progression after G2 checkpoint \| Cell cycle TWIK-releated acid-sensitive K+ channel (TASK) \| Neuronal system Tachykinin receptors bind tachykinins \| Signal transduction Relaxin receptors \| Signal transduction TNFR2 non-canonical NF-kB pathway \| Immune system Rhesus blood group biosynthesis \| Metabolism Vpu mediated degradation of CD4 \| Disease Inactivation of CDC42 and RAC1 \| Developmental biology Regulation of RUNX3 expression and activity \| Gene expression (Transcription) PCP/CE pathway \| Signal transduction Tandem pore domain potassium channels \| Neuronal system Crosslinking of collagen fibrils \| Extracellular matrix organization Adenosine P1 receptors \| Signal transduction Dopamine receptors \| Signal transduction Generation of second messenger molecules \| Immune system NCAM1 interactions \| Developmental biology RUNX1 regulates transcription of genes involved in differentiation of HSCs \| Gene expression (Transcription) Muscarinic acetylcholine receptors \| Signal transduction p53-Dependent G1/S DNA damage checkpoint \| Cell cycle p53-Dependent G1 DNA Damage Response \| Cell cycle Regulation of expression of SLITs and ROBOs \| Developmental biology Activation of Matrix Metalloproteinases \| Extracellular matrix organization Signaling by NOTCH4 \| Signal transduction Glutathione synthesis and recycling \| Metabolism Antigen processing-Cross presentation \| Immune system G1/S DNA Damage Checkpoints \| Cell cycle FCERI mediated NF-kB activation \| Immune system Antigen processing: Ubiquitination & Proteasome degradation \| Immune system Phase 4 - resting membrane potential \| Muscle contraction Erythrocytes take up oxygen and release carbon dioxide \| Transport of small molecules |
| Hematologic | sickle cell anemia hemorrhagic disease myofascial pain syndrome autoimmune lymphoproliferative syndrome methemoglobinemia | ACKR3 CCR9 OR2A1 ACKR2 LTB4R HCRTR1 OR13A1 LOC100996758 GPR50 NPFFR1 GPR65 CHRM2 GPR55 SLC6A4 GPBAR1 SSTR2 CRHR1 PTGER3 GPR4 CCR6 OR10H5 OR10H2 CXCR2 OR2AE1 KISS1R GALR1 GPR88 GALR2 GPR21 OR2A7 GPR25 OR5M11 OR5M1 OR5AK2 APLNR PTGDR2 HCAR1 HCAR2 OR4S2 CMKLR1 OR13G1 LPAR5 OR5M8 OR4A5 OPRK1 FSHR CXCR6 GPR148 GPR132 OR52J3 LHCGR MRGPRX2 SSTR1 OR52D1 OXER1 OR4K5 OR4Q3 GPR183 P2RY13 AGTR1 P2RY10 RRH OPRM1 SSTR3 CNR2 NPY2R OR2V2 VN1R4 OR5V1 NMBR F2R GPR128 F2RL2 OPRL1 BRS3 GPR78 SUCNR1 GPR34 OR52A5 ADRA2B GHSR GPR150 OR52N2 PRLHR OR52B4 MRGPRX3 RGR MRGPRX4 MRGPRX1 CARTPT TACR1 OR51Q1 OR51I2 P2RY1 P2RY12 OR51B4 OR52I2 OR51E2 OR51G2 OR51A4 | Olfactory Signaling Pathway \| Signal transduction Peptide ligand-binding receptors \| Signal transduction Class A/1 (Rhodopsin-like receptors) \| Signal transduction GPCR ligand binding \| Signal transduction G alpha (s) signalling events \| Signal transduction Signaling by GPCR \| Signal transduction GPCR downstream signalling \| Signal transduction G alpha (i) signalling events \| Signal transduction Signal Transduction \| Signal transduction G alpha (q) signalling events \| Signal transduction Chemokine receptors bind chemokines \| Signal transduction P2Y receptors \| Signal transduction Nucleotide-like (purinergic) receptors \| Signal transduction Hydroxycarboxylic acid-binding receptors \| Signal transduction Eicosanoid ligand-binding receptors \| Signal transduction ADORA2B mediated anti-inflammatory cytokines production \| Disease Opsins \| Signal transduction Orexin and neuropeptides FF and QRFP bind to their respective receptors \| Signal transduction Prostanoid ligand receptors \| Signal transduction Hormone ligand-binding receptors \| Signal transduction MECP2 regulates neuronal receptors and channels \| Gene expression (Transcription) Adrenaline signalling through Alpha-2 adrenergic receptor \| Hemostasis Muscarinic acetylcholine receptors \| Signal transduction |
| Neurologic | Asperger syndrome Gilles de la Tourette syndrome autonomic nervous system disease idiopathic peripheral autonomic neuropathy Riley-Day syndrome facioscapulohumeral muscular dystrophy carpal tunnel syndrome Lewy body dementia juvenile spinal muscular atrophy neurodegenerative disease facial neuralgia Parkinson's disease critical illness polyneuropathy neuroleptic malignant syndrome reflex sympathetic dystrophy encephalomalacia focal epilepsy reflex epilepsy Parkinson's disease 19A Parkinson's disease 4 Parkinson's disease 23 early myoclonic encephalopathy complex regional pain syndrome Lambert-Eaton myasthenic syndrome congenital myasthenic syndrome femoral neuropathy multiple system atrophy movement disease anterior horn cell disease juvenile myoclonic epilepsy essential tremor blepharospasm sleep disorder restless legs syndrome dystonia peripheral nervous system disease muscular atrophy epilepsy with generalized tonic-clonic seizures chronic fatigue syndrome nervous system disease narcolepsy lingual-facial-buccal dyskinesia Charcot-Marie-Tooth disease type 2 generalized dystonia focal hand dystonia sleep apnea obstructive sleep apnea Alzheimer's disease | ACKR2 XCR1 CCR9 GPR65 CCR6 GPR4 OR2A1 APLNR BRS3 F2RL2 GPR34 SUCNR1 AGTR1 NPFFR1 GPR50 OXER1 PTGDR2 CXCR2 OR4A5 OR5M10 SLC6A6 SLC6A2 LOC100996758 OR13A1 GPR55 SLC6A5 SLC6A1 GPR25 CXCR6 F2R GALR3 SLC6A13 TMEM64 RNPEP SSTR3 NPY2R SLC6A4 OR6B2 GPR171 OR1S1 SLC16A13 OR52B4 CRHR1 GPR35 EPYC TACR1 OR10H5 KISS1R PRLHR ACKR4 VN1R4 GALR1 OR2A7 CCR7 NMBR OR51L1 SLC6A11 TACR2 AVPR2 ACKR3 GPR132 SSTR1 LTB4R OR10H2 PTGER1 SLC6A19 OR7A17 KCNK9 SLC6A12 KCNK3 FARSB HCRTR2 OPRM1 LPAR5 SLC6A9 SLC6A3 PTH2 PROKR1 SSTR2 SLC6A15 GPR128 MC1R OR12D2 SLC6A16 NPBWR1 GALR2 PROKR2 SLC6A7 SLC6A18 OR2AT4 OR2T1 CNR2 SLC6A14 GIPR PIK3R3 SLC6A17 SLC6A20 OR1J2 GHSR GNRHR | Na+/Cl- dependent neurotransmitter transporters \| Transport of small molecules Peptide ligand-binding receptors \| Signal transduction Class A/1 (Rhodopsin-like receptors) \| Signal transduction GPCR ligand binding \| Signal transduction Signaling by GPCR \| Signal transduction GPCR downstream signalling \| Signal transduction G alpha (q) signalling events \| Signal transduction Chemokine receptors bind chemokines \| Signal transduction G alpha (s) signalling events \| Signal transduction Olfactory Signaling Pathway \| Signal transduction G alpha (i) signalling events \| Signal transduction "Transport of bile salts and organic acids \| metal ions and amine compounds" Reuptake of GABA \| Neuronal system Signal Transduction \| Signal transduction Amino acid transport across the plasma membrane \| Transport of small molecules SLC transporter disorders \| Disease TWIK-releated acid-sensitive K+ channel (TASK) \| Neuronal system SLC-mediated transmembrane transport \| Transport of small molecules Creatine metabolism \| Metabolism Eicosanoid ligand-binding receptors \| Signal transduction Tachykinin receptors bind tachykinins \| Signal transduction Transport of inorganic cations/anions and amino acids/oligopeptides \| Transport of small molecules ADORA2B mediated anti-inflammatory cytokines production \| Disease Disorders of transmembrane transporters \| Disease Tandem pore domain potassium channels \| Neuronal system Orexin and neuropeptides FF and QRFP bind to their respective receptors \| Signal transduction Prostanoid ligand receptors \| Signal transduction Phase 4 - resting membrane potential \| Muscle contraction Defective SLC6A18 may confer susceptibility to iminoglycinuria and/or hyperglycinuria \| Disease Defective SLC6A18 may confer susceptibility to iminoglycinuria and/or hyperglycinuria \| Disease Defective SLC6A3 causes Parkinsonism-dystonia infantile (PKDYS) \| Disease Defective SLC6A3 causes Parkinsonism-dystonia infantile (PKDYS) \| Disease Defective SLC6A2 causes orthostatic intolerance (OI) \| Disease Defective SLC6A5 causes hyperekplexia 3 (HKPX3) \| Disease Defective AVP does not bind AVPR2 and causes neurohypophyseal diabetes insipidus (NDI) \| Disease Class B/2 (Secretin family receptors) \| Signal transduction |
| Cerebral ischemia/infarction | carotid stenosis thoracic aortic aneurysm brain ischemia renal artery disease cerebral arterial disease aortic aneurysm lymphedema arteriosclerosis obliterans cerebrovascular disease abdominal aortic aneurysm varicose veins CADASIL 1 CADASIL 2 | F2RL2 AGTR1 GHSR NPBWR1 SUCNR1 GPR151 KISS1R APLNR GPR25 GALR3 CXCR6 PRLHR PTGDR2 XCR1 AVPR2 OR5M10 TACR3 NPFFR1 TACR2 GNRHR OR51B6 TACR1 GALR1 F2R CRHR1 VCAM1 GJA1 TNNI2 HCRTR1 NPBWR2 GALR2 GPR6 GLP1R RXFP2 GIPR OR52D1 OR5M1 NPY2R CCKBR OR52B4 OR51L1 OR52J3 OR52N2 TNNT3 ADRB2 OR10A2 RXFP1 TAAR6 OR12D2 OR13A1 MTNR1B OR5M8 TAAR9 OR52I2 OR5H1 CNR2 TAS2R14 ADRB3 TNNI1 THRA TNNT2 TNNT1 BRS3 LGR6 GPR149 ADRB1 INS MC5R BCAN SLC35G3 GPR68 ACKR3 ATP5D C1orf87 OR5V1 LPAR6 TAAR1 TAAR5 INS-IGF2 OR8U9 INSL3 RAB11FIP1 HTR4 OR56A1 NPPB OR2T6 GLRA1 GPR88 AGTR2 DRD1 GPR21 CCR9 GPR15 HTR6 GPR150 RGR GPR4 KCNA6 LPAR4 TAS2R41 | G alpha (s) signalling events \| Signal transduction Peptide ligand-binding receptors \| Signal transduction Class A/1 (Rhodopsin-like receptors) \| Signal transduction GPCR ligand binding \| Signal transduction Signaling by GPCR \| Signal transduction GPCR downstream signalling \| Signal transduction ADORA2B mediated anti-inflammatory cytokines production \| Disease Leishmania parasite growth and survival \| Disease Anti-inflammatory response favouring Leishmania parasite infection \| Disease Olfactory Signaling Pathway \| Signal transduction Amine ligand-binding receptors \| Signal transduction Leishmania infection \| Disease Signal Transduction \| Signal transduction G alpha (q) signalling events \| Signal transduction Striated Muscle Contraction \| Muscle contraction Tachykinin receptors bind tachykinins \| Signal transduction Relaxin receptors \| Signal transduction G alpha (i) signalling events \| Signal transduction Chemokine receptors bind chemokines \| Signal transduction Orexin and neuropeptides FF and QRFP bind to their respective receptors \| Signal transduction Serotonin receptors \| Signal transduction Adrenoceptors \| Signal transduction Cargo recognition for clathrin-mediated endocytosis \| Vesicle-mediated transport Defective AVP does not bind AVPR2 and causes neurohypophyseal diabetes insipidus (NDI) \| Disease P2Y receptors \| Signal transduction Glucagon-type ligand receptors \| Signal transduction Nucleotide-like (purinergic) receptors \| Signal transduction Class B/2 (Secretin family receptors) \| Signal transduction IRS activation \| Signal transduction Defective CHST3 causes SEDCJD \| Disease "Defective CHST14 causes EDS, musculocontractural type \| Disease |
| Endocrine | familial hyperlipidemia hyperlipoproteinemia type IV morbid obesity Conn's syndrome obsolete Cushing's syndrome diabetic polyneuropathy nodular goiter hyperinsulinemic hypoglycemia alpha 1-antitrypsin deficiency Addison's disease abdominal obesity-metabolic syndrome 1 hypogonadism xanthomatosis empty sella syndrome primary hyperaldosteronism cerebrotendinous xanthomatosis premature ovarian failure hyperthyroidism amino acid metabolic disorder diabetes mellitus diabetic neuropathy obesity hypoglycemia familial partial lipodystrophy adrenal cortical hypofunction premature menopause | OR13A1 GPR4 PROKR2 F2RL2 NPBWR2 SUCNR1 CCR9 OR1S1 GPER1 OR2A1 OR10H2 PTGER1 NMBR GPR171 KALRN BRS3 XCR1 OR52B4 NPFFR1 GPR35 OR4A5 COX7A2 GPR88 OR2AT4 NPBWR1 COX7A1 CXCR2 ACKR4 GPR142 AGTR1 OR1J2 GPR65 OR10H5 GPR21 OR1F1 GALR3 GPR31 APLNR EDNRB COX7A2L CELA2B NR1I3 OPRL1 GPR139 CRHR1 OR2A7 HCRTR2 OR10A7 PTGER3 NPY2R SSTR3 ADORA2A VN1R4 GALR1 OR6B2 CXCR6 GPR174 MC1R CXCR5 AVPR2 MRGPRX3 ACKR3 OR5AK2 MAS1 ACKR2 KISS1R FPR2 OR52M1 CCR6 OPRM1 CCR4 LOC100996758 OXER1 LPAR5 AR GHSR OR7A17 SSTR2 OR10H3 GPR52 SCTR OR2F1 OR7G2 NR4A3 SHBG MRGPRX1 MC3R OR2V2 MAS1L SLC8A3 LTB4R GRPR P2RY6 HCAR3 MRGPRF PLEKHG4 OR51B6 CCR3 PTAFR OR2K2 | Peptide ligand-binding receptors \| Signal transduction Class A/1 (Rhodopsin-like receptors) \| Signal transduction GPCR ligand binding \| Signal transduction Signaling by GPCR \| Signal transduction GPCR downstream signalling \| Signal transduction G alpha (s) signalling events \| Signal transduction Olfactory Signaling Pathway \| Signal transduction Chemokine receptors bind chemokines \| Signal transduction G alpha (i) signalling events \| Signal transduction G alpha (q) signalling events \| Signal transduction Signal Transduction \| Signal transduction Nuclear Receptor transcription pathway \| Gene expression (Transcription) Eicosanoid ligand-binding receptors \| Signal transduction ADORA2B mediated anti-inflammatory cytokines production \| Disease Orexin and neuropeptides FF and QRFP bind to their respective receptors \| Signal transduction Prostanoid ligand receptors \| Signal transduction Formyl peptide receptors bind formyl peptides and many other ligands \| Signal transduction Defective AVP does not bind AVPR2 and causes neurohypophyseal diabetes insipidus (NDI) \| Disease Adenosine P1 receptors \| Signal transduction Nucleotide-like (purinergic) receptors \| Signal transduction Hydroxycarboxylic acid-binding receptors \| Signal transduction |
| Gastrointestinal symptoms | ischemic colitis microscopic colitis collagenous colitis gastroparesis alexia sexual dysfunction constipation papilloma disease hyperglycemia intestinal disease esophageal disease stomach disease intestinal obstruction chronic fatigue syndrome atrophic gastritis irritable bowel syndrome | GPR65 GPR132 OR51F2 GPR4 HCAR1 CCR9 ACKR2 ACKR3 CCR6 LPAR5 CX3CR1 OR51G2 OR51E2 OR4P4 NMBR OR51M1 MAS1 CCR1 CCRL2 OR4K5 LTB4R MRGPRF OR4S2 OR6K6 OR10S1 GPR55 PTGER3 GPR31 CCR2 CCR5 OR2A4 CMKLR1 FFAR3 FFAR2 ELTD1 GPR37 GPR151 OR2AT4 GPR152 FSHR OXER1 LHCGR GPR183 OR4Q3 MRGPRD GPR32 EDNRB CXCR3 OR10H5 CCR10 CYSLTR1 PTGER1 P2RY10 TRHR MCHR2 OR2A7 HCAR3 OR2C3 OR4A5 OR2T27 TMEM116 VN1R4 OPRK1 OR1S1 GPR20 OPRM1 SSTR4 CCR4 XCR1 EDNRA C5AR1 HCAR2 LOC100996758 OR2A1 GPR50 GPR171 AGTR1 HCRTR1 GPR17 MAS1L HCRTR2 AGTR2 GPR174 MLNR OR8I2 LPAR4 LPAR6 CCR3 OPRL1 OR10A5 OR51D1 VN1R5 OR2Z1 OR10G7 OR51G1 F2RL3 TAS2R19 P2RY8 OR51Q1 MCHR1 | Olfactory Signaling Pathway \| Signal transduction Peptide ligand-binding receptors \| Signal transduction Class A/1 (Rhodopsin-like receptors) \| Signal transduction GPCR ligand binding \| Signal transduction G alpha (s) signalling events \| Signal transduction Signaling by GPCR \| Signal transduction GPCR downstream signalling \| Signal transduction Chemokine receptors bind chemokines \| Signal transduction G alpha (q) signalling events \| Signal transduction Signal Transduction \| Signal transduction G alpha (i) signalling events \| Signal transduction Eicosanoid ligand-binding receptors \| Signal transduction Hydroxycarboxylic acid-binding receptors \| Signal transduction Leukotriene receptors \| Signal transduction P2Y receptors \| Signal transduction Nucleotide-like (purinergic) receptors \| Signal transduction Interleukin-10 signaling \| Immune system Orexin and neuropeptides FF and QRFP bind to their respective receptors \| Signal transduction LTC4-CYSLTR mediated IL4 production \| Disease Prostanoid ligand receptors \| Signal transduction Hormone ligand-binding receptors \| Signal transduction MECP2 regulates neuronal receptors and channels \| Gene expression (Transcription) ADORA2B mediated anti-inflammatory cytokines production \| Disease Beta defensins \| Immune system Inhibition of nitric oxide production \| Disease Defective GIF causes intrinsic factor deficiency \| Disease |
| Hepatocellular injury/ Acute hepatitis/ liver failure | hepatorenal syndrome alcoholic liver cirrhosis intrahepatic cholestasis drug-induced hepatitis acute pancreatitis coronary artery disease liver disease pancreatitis short bowel syndrome blind loop syndrome | GPR35 CCR9 CXCR2 NMBR GPR25 CCR7 APLNR OR52J3 OR10A5 GPBAR1 OR51A4 OR52I2 OR10H5 OR8S1 CCR6 F2RL2 GPR151 AGTR1 GPR142 OR2A1 OR51F2 F2R OR4A5 XCR1 VN1R4 ACKR3 NPBWR1 PTGDR2 GLP2R GPR78 GPR4 GPR132 OR2V2 GPER1 OR56B4 OR52B4 GPR65 OR51L1 RGR OR51A2 TACR1 GPR171 GPR26 GPR87 PRLHR NPY1R MRGPRF OR2T11 SUCNR1 PTGER3 AVPR2 P2RY6 P2RY2 CCRL2 MRGPRD ADORA2B LHCGR OR1N1 CMKLR1 GPR182 RBP4 CXCR3 CYSLTR2 CYSLTR1 GALR3 LPAR4 GPR146 GALR2 P2RY10 RXFP2 PTGER1 OR13A1 ACKR4 NPFFR1 BRS3 OR2K2 GPR18 ELTD1 OPRM1 OR10G8 TAS2R41 LTB4R P2RY8 LPAR5 TAS2R19 PROKR2 GPR15 OR51B4 GPR174 GPR183 SSTR3 HCAR1 OR4S2 OR51V1 TACR3 CCR3 CCR5 GALR1 PTAFR CCKBR | Peptide ligand-binding receptors \| Signal transduction Class A/1 (Rhodopsin-like receptors) \| Signal transduction GPCR ligand binding \| Signal transduction G alpha (s) signalling events \| Signal transduction Signaling by GPCR \| Signal transduction GPCR downstream signalling \| Signal transduction Signal Transduction \| Signal transduction Olfactory Signaling Pathway \| Signal transduction G alpha (q) signalling events \| Signal transduction Chemokine receptors bind chemokines \| Signal transduction G alpha (i) signalling events \| Signal transduction ADORA2B mediated anti-inflammatory cytokines production \| Disease Eicosanoid ligand-binding receptors \| Signal transduction Nucleotide-like (purinergic) receptors \| Signal transduction Prostanoid ligand receptors \| Signal transduction P2Y receptors \| Signal transduction Leukotriene receptors \| Signal transduction Anti-inflammatory response favouring Leishmania parasite infection \| Disease Leishmania parasite growth and survival \| Disease Tachykinin receptors bind tachykinins \| Signal transduction LTC4-CYSLTR mediated IL4 production \| Disease Retinoid metabolism disease events \| Disease Interleukin-10 signaling \| Immune system Leishmania infection \| Disease Defective AVP does not bind AVPR2 and causes neurohypophyseal diabetes insipidus (NDI) \| Disease Adenosine P1 receptors \| Signal transduction Glucagon-type ligand receptors \| Signal transduction Hydroxycarboxylic acid-binding receptors \| Signal transduction |
| Renal/Acute kidney failure or injury | nephrosclerosis enterocele interstitial cystitis ureterolithiasis chronic interstitial cystitis endometriosis nephrolithiasis urethral disease breast fibrocystic disease | ACKR4 CXCR3 OR13C8 GPR174 FFAR2 GPR82 GPER1 SSTR4 OR7G2 EDNRB PROKR1 GRPR CXCR2 NPY1R GPR83 GPR35 MRGPRX1 OR10H5 MRGPRX3 CCR9 MRGPRF GPR87 MAS1L OR13A1 OPRM1 PTGER1 F2RL3 GPR1 OR4X1 GPR85 VN1R5 GPR20 OR4Q3 AVPR1B HCAR3 AGTR1 GPR15 OR4A5 CRHR1 EDNRA CCR7 MRGPRD OR2A4 OXER1 CXCR5 GPR182 OXGR1 GPR37 GPR18 FPR1 MRGPRX4 MCHR1 CCR4 OR1I1 LHCGR NPFFR1 GPR31 BDKRB1 OR51L1 OR2A7 PTGER3 ADCYAP1R1 CCR3 AVPR1A OR2K2 NTSR1 TAS2R19 LPAR4 OR4K1 CYSLTR1 ASPH P2RY2 GPR151 OR13C4 OR4K14 OR6B1 CDC25B GPR171 PTGFR OR4S2 NMUR2 ELTD1 OR2AE1 LTB4R GPR27 SUCNR1 OR4P4 SEC14L3 CCDC117 TMEM116 CYSLTR2 GPR17 OR10H2 TRIM13 ATP6 POMC OR51E2 QRFPR SEC14L4 UFL1 | Peptide ligand-binding receptors \| Signal transduction Class A/1 (Rhodopsin-like receptors) \| Signal transduction GPCR ligand binding \| Signal transduction G alpha (s) signalling events \| Signal transduction Signaling by GPCR \| Signal transduction GPCR downstream signalling \| Signal transduction Olfactory Signaling Pathway \| Signal transduction G alpha (q) signalling events \| Signal transduction G alpha (i) signalling events \| Signal transduction Eicosanoid ligand-binding receptors \| Signal transduction Chemokine receptors bind chemokines \| Signal transduction Signal Transduction \| Signal transduction Prostanoid ligand receptors \| Signal transduction ADORA2B mediated anti-inflammatory cytokines production \| Disease Leukotriene receptors \| Signal transduction "Defective AVP does not bind AVPR1A \| B and causes neurohypophyseal diabetes insipidus (NDI)" Anti-inflammatory response favouring Leishmania parasite infection \| Disease Leishmania parasite growth and survival \| Disease P2Y receptors \| Signal transduction Orexin and neuropeptides FF and QRFP bind to their respective receptors \| Signal transduction Vasopressin-like receptors \| Signal transduction LTC4-CYSLTR mediated IL4 production \| Disease Nucleotide-like (purinergic) receptors \| Signal transduction Formyl peptide receptors bind formyl peptides and many other ligands \| Signal transduction TGFBR1 LBD Mutants in Cancer \| Disease Defective ACTH causes Obesity and Pro-opiomelanocortinin deficiency (POMCD) \| Disease Glucagon-type ligand receptors \| Signal transduction Class B/2 (Secretin family receptors) \| Signal transduction Loss of Function of TGFBR1 in Cancer \| Disease Hydroxycarboxylic acid-binding receptors \| Signal transduction Leishmania infection \| Disease |
| Sepsis | fungal meningitis respiratory syncytial virus infectious disease tinea unguium bacterial sepsis | SLC6A15 SLC6A20 SLC6A18 SLC6A5 SLC6A19 SLC6A11 SLC6A16 SLC6A6 SLC6A9 SLC6A12 SLC6A13 SLC6A3 SLC6A1 SLC6A14 SLC6A2 SLC6A4 SLC6A7 CARTPT SLC6A17 KDM1B SLC16A13 YARS2 SMOX SIGMAR1 IL4I1 TMEM64 YARS SLC18A1 NGF WARS2 PPOX MAOA SLC18A2 BDNF PAOX MAOB PIK3R1 HTR1A NCOA5 RXFP1 NPY2R MFSD11 HRH4 DRD5 EPOR LGR6 TNNT2 TNNI1 F2R KCNMB3 GPR25 HTR4 GPR119 SH2D1B ADRB2 RXFP4 LCE6A OR7A17 GPR149 HRH2 AVP ADRA2B FZD7 ADRA1D CCR9 CXCR2 TACR1 HRH3 ACKR2 VIPR1 SUSD5 GPR55 TNNT1 ACKR3 GIPR GPR34 OR2T8 OR2T4 OR2T6 SUCNR1 GPR4 DRD3 GPR128 NTF4 OR2T5 ABHD1 SLC30A6 FSHR ADRA1B CCKBR DRD2 TH TNNT3 TNNI2 PRLHR OR52I2 ADRB1 NTF3 TAS2R14 GPR19 | Na+/Cl- dependent neurotransmitter transporters \| Transport of small molecules GPCR ligand binding \| Signal transduction Class A/1 (Rhodopsin-like receptors) \| Signal transduction Amine ligand-binding receptors \| Signal transduction "Transport of bile salts and organic acids \| metal ions and amine compounds" ADORA2B mediated anti-inflammatory cytokines production \| Disease Signaling by GPCR \| Signal transduction GPCR downstream signalling \| Signal transduction G alpha (s) signalling events \| Signal transduction Reuptake of GABA \| Neuronal system Histamine receptors \| Signal transduction SLC-mediated transmembrane transport \| Transport of small molecules Peptide ligand-binding receptors \| Signal transduction Amine Oxidase reactions \| Metabolism Striated Muscle Contraction \| Muscle contraction Dopamine receptors \| Signal transduction Amino acid transport across the plasma membrane \| Transport of small molecules Anti-inflammatory response favouring Leishmania parasite infection \| Disease Leishmania parasite growth and survival \| Disease Adrenoceptors \| Signal transduction SLC transporter disorders \| Disease Chemokine receptors bind chemokines \| Signal transduction Creatine metabolism \| Metabolism Relaxin receptors \| Signal transduction Biogenic amines are oxidatively deaminated to aldehydes by MAOA and MAOB \| Metabolism PAOs oxidise polyamines to amines \| Metabolism Transport of inorganic cations/anions and amino acids/oligopeptides \| Transport of small molecules Serotonin receptors \| Signal transduction Disorders of transmembrane transporters \| Disease Serotonin clearance from the synaptic cleft \| Neuronal system Leishmania infection \| Disease MECP2 regulates transcription of neuronal ligands \| Gene expression (Transcription) Defective SLC6A18 may confer susceptibility to iminoglycinuria and/or hyperglycinuria \| Disease Defective SLC6A18 may confer susceptibility to iminoglycinuria and/or hyperglycinuria \| Disease "Defective AVP does not bind AVPR1A \| B and causes neurohypophyseal diabetes insipidus (NDI)" Defective SLC6A3 causes Parkinsonism-dystonia infantile (PKDYS) \| Disease Defective SLC6A3 causes Parkinsonism-dystonia infantile (PKDYS) \| Disease G alpha (q) signalling events \| Signal transduction Defective SLC6A2 causes orthostatic intolerance (OI) \| Disease Defective SLC6A5 causes hyperekplexia 3 (HKPX3) \| Disease Activated NTRK2 signals through PI3K \| Signal transduction Neurotransmitter release cycle \| Neuronal system Defective AVP does not bind AVPR2 and causes neurohypophyseal diabetes insipidus (NDI) \| Disease Defective MAOA causes Brunner syndrome (BRUNS) \| Disease Adrenaline signalling through Alpha-2 adrenergic receptor \| Hemostasis Loss of MECP2 binding ability to 5mC-DNA \| Disease Activated NTRK2 signals through PLCG1 \| Signal transduction NTF4 activates NTRK2 (TRKB) signaling \| Signal transduction NTF3 activates NTRK3 signaling \| Signal transduction Transfer of LPS from LBP carrier to CD14 \| Immune system UNC93B1 deficiency - HSE \| Disease Activated NTRK2 signals through RAS \| Signal transduction Glucagon-type ligand receptors \| Signal transduction NTF3 activates NTRK2 (TRKB) signaling \| Signal transduction BDNF activates NTRK2 (TRKB) signaling \| Signal transduction SMAC(DIABLO)-mediated dissociation of IAP:caspase complexes \| Programmed cell death Transport of small molecules \| Transport of small molecules Neurotransmitter clearance \| Neuronal system |
| Bacteremia | alexia sexual dysfunction papilloma disease hyperglycemia chronic fatigue syndrome | CCR9 CCR6 HCRTR2 NMBR HOMER3 KDM1B ACKR3 IYD CNR2 KCNK3 ABHD1 LPAR5 PAOX LTB4R SLC18A3 PPOX MC1R FAM57A SMOX CLDN6 RPTOR AVP MAOA MAOB YARS OR2T6 ADRB3 GPR55 TAAR5 GPR65 TAAR1 GPR4 ACKR2 OXER1 IL4I1 OR4A5 LCE6A GLP1R KCNK9 HOMER1 SLC18A1 MRPL23 SLC6A6 YARS2 RXFP1 SLC6A19 OR2A7 PIK3R3 SLC6A20 ADRA1A WARS2 HTR7 SLC6A9 GPR34 GPR128 GPR132 OR8U9 OR2AT4 SLC6A2 LOC100996758 DRD3 SLC6A11 SLC6A16 OR2A1 DRD2 SLC6A13 SLC6A12 TACR2 SLC6A14 PRLHR SLC18A2 OR6B2 CARTPT SLC6A1 DRD4 ADRA2A FFAR1 SLC6A3 SLC6A18 F2R SLC6A17 OR7E24 SLC6A4 OPRM1 SLC6A15 HRH2 SLC6A5 LGR5 PTGDR2 SLC6A7 LHCGR HRH4 HTR4 HTR6 FSHR TAAR6 OR5M1 RXFP2 HRH3 TMEM64 | Na+/Cl- dependent neurotransmitter transporters \| Transport of small molecules Class A/1 (Rhodopsin-like receptors) \| Signal transduction GPCR ligand binding \| Signal transduction Signaling by GPCR \| Signal transduction GPCR downstream signalling \| Signal transduction Amine ligand-binding receptors \| Signal transduction G alpha (s) signalling events \| Signal transduction ADORA2B mediated anti-inflammatory cytokines production \| Disease "Transport of bile salts and organic acids \| metal ions and amine compounds" Leishmania parasite growth and survival \| Disease Anti-inflammatory response favouring Leishmania parasite infection \| Disease Peptide ligand-binding receptors \| Signal transduction Reuptake of GABA \| Neuronal system G alpha (q) signalling events \| Signal transduction Histamine receptors \| Signal transduction Amine Oxidase reactions \| Metabolism SLC-mediated transmembrane transport \| Transport of small molecules Dopamine receptors \| Signal transduction Amino acid transport across the plasma membrane \| Transport of small molecules Serotonin receptors \| Signal transduction SLC transporter disorders \| Disease TWIK-releated acid-sensitive K+ channel (TASK) \| Neuronal system Leishmania infection \| Disease Chemokine receptors bind chemokines \| Signal transduction Olfactory Signaling Pathway \| Signal transduction Creatine metabolism \| Metabolism Adrenoceptors \| Signal transduction Relaxin receptors \| Signal transduction Biogenic amines are oxidatively deaminated to aldehydes by MAOA and MAOB \| Metabolism PAOs oxidise polyamines to amines \| Metabolism Tandem pore domain potassium channels \| Neuronal system Transport of inorganic cations/anions and amino acids/oligopeptides \| Transport of small molecules Orexin and neuropeptides FF and QRFP bind to their respective receptors \| Signal transduction Disorders of transmembrane transporters \| Disease Serotonin clearance from the synaptic cleft \| Neuronal system Hormone ligand-binding receptors \| Signal transduction Neurotransmitter release cycle \| Neuronal system Phase 4 - resting membrane potential \| Muscle contraction Defective SLC6A18 may confer susceptibility to iminoglycinuria and/or hyperglycinuria \| Disease Defective SLC6A18 may confer susceptibility to iminoglycinuria and/or hyperglycinuria \| Disease Eicosanoid ligand-binding receptors \| Signal transduction G alpha (i) signalling events \| Signal transduction "Defective AVP does not bind AVPR1A \| B and causes neurohypophyseal diabetes insipidus (NDI)" Defective SLC6A3 causes Parkinsonism-dystonia infantile (PKDYS) \| Disease Defective SLC6A3 causes Parkinsonism-dystonia infantile (PKDYS) \| Disease Defective SLC6A2 causes orthostatic intolerance (OI) \| Disease Defective SLC6A5 causes hyperekplexia 3 (HKPX3) \| Disease Defective AVP does not bind AVPR2 and causes neurohypophyseal diabetes insipidus (NDI) \| Disease Defective MAOA causes Brunner syndrome (BRUNS) \| Disease Adrenaline signalling through Alpha-2 adrenergic receptor \| Hemostasis Neurotransmitter clearance \| Neuronal system Transport of small molecules \| Transport of small molecules |
| Dermatologic complications/pressure ulcer | Ehlers-Danlos syndrome carbuncle capillary hemangioma skin hemangioma dermatographia psoriasis hemorrhoid | LGR6 OR52D1 OR52N2 PTGDR2 TAS2R41 SUCNR1 APLNR AGTR1 OR5M10 CXCR6 OR2T6 OR5V1 GALR1 OR5M1 OR51B6 TAS2R14 PRLHR GPR25 F2RL2 GPR148 F2R RGR NPY2R OR8S1 GPR151 OR52A5 CLDN6 GPR146 GPR88 SLC6A15 SCN8A COX7A2 ELOVL4 GPR19 TAS2R31 GLP1R NUP107 LGR5 OPN3 SLC8A3 RXFP2 HTR4 GPR150 SLC6A2 NTSR2 APH1B ADRB2 SLC6A7 TMEM62 SLC6A17 OR12D2 DRD1 KCNK2 GPR6 SLC6A13 OR10A5 CCKBR OR56A1 OR56A4 OR52J3 OR51L1 OR51A2 OR51A4 NR4A3 OR52I2 OR52B4 GPR21 SDCCAG3 SLC18A2 ADRB1 ELOVL3 NPFFR1 TACR2 OR10A2 SLC6A5 BDNF APH1A TAAR9 SLC6A12 TAAR6 NGF TAAR1 TSPAN13 ELOVL7 OR5A1 OR5AK2 OR5M11 SLC18A1 OR8U1 FCAMR ADRB3 NPBWR1 OR13A1 GPR149 GHSR IL9R XCR1 GPR78 KISS1R GRP | G alpha (s) signalling events \| Signal transduction Olfactory Signaling Pathway \| Signal transduction Signaling by GPCR \| Signal transduction GPCR downstream signalling \| Signal transduction GPCR ligand binding \| Signal transduction Class A/1 (Rhodopsin-like receptors) \| Signal transduction Peptide ligand-binding receptors \| Signal transduction ADORA2B mediated anti-inflammatory cytokines production \| Disease Na+/Cl- dependent neurotransmitter transporters \| Transport of small molecules Amine ligand-binding receptors \| Signal transduction Anti-inflammatory response favouring Leishmania parasite infection \| Disease Leishmania parasite growth and survival \| Disease Signal Transduction \| Signal transduction Opsins \| Signal transduction Reuptake of GABA \| Neuronal system G alpha (q) signalling events \| Signal transduction NOTCH4 Activation and Transmission of Signal to the Nucleus \| Signal transduction Synthesis of very long-chain fatty acyl-CoAs \| Metabolism Leishmania infection \| Disease MECP2 regulates transcription of neuronal ligands \| Gene expression (Transcription) Adrenoceptors \| Signal transduction "Transport of bile salts and organic acids \| metal ions and amine compounds" Creatine metabolism \| Metabolism NOTCH2 Activation and Transmission of Signal to the Nucleus \| Signal transduction Regulation of FZD by ubiquitination \| Signal transduction Class C/3 (Metabotropic glutamate/pheromone receptors) \| Signal transduction Defective SLC6A2 causes orthostatic intolerance (OI) \| Disease Defective SLC6A5 causes hyperekplexia 3 (HKPX3) \| Disease TWIK related potassium channel (TREK) \| Neuronal system Loss of MECP2 binding ability to 5mC-DNA \| Disease NRIF signals cell death from the nucleus \| Signal transduction Transfer of LPS from LBP carrier to CD14 \| Immune system Fatty acyl-CoA biosynthesis \| Metabolism BDNF activates NTRK2 (TRKB) signaling \| Signal transduction G alpha (i) signalling events \| Signal transduction |
| Ocular symptoms | hypersecretion glaucoma aqueous misdirection ptosis ocular hyperemia retinopathy of prematurity low tension glaucoma interval angle-closure glaucoma acute closed-angle glaucoma residual stage angle-closure glaucoma primary angle-closure glaucoma Marfan syndrome chronic closed-angle glaucoma glaucoma optic nerve disease central nervous system origin vertigo auditory system disease retinal disease retinal degeneration ocular hypertension borderline glaucoma mechanical strabismus hypertropia eye accommodation disease suppression amblyopia amblyopia residual stage of open angle glaucoma open-angle glaucoma primary open angle glaucoma | GPR50 OR52I2 CRHR1 AGTR1 PTGDR2 XCR1 F2RL2 OR10A7 OR5V1 OR12D2 GPR25 SUCNR1 OR51L1 GLP1R OR5M11 GPR146 OR5AK2 APLNR GPBAR1 OR51B6 OR51A2 CNR2 NPFFR1 ADRB2 RXFP2 RXFP1 OR51A4 CLDN6 GPR21 OR13A1 OR52B4 GIPR ADRB3 OR8U9 GPR148 TACR1 MTNR1B GALR1 OR8S1 MC1R TAS2R41 NPY2R AVPR2 RRH GALR3 BRS3 ADRB1 OR10A5 GPR88 OR56A1 OR52D1 GPR149 NPBWR2 OR8U1 PRLHR GPR78 OR5M8 GPR6 OR5M1 RGR CCKBR ADORA2A TAS2R14 OR52J3 F2R TACR2 ADRA2B OR2T6 GPR119 GPR19 GPR151 KCNA10 KISS1R CXCR6 TAAR9 LGR6 TAAR6 OR5M10 MC5R TAAR5 DRD1 OR1J1 CCKAR TACR3 CDKN2B DRD5 GALR2 HTR4 HTR6 OR52N2 MAGEB2 PPAN-P2RY11 TMEM86B TAAR1 NPBWR1 GPR150 VDR PPP2R1B OR52A5 GNRHR | G alpha (s) signalling events \| Signal transduction Olfactory Signaling Pathway \| Signal transduction Peptide ligand-binding receptors \| Signal transduction Class A/1 (Rhodopsin-like receptors) \| Signal transduction GPCR ligand binding \| Signal transduction ADORA2B mediated anti-inflammatory cytokines production \| Disease Signaling by GPCR \| Signal transduction GPCR downstream signalling \| Signal transduction Leishmania parasite growth and survival \| Disease Anti-inflammatory response favouring Leishmania parasite infection \| Disease Signal Transduction \| Signal transduction Amine ligand-binding receptors \| Signal transduction Leishmania infection \| Disease Tachykinin receptors bind tachykinins \| Signal transduction G alpha (i) signalling events \| Signal transduction G alpha (q) signalling events \| Signal transduction Adrenoceptors \| Signal transduction Relaxin receptors \| Signal transduction Opsins \| Signal transduction Dopamine receptors \| Signal transduction Serotonin receptors \| Signal transduction Cargo recognition for clathrin-mediated endocytosis \| Vesicle-mediated transport Cyclin D associated events in G1 \| Cell cycle G1 Phase \| Cell cycle Defective AVP does not bind AVPR2 and causes neurohypophyseal diabetes insipidus (NDI) \| Disease Adrenaline signalling through Alpha-2 adrenergic receptor \| Hemostasis Adenosine P1 receptors \| Signal transduction Glucagon-type ligand receptors \| Signal transduction Class B/2 (Secretin family receptors) \| Signal transduction Senescence-Associated Secretory Phenotype (SASP) \| Cellular responses to external stimuli PP2A-mediated dephosphorylation of key metabolic factors \| Metabolism Blockage of phagosome acidification \| Disease |

**Table S3.** Hierarchically ranked comorbidities, comorbidity enriched MOA proteins, and the pathways for each uncharacterized COVID-19 manifestation using the SARS-CoV-2 interactome(1) as input.

| **Uncharacterized manifestation** | **Comorbidities** | **Comorbidity enriched MOA Proteins** | **Pathways \| Top Pathways** |
| --- | --- | --- | --- |
| Neoplasms | alveolar soft part sarcoma T-cell lymphoblastic leukemia/lymphoma acute monocytic leukemia prolymphocytic leukemia chronic myeloid leukemia fibrosarcoma adenosquamous carcinoma synovial sarcoma thrombocytosis acinar cell carcinoma fallopian tube carcinoma papillary adenocarcinoma malignant mesothelioma lymphoma leukemia reticulosarcoma ovarian carcinoma skin cancer obsolete lymphosarcoma Ewing sarcoma meningioma obsolete lymphoid leukemia hemangiopericytoma Leydig cell tumor megakaryocytic leukemia Hodgkin's lymphomai, mixed cellularity pleomorphic liposarcoma liposarcoma Klatskin's tumor osteosarcoma chondrosarcoma mesenchymoma chronic lymphocytic leukemia hairy cell leukemia Langerhans-cell histiocytosis essential thrombocythemia polycythemia vera Hodgkin's granuloma Hodgkin's lymphoma, lymphocytic depletion Hodgkin's lymphoma Hodgkin's lymphoma, lymphocytic-histiocytic predominance Burkitt lymphoma plasmacytoma skin benign neoplasm obsolete chronic erythremia subacute monocytic leukemia intrapelvic lymph node leukemic reticuloendotheliosis intra-abdominal lymph node mast cell malignancy subacute leukemia splenic manifestation of hairy cell leukemia lymphoplasmacytic lymphoma macroglobulinemia neuroendocrine tumor pancreatic carcinoma thyroid adenoma thyroid gland carcinoma urinary system benign neoplasm myeloid sarcoma rectal benign neoplasm embryonal rhabdomyosarcoma spindle cell sarcoma epithelioid sarcoma pancreatic adenocarcinoma benign giant cell tumor myelodysplastic syndrome Sezary's disease multiple myeloma lymphoid leukemia mycosis fungoides mast-cell leukemia myeloid leukemia melanoma myelodysplastic/myeloproliferative neoplasm angiosarcoma neurofibrosarcoma chronic monocytic leukemia acute lymphoblastic leukemia acute myeloid leukemia fibrillary astrocytoma protoplasmic astrocytoma childhood cerebral astrocytoma gemistocytic astrocytoma juvenile pilocytic astrocytoma grade III astrocytoma pilocytic astrocytoma subependymal giant cell astrocytoma bone marrow cancer olfactory neuroblastoma inverted papilloma T-cell acute lymphoblastic leukemia acute promyelocytic leukemia gliosarcoma systemic mastocytosis neuroendocrine carcinoma familial retinoblastoma juvenile myelomonocytic leukemia thymic carcinoma plasma cell leukemia acute leukemia alveolar rhabdomyosarcoma medullomyoblastoma childhood medulloblastoma adult medulloblastoma laryngeal benign neoplasm pseudomyxoma peritonei angiolipoma serous cystadenocarcinoma clear cell sarcoma childhood oligodendroglioma oligodendroglioma adult oligodendroglioma mixed oligodendroglioma-astrocytoma ureteral benign neoplasm squamous cell papilloma carcinosarcoma sarcoma benign mesothelioma somatostatinoma hemangioma in situ carcinoma plexiform neurofibroma mucoepidermoid carcinoma germinoma verrucous carcinoma parasagittal meningioma intraventricular meningioma intraorbital meningioma clear cell meningioma benign meningioma secretory meningioma microcystic meningioma rhabdoid meningioma cerebral convexity meningioma angiomatous meningioma fibrous meningioma psammomatous meningioma meningothelial meningioma transitional meningioma olfactory groove meningioma spinal meningioma medulloblastoma salivary gland adenoid cystic carcinoma cystadenocarcinoma gastrinoma retinoblastoma colonic benign neoplasm hepatocellular carcinoma large cell carcinoma malignant peripheral nerve sheath tumor peripheral nerve sheath neoplasm leiomyosarcoma epithelioid leiomyosarcoma myxoid leiomyosarcoma paranasal sinus benign neoplasm monocytic leukemia transitional cell carcinoma endometrial carcinoma histiocytoid hemangioma intramuscular hemangioma chorioangioma angiomyolipoma thyroid gland cancer sarcomatoid renal cell carcinoma chromophobe renal cell carcinoma clear cell renal cell carcinoma papillary renal cell carcinoma collecting duct carcinoma squamous cell neoplasm adenoma basal cell carcinoma lung oat cell carcinoma paraganglioma neurilemmoma cancer nevoid basal cell carcinoma syndrome spindle cell carcinoma carcinoma germ cell and embryonal cancer germ cell cancer embryonal cancer nephroblastoma benign ependymoma Kaposi's sarcoma ganglioneuroblastoma pancreatic cancer follicular adenoma microcystic adenoma papillary adenoma head and neck cancer myxopapillary ependymoma papillary ependymoma cellular ependymoma granular cell carcinoma adenocarcinoma tubular adenocarcinoma cribriform carcinoma bile duct carcinoma cavernous hemangioma stomach cancer lung small cell carcinoma renal cell carcinoma cystadenoma ocular cancer mucinous cystadenocarcinoma mixed glioma malignant glioma mixed phenotype acute leukemia | PAN2 CCT6B TUBAL3 SEPT10 PACRGL TUBA4A IL7R TUBB1 IL21R SEPT11 IL4R IL9R RIOK1 TUBE1 TUBD1 ASMTL FKBP15 C2orf48 RIOK3 CAMK2G TUBG2 CAMK2A MAP2K6 MAP2K3 EEF1D PRKAA1 FKBP5 SCIN CDC20 FKBP4 GSN VPS41 FKBP11 IL2RA CORO1C FKBP14 FKBP1B AWAT1 SRPK1 FKBP3 CDC20B DCAF4L2 FKBP10 DCAF8L2 EML4 WDR89 FKBP1A WDR13 EIF2A FKBP7 TUBG1 ADCK3 WDR33 TLE1 TLE3 FKBP6 PRKAA2 METTL21A FKBP2 FKBP9 MKL1 TFB1M CAMK2B TUBA1C FGFR1 FGFR2 DNMT1 MTIF2 TUBB4Q AK2 DUT TREX2 GMPR AK4 AK7 AMPD3 METTL17 LOC100506422 METTL21B NME9 AMPD1 ERI3 ERI1 AK8 AMPD2 USB1 RAB35 ARAF RRM1 NPIPB3 POLA1 AK5 OAS2 TUT1 NTPCR PCMTD1 METTL4 CMTR1 CMPK1 GNL3 | Activation of AMPK downstream of NMDARs \| Neuronal system Signaling by FGFR2 IIIa TM \| Disease FGFR2 mutant receptor activation \| Disease Interconversion of nucleotide di- and triphosphates \| Metabolism Recruitment of NuMA to mitotic centrosomes \| Cell cycle Metabolism of nucleotides \| Metabolism of RNA Microtubule-dependent trafficking of connexons from Golgi to the plasma membrane \| Vesicle-mediated transport Transport of connexons to the plasma membrane \| Vesicle-mediated transport Carboxyterminal post-translational modifications of tubulin \| Metabolism of proteins Prefoldin mediated transfer of substrate to CCT/TriC \| Metabolism of proteins FGFR2 ligand binding and activation \| Signal transduction HSF1-dependent transactivation \| Cellular responses to external stimuli Formation of tubulin folding intermediates by CCT/TriC \| Metabolism of proteins Post-chaperonin tubulin folding pathway \| Metabolism of proteins Mitotic Prometaphase \| Cell cycle Purine salvage \| Metabolism RHO GTPases activate IQGAPs \| Signal transduction HSP90 chaperone cycle for steroid hormone receptors (SHR) \| Cellular responses to external stimuli Signaling by RAF1 mutants \| Disease Post NMDA receptor activation events \| Neuronal system Signaling by FGFR in disease \| Disease RAF activation \| Signal transduction Negative regulation of NMDA receptor-mediated neuronal transmission \| Neuronal system SHC-mediated cascade:FGFR2 \| Signal transduction Activated point mutants of FGFR2 \| Disease Cooperation of Prefoldin and TriC/CCT in actin and tubulin folding \| Metabolism of proteins PI3K Cascade \| Signal transduction Signaling downstream of RAS mutants \| Disease Signaling by moderate kinase activity BRAF mutants \| Disease Signaling by RAS mutants \| Disease Nucleotide salvage \| Metabolism Paradoxical activation of RAF signaling by kinase inactive BRAF \| Disease Signaling by BRAF and RAF fusions \| Disease Signaling by FGFR2 amplification mutants \| Disease RAF/MAP kinase cascade \| Signal transduction Chaperonin-mediated protein folding \| Metabolism of proteins "Unblocking of NMDA receptors \| glutamate binding and activation" EML4 and NUDC in mitotic spindle formation \| Cell cycle Constitutive Signaling by Aberrant PI3K in Cancer \| Disease RHO GTPases Activate Formins \| Signal transduction Phase 0 - rapid depolarisation \| Muscle contraction Signaling by FGFR1 amplification mutants \| Disease Protein folding \| Metabolism of proteins IRS-mediated signalling \| Signal transduction Phospholipase C-mediated cascade; FGFR2 \| Signal transduction IRS-related events triggered by IGF1R \| Signal transduction Assembly and cell surface presentation of NMDA receptors \| Neuronal system IGF1R signaling cascade \| Signal transduction Signaling by Type 1 Insulin-like Growth Factor 1 Receptor (IGF1R) \| Signal transduction Sealing of the nuclear envelope (NE) by ESCRT-III \| Cell cycle FGFR2b ligand binding and activation \| Signal transduction Signaling by activated point mutants of FGFR1 \| Disease MAPK1/MAPK3 signaling \| Signal transduction Signaling by FGFR2 in disease \| Disease FGFR2c ligand binding and activation \| Signal transduction Ras activation upon Ca2+ influx through NMDA receptor \| Neuronal system Oncogenic MAPK signaling \| Disease Insulin receptor signalling cascade \| Signal transduction COPI-independent Golgi-to-ER retrograde traffic \| Vesicle-mediated transport Gap junction assembly \| Vesicle-mediated transport Resolution of Sister Chromatid Cohesion \| Cell cycle Aggrephagy \| Autophagy activated TAK1 mediates p38 MAPK activation \| Immune system FGFR1 ligand binding and activation \| Signal transduction Recruitment of mitotic centrosome proteins and complexes \| Cell cycle Separation of Sister Chromatids \| Cell cycle Translocation of SLC2A4 (GLUT4) to the plasma membrane \| Vesicle-mediated transport Signaling by FGFR2 fusions \| Disease TWIK-related alkaline pH activated K+ channel (TALK) \| Neuronal system Signaling by Insulin receptor \| Signal transduction AURKA Activation by TPX2 \| Cell cycle "PI5P \| PP2A and IER3 Regulate PI3K/AKT Signaling" COPI-dependent Golgi-to-ER retrograde traffic \| Vesicle-mediated transport COPI-mediated anterograde transport \| Metabolism of proteins CaMK IV-mediated phosphorylation of CREB \| Signal transduction Gap junction trafficking \| Vesicle-mediated transport Activation of NMDA receptors and postsynaptic events \| Neuronal system Recycling pathway of L1 \| Developmental biology TGFBR1 LBD Mutants in Cancer \| Disease Centrosome maturation \| Cell cycle CREB1 phosphorylation through NMDA receptor-mediated activation of RAS signaling \| Neuronal system Gap junction trafficking and regulation \| Vesicle-mediated transport Kinesins \| Hemostasis Negative regulation of the PI3K/AKT network \| Signal transduction Ion homeostasis \| Muscle contraction Signaling by plasma membrane FGFR1 fusions \| Disease Cardiac conduction \| Muscle contraction Association of TriC/CCT with target proteins during biosynthesis \| Metabolism of proteins Ion transport by P-type ATPases \| Transport of small molecules Long-term potentiation \| Neuronal system |
| Congenital malformations | osteopetrosis Down syndrome Crouzon syndrome tuberous sclerosis Cowden syndrome Klippel-Trenaunay syndrome Peutz-Jeghers syndrome pachyonychia congenita neurofibromatosis capillary hemangioma placental insufficiency pulmonary valve stenosis McCune Albright syndrome | CCT6B PAN2 FKBP6 VPS41 STAC2 GSN IL21R FKBP1B SCIN IL4R FKBP11 FKBP2 AIP IL9R WRAP53 FKBP9 DGKE IL2RA EML4 FKBP14 TLE1 ARHGEF28 FKBP15 FKBP7 TMEM81 PACRGL FKBP4 DGKB RPTOR C2orf48 IL7R IARS2 WDR33 FKBP10 AGPAT9 CORO1C EIF2A DCAF8L2 CDC20B WDR13 FKBP1A WDR89 FKBP3 PHTF2 NEXN AWAT1 DCAF4L2 CDC20 WDR38 TENC1 FKBP5 EEF1D TLE3 CILP MKL1 RASGRP2 PPP3R2 MRPL32 CD248 PPP3CB PGLYRP4 TOMM5 UBP1 IL2RB NAGPA NOMO2 PLXDC2 PPP3CC PGLYRP3 CLSTN3 ARHGEF2 WDR5 MRPL4 WDR83 WDR26 STRN4 PGLYRP2 ERCC8 ITGB1 RBBP5 FGF21 GRWD1 PGLYRP1 FGF7 RFWD2 WDR47 GNB1L SMU1 TBC1D31 FGF18 WDR77 THOC3 TBL1X PPP2R2B RBBP7 GNB2L1 PPP2R2A FGF17 FGF20 IL2RG | FGFR2 ligand binding and activation \| Signal transduction Activated point mutants of FGFR2 \| Disease FGFR3b ligand binding and activation \| Signal transduction FGFR2c ligand binding and activation \| Signal transduction FGFR4 ligand binding and activation \| Signal transduction Signaling by activated point mutants of FGFR3 \| Disease FGFR3c ligand binding and activation \| Signal transduction FGFR3 ligand binding and activation \| Signal transduction FGFR2 mutant receptor activation \| Disease SHC-mediated cascade:FGFR2 \| Signal transduction FGFR3 mutant receptor activation \| Disease Calcineurin activates NFAT \| Immune system Interleukin receptor SHC signaling \| Immune system Neddylation \| Metabolism of proteins Association of TriC/CCT with target proteins during biosynthesis \| Metabolism of proteins Interleukin-21 signaling \| Immune system Phospholipase C-mediated cascade; FGFR2 \| Signal transduction Effects of PIP2 hydrolysis \| Hemostasis FGFRL1 modulation of FGFR1 signaling \| Signal transduction Signaling by activated point mutants of FGFR1 \| Disease SHC-mediated cascade:FGFR3 \| Signal transduction SHC-mediated cascade:FGFR4 \| Signal transduction HSF1-dependent transactivation \| Cellular responses to external stimuli FGFR1c ligand binding and activation \| Signal transduction CLEC7A (Dectin-1) induces NFAT activation \| Immune system Mitochondrial translation elongation \| Metabolism of proteins Mitochondrial translation termination \| Metabolism of proteins FGFR1 ligand binding and activation \| Signal transduction Signaling by Type 1 Insulin-like Growth Factor 1 Receptor (IGF1R) \| Signal transduction Phospholipase C-mediated cascade; FGFR3 \| Signal transduction Phospholipase C-mediated cascade; FGFR4 \| Signal transduction TGFBR1 LBD Mutants in Cancer \| Disease Antimicrobial peptides \| Immune system Interleukin-2 signaling \| Immune system PI3K Cascade \| Signal transduction Mitochondrial translation initiation \| Metabolism of proteins GABA synthesis \| Neuronal system RAF/MAP kinase cascade \| Signal transduction Attenuation phase \| Cellular responses to external stimuli Loss of Function of TGFBR1 in Cancer \| Disease |

**Table S4.** Hierarchically ranked comorbidities, comorbidity enriched MOA proteins, and pathways for each uncharacterized COVID-19 manifestation using the GWAS risk genes(2) as input.

| **Uncharacterized manifestation** | **Comorbidities** | **Comorbidity enriched MOA Proteins** | **Pathways \| Top Pathways** |
| --- | --- | --- | --- |
| Neoplasms | subependymal glioma diabetic autonomic neuropathy endometrial adenocarcinoma pleomorphic lipoma histiocytoid hemangioma intramuscular hemangioma chorioangioma prolactinoma hemangioma lipoma squamous cell papilloma myofibroma prostatic adenoma uterine fibroid leiomyoma duodenal benign neoplasm bone marrow cancer pituitary carcinoma pituitary adenoma embryonal cancer germ cell and embryonal cancer germ cell cancer cerebral primitive neuroectodermal tumor ependymoblastoma medulloepithelioma neuroectodermal tumor head and neck cancer | NUP107 APH1A COX7A2 COX7A2L SLC8A2 NR4A3 ELOVL3 AR SLC8A1 ELOVL7 ELOVL4 SLC8A3 APH1B COX7A1 ELOVL1 GRP CELA2B KCNA10 OR52B4 OSBPL5 TSPAN13 OR52I2 ANXA1 CSF2RA HCRTR1 TACR2 RXFP1 OSBPL8 IL22RA2 TMEM86B NR3C2 FKBP14 PRLR CCKBR PROKR2 MC1R UGGT1 NPFFR1 APLNR SUCNR1 OR13A1 CCR9 CNR2 OR2A7 NMBR RXFP2 MRGPRX1 TACR3 GHSR GALR2 GLP1R OR8U9 OR10H2 NPY2R GIPR OR12D2 PGR BRS3 LPAR5 OR51B6 GALR1 OR52N2 LTB4R MC5R OR52D1 CCT6B LHCGR SCIN OR6B2 CDC20 TACR1 IL9R GPR151 MRGPRX3 VPS41 PRLHR GPR19 AVPR2 NR3C1 KISS1R ACKR3 ICMT OR7A17 IL21R PTGDR2 IL4R IL7R MTNR1B OR2AT4 OR2A1 PAN2 CXCR2 ADRB3 XCR1 CXCR6 OR5V1 NPBWR1 LGR6 GPR149 FKBP15 | Nuclear Receptor transcription pathway \| Gene expression (Transcription) Peptide ligand-binding receptors \| Signal transduction Class A/1 (Rhodopsin-like receptors) \| Signal transduction GPCR ligand binding \| Signal transduction GPCR downstream signalling \| Signal transduction Signaling by GPCR \| Signal transduction G alpha (s) signalling events \| Signal transduction Olfactory Signaling Pathway \| Signal transduction ADORA2B mediated anti-inflammatory cytokines production \| Disease G alpha (q) signalling events \| Signal transduction Tachykinin receptors bind tachykinins \| Signal transduction Chemokine receptors bind chemokines \| Signal transduction Reduction of cytosolic Ca++ levels \| Hemostasis Synthesis of very long-chain fatty acyl-CoAs \| Metabolism Sodium/Calcium exchangers \| Transport of small molecules Relaxin receptors \| Signal transduction Orexin and neuropeptides FF and QRFP bind to their respective receptors \| Signal transduction NOTCH4 Activation and Transmission of Signal to the Nucleus \| Signal transduction Platelet calcium homeostasis \| Hemostasis G alpha (i) signalling events \| Signal transduction Anti-inflammatory response favouring Leishmania parasite infection \| Disease Leishmania parasite growth and survival \| Disease Fatty acyl-CoA biosynthesis \| Metabolism HSP90 chaperone cycle for steroid hormone receptors (SHR) \| Cellular responses to external stimuli NOTCH2 Activation and Transmission of Signal to the Nucleus \| Signal transduction Acyl chain remodelling of PS \| Metabolism Linoleic acid (LA) metabolism \| Metabolism Defective AVP does not bind AVPR2 and causes neurohypophyseal diabetes insipidus (NDI) \| Disease Glucagon-type ligand receptors \| Signal transduction |
| Behavioral | obsessive-compulsive disorder panic disorder obsessive-compulsive personality disorder dyslexia Rett syndrome substance abuse substance-related disorder mental depression autistic disorder depersonalization disorder pyromania intermittent explosive disorder kleptomania impulse control disorder neurotic disorder dementia somatoform disorder somatization disorder borderline personality disorder cannabis dependence cannabis abuse dissociative disorder conversion disorder pathological gambling post-traumatic stress disorder attention deficit hyperactivity disorder social phobia phobic disorder endogenous depression melancholia cocaine dependence cocaine abuse heroin dependence substance dependence morphine dependence hypoactive sexual desire disorder intellectual disability nicotine dependence alcohol use disorder eating disorder dysthymic disorder stereotypic movement disorder learning disability bulimia nervosa bipolar disorder mood disorder schizophreniform disorder disease of mental health schizoaffective disorder cognitive disorder conduct disorder agoraphobia gender identity disorder paranoid personality disorder expressive language disorder opioid abuse psychologic dyspareunia sexual masochism schizoid personality disorder multiple personality disorder dependent personality disorder voyeurism transvestism sexual sadism fetishism inhibited male orgasm mixed receptive-expressive language disorder exhibitionism psychologic vaginismus hallucinogen dependence developmental coordination disorder barbiturate abuse inhibited female orgasm language disorder histrionic personality disorder factitious disorder hallucinogen abuse X-linked intellectual disability-psychosis-macroorchidism syndrome premature ejaculation childhood disintegrative disease syndromic X-linked intellectual disability 94 communication disorder hypochondriasis cyclothymic disorder major depressive disorder anxiety disorder narcissistic personality disorder avoidant personality disorder opiate dependence schizophrenia vascular dementia generalized anxiety disorder alcohol dependence combat disorder schizotypal personality disorder | FSHR SLC6A3 SLC6A2 SLC6A20 SLC6A7 SLC6A14 SLC6A18 SLC6A16 SLC6A17 OR4K5 TMEM64 OR10S1 FFAR2 OR1G1 GPR39 OR52H1 OR4K1 HRH4 OR2L8 GPR31 OR1I1 GHSR NPFFR2 DRD2 FFAR4 ND1 GPR85 DRD3 TMEM116 ADRA2A EDNRB OR5AN1 DRD4 GPR50 OR2A4 CRHR1 GPR87 OR8I2 HTR1A BDKRB2 ADRA1A ADRA1B AGTR2 GPR173 CXCR5 GPR37 CHRM4 NPSR1 P2RY2 HTR7 NMUR2 GPR84 TAAR2 QRFPR HRH2 GPR161 GLP1R OPRM1 OR2T5 EDNRA ADRA1D TAAR1 C11orf49 MTNR1B HTR4 S100A4 S100A5 GPR176 LOC102723532 OR1L3 OR6Q1 FZD10 GPR52 HTR6 HTR2C NMUR1 HTR2A GPR75 OR4K14 S100A6 C1orf87 PTH2R OR5T2 OR10R2 OR2T1 OR8G2 ADORA2A GPR135 USP25 OR52K1 OR2T11 SLC6A15 TAS2R39 TAS2R60 OR4C13 MTNR1A TAS2R46 OR13G1 OR8K1 GPR61 | Amine ligand-binding receptors \| Signal transduction G alpha (s) signalling events \| Signal transduction Class A/1 (Rhodopsin-like receptors) \| Signal transduction GPCR ligand binding \| Signal transduction Signaling by GPCR \| Signal transduction GPCR downstream signalling \| Signal transduction Olfactory Signaling Pathway \| Signal transduction ADORA2B mediated anti-inflammatory cytokines production \| Disease Serotonin receptors \| Signal transduction G alpha (q) signalling events \| Signal transduction Anti-inflammatory response favouring Leishmania parasite infection \| Disease Leishmania parasite growth and survival \| Disease Na+/Cl- dependent neurotransmitter transporters \| Transport of small molecules Peptide ligand-binding receptors \| Signal transduction Signal Transduction \| Signal transduction Dopamine receptors \| Signal transduction G alpha (i) signalling events \| Signal transduction Leishmania infection \| Disease Histamine receptors \| Signal transduction Adrenoceptors \| Signal transduction Orexin and neuropeptides FF and QRFP bind to their respective receptors \| Signal transduction Class B/2 (Secretin family receptors) \| Signal transduction Defective SLC6A18 may confer susceptibility to iminoglycinuria and/or hyperglycinuria \| Disease Defective SLC6A18 may confer susceptibility to iminoglycinuria and/or hyperglycinuria \| Disease SLC transporter disorders \| Disease Defective SLC6A3 causes Parkinsonism-dystonia infantile (PKDYS) \| Disease Defective SLC6A3 causes Parkinsonism-dystonia infantile (PKDYS) \| Disease Amino acid transport across the plasma membrane \| Transport of small molecules Surfactant metabolism \| Metabolism of proteins Class C/3 (Metabotropic glutamate/pheromone receptors) \| Signal transduction Defective SLC6A2 causes orthostatic intolerance (OI) \| Disease Adrenaline signalling through Alpha-2 adrenergic receptor \| Hemostasis Adenosine P1 receptors \| Signal transduction Muscarinic acetylcholine receptors \| Signal transduction "Transport of bile salts and organic acids \| metal ions and amine compounds" Nucleotide-like (purinergic) receptors \| Signal transduction Defective GIF causes intrinsic factor deficiency \| Disease |

**Table S5.** Predicted neoplasm comorbidity enriched MOA proteins that are labeled as cancer associated in the COSMIC(30) database using the SARS-CoV-2 interactome (1)as input.

| **Gene name** | **Gene description** |
| --- | --- |
| IL7R | interleukin 7 receptor |
| IL21R | interleukin 21 receptor |
| EML4 | echinoderm microtubule associated protein like 4 |
| FKBP9 | FK506 binding protein 9 |
| FGFR1 | fibroblast growth factor receptor 1 |
| FGFR2 | fibroblast growth factor receptor 2 |
| ARAF | A-Raf proto-oncogene, serine/threonine kinase |
| HRAS | v-Ha-ras Harvey rat sarcoma viral oncogene homolog |
| KRAS | v-Ki-ras2 Kirsten rat sarcoma 2 viral oncogene homolog |
| 6-Sep | septin 6 |
| ERG | v-ets erythroblastosis virus E26 oncogene like (avian) |
| 9-Sep | septin 9 |
| NRAS | neuroblastoma RAS viral (v-ras) oncogene homolog |
| 5-Sep | septin 5 |
| PRKAR1A | protein kinase, cAMP-dependent, regulatory, type I, alpha (tissue specific extinguisher 1) |
| ACSL6 | acyl-CoA synthetase long-chain family member 6 |
| ARHGAP5 | Rho GTPase activating protein 5 |
| RAC1 | ras-related C3 botulinum toxin substrate 1 (rho family, small GTP binding protein Rac1) |
| CHD2 | chromodomain helicase DNA binding protein 2 |
| RHOA | ras homolog family member A |
| SETDB1 | SET domain bifurcated 1 |
| RHOH | ras homolog family member H |
| FHIT | fragile histidine triad gene |
| FGFR3 | fibroblast growth factor receptor 3 |
| GNAS | guanine nucleotide binding protein (G protein), alpha stimulating activity polypeptide 1 |
| GNAQ | guanine nucleotide binding protein (G protein), q polypeptide |
| POLD1 | DNA polymerase delta 1, catalytic subunit |
| GNA11 | guanine nucleotide binding protein (G protein), alpha 11 (Gq class) |
| MAP2K4 | mitogen-activated protein kinase kinase 4 |
| WRN | Werner syndrome (RECQL2) |
| PDGFRA | platelet-derived growth factor, alpha-receptor |
| PDGFRB | platelet-derived growth factor receptor, beta polypeptide |
| ELN | elastin |
| ATIC | 5-aminoimidazole-4-carboxamide ribonucleotide formyltransferase/IMP cyclohydrolase |
| POT1 | protection of telomeres 1 |
| CIITA | class II, major histocompatibility complex, transactivator |
| RAD17 | RAD17 checkpoint clamp loader component |
| FGFR4 | fibroblast growth factor receptor 4 |
| PMS2 | PMS2 postmeiotic segregation increased 2 (S. cerevisiae) |
| ACSL3 | acyl-CoA synthetase long-chain family member 3 |
| BCL10 | B-cell CLL/lymphoma 10 |
| MLLT11 | myeloid/lymphoid or mixed-lineage leukemia (trithorax homolog, Drosophila); translocated to, 11 |
| ERCC3 | excision repair cross-complementing rodent repair deficiency, complementation group 3 (xeroderma pigmentosum group B complementing) |
| EZH2 | enhancer of zeste homolog 2 |
| ERBB2 | v-erb-b2 erythroblastic leukemia viral oncogene homolog 2, neuro/glioblastoma derived oncogene homolog (avian) |
| CDK12 | cyclin-dependent kinase 12 |
| ERBB3 | erb-b2 receptor tyrosine kinase 3 |
| EGFR | epidermal growth factor receptor (erythroblastic leukemia viral (v-erb-b) oncogene homolog, avian) |
| DICER1 | dicer 1, ribonuclease type III |
| RAD51B | RAD51 paralog B |
| MAP3K1 | mitogen-activated protein kinase kinase kinase 1, E3 ubiquitin protein ligase |
| PIM1 | pim-1 oncogene |
| KIF5B | kinesin family member 5B |
| KNSTRN | kinetochore localized astrin/SPAG5 binding protein |
| MSH6 | mutS homolog 6 (E. coli) |
| DDX5 | DEAD (Asp-Glu-Ala-Asp) box polypeptide 5 |
| DDX10 | DEAD (Asp-Glu-Ala-Asp) box polypeptide 10 |
| BTK | Bruton agammaglobulinemia tyrosine kinase |
| FES | FES proto-oncogene, tyrosine kinase |
| FLT3 | fms-related tyrosine kinase 3 |
| KDR | vascular endothelial growth factor receptor 2 |
| MET | met proto-oncogene (hepatocyte growth factor receptor) |
| RET | ret proto-oncogene |
| BRAF | v-raf murine sarcoma viral oncogene homolog B1 |
| FLT4 | fms-related tyrosine kinase 4 |
| NTRK3 | neurotrophic tyrosine kinase, receptor, type 3 |
| ALK | anaplastic lymphoma kinase (Ki-1) |
| NTRK1 | neurotrophic tyrosine kinase, receptor, type 1 |
| KIT | v-kit Hardy-Zuckerman 4 feline sarcoma viral oncogene homolog |
| EIF4A2 | eukaryotic translation initiation factor 4A, isoform 2 |
| CSF1R | colony stimulating factor 1 receptor |
| MSH2 | mutS homolog 2 (E. coli) |
| CNBD1 | cyclic nucleotide binding domain containing 1 |
| JAK1 | Janus kinase 1 |
| JAK2 | Janus kinase 2 |
| DDR2 | discoidin domain receptor 2 |
| JAK3 | Janus kinase 3 |
| CDK4 | cyclin-dependent kinase 4 |
| ABL1 | v-abl Abelson murine leukemia viral oncogene homolog 1 |
| CDK6 | cyclin-dependent kinase 6 |
| ERBB4 | erb-b2 receptor tyrosine kinase 4 |
| TEC | tec protein tyrosine kinase |
| DNM2 | dynamin 2 |
| KDSR | 3-ketodihydrosphingosine reductase |
| TGFBR2 | transforming growth factor beta receptor II |
| KAT7 | lysine acetyltransferase 7 |
| MAP3K13 | mitogen-activated protein kinase kinase kinase 13 |
| BLM | Bloom Syndrome |
| ALDH2 | aldehyde dehydrogenase 2 family (mitochondrial) |
| DDX3X | DEAD-box helicase 3, X-linked |
| ACVR2A | activin A receptor type 2A |
| ACVR1 | activin A receptor, type I |
| CARS | cysteinyl-tRNA synthetase |
| RECQL4 | RecQ protein-like 4 |
| SMC1A | structural maintenance of chromosomes 1A |
| CREB1 | cAMP responsive element binding protein 1 |
| AR | Androgen Receptor |
| MAP2K2 | mitogen-activated protein kinase kinase 2 |
| MAP2K1 | mitogen-activated protein kinase kinase 1 |
| ABL2 | c-abl oncogene 2, non-receptor tyrosine kinase |
| PTK6 | protein tyrosine kinase 6 |
| SYK | spleen tyrosine kinase |
| EPS15 | epidermal growth factor receptor pathway substrate 15 (AF1p) |
| NR4A3 | nuclear receptor subfamily 4, group A, member 3 (NOR1) |
| EPHA7 | EPH receptor A7 |
| CHEK2 | CHK2 checkpoint homolog (S. pombe) |
| ITK | IL2-inducible T-cell kinase |
| EPHA3 | EPH receptor A3 |
| BMPR1A | bone morphogenetic protein receptor, type IA |
| BUB1B | BUB1 budding uninhibited by benzimidazoles 1 homolog beta (yeast) |
| STK11 | serine/threonine kinase 11 gene (LKB1) |
| SRC | SRC proto-oncogene, non-receptor tyrosine kinase |
| RAF1 | v-raf-1 murine leukemia viral oncogene homolog 1 |
| PRDM1 | PR domain containing 1, with ZNF domain |
| LCK | lymphocyte-specific protein tyrosine kinase |
| PRKACA | protein kinase cAMP-activated catalytic subunit alpha |
| DDX6 | DEAD (Asp-Glu-Ala-Asp) box polypeptide 6 |
| ANK1 | ankyrin 1 |
| BCL2 | B-cell CLL/lymphoma 2 |
| SGK1 | serum/glucocorticoid regulated kinase 1 |
| PHF6 | PHD finger protein 6 |
| COL3A1 | collagen type III alpha 1 chain |
| LATS1 | large tumor suppressor kinase 1 |
| EXT2 | multiple exostoses type 2 gene |
| AKT2 | v-akt murine thymoma viral oncogene homolog 2 |
| LATS2 | large tumor suppressor kinase 2 |
| P2RY8 | purinergic receptor P2Y, G-protein coupled, 8 |
| MYH9 | myosin, heavy polypeptide 9, non-muscle |
| SF3B1 | splicing factor 3b, subunit 1, 155kDa |
| NF1 | neurofibromatosis type 1 gene |
| CHST11 | carbohydrate sulfotransferase 11 |
| MYO5A | myosin VA (heavy chain 12, myoxin) |
| AKT3 | v-akt murine thymoma viral oncogene homolog 3 |
| N4BP2 | NEDD4 binding protein 2 |
| MLH1 | E.coli MutL homolog gene |
| MYH11 | myosin, heavy polypeptide 11, smooth muscle |
| CLP1 | cleavage and polyadenylation factor I subunit 1 |
| ZEB1 | zinc finger E-box binding homeobox 1 |
| MYB | v-myb myeloblastosis viral oncogene homolog |
| PPP2R1A | protein phosphatase 2, regulatory subunit A, alpha |
| PPARG | peroxisome proliferative activated receptor, gamma |
| MUC1 | mucin 1, transmembrane |
| BCR | breakpoint cluster region |
| CAMTA1 | calmodulin binding transcription activator 1 |
| ELK4 | ELK4, ETS-domain protein (SRF accessory protein 1) |
| NFIB | nuclear factor I/B |
| FIP1L1 | FIP1 like 1 (S. cerevisiae) |
| GMPS | guanine monphosphate synthetase |
| CTNNA2 | catenin alpha 2 |
| PPP6C | protein phosphatase 6, catalytic subunit |
| LMO2 | LIM domain only 2 (rhombotin-like 1) (RBTN2) |
| PRDM2 | PR/SET domain 2 |
| HOOK3 | hook homolog 3 |
| ETV4 | ets variant gene 4 (E1A enhancer binding protein, E1AF) |
| GRIN2A | glutamate receptor, ionotropic, N-methyl D-aspartate 2A |
| DNMT3A | DNA (cytosine-5-)-methyltransferase 3 alpha |
| CALR | calreticulin |
| BRIP1 | BRCA1 interacting protein C-terminal helicase 1 |
| NRG1 | neuregulin 1 |
| IKBKB | inhibitor of kappa light polypeptide gene enhancer in B-cells, kinase beta |
| SMARCB1 | SWI/SNF related, matrix associated, actin dependent regulator of chromatin, subfamily b, member 1 |
| FOXA1 | forkhead box A1 |
| TSHR | thyroid stimulating hormone receptor |
| CHCHD7 | coiled-coil-helix-coiled-coil-helix domain containing 7 |
| IDH1 | isocitrate dehydrogenase 1 (NADP+), soluble |
| IDH2 | isocitrate dehydrogenase 2 (NADP+), mitochondrial |
| NUP98 | nucleoporin 98kDa |
| GPC3 | glypican 3 |
| FOXP1 | forkhead box P1 |
| STAT6 | signal transducer and activator of transcription 6, interleukin-4 induced |
| ETNK1 | ethanolamine kinase 1 |
| CD79A | CD79a molecule, immunoglobulin-associated alpha |
| MB21D2 | Mab-21 domain containing 2 |
| SMAD2 | SMAD family member 2 |
| MECOM | MDS1 and EVI1 complex locus |
| AKT1 | v-akt murine thymoma viral oncogene homolog 1 |
| PREX2 | phosphatidylinositol-3,4,5-trisphosphate dependent Rac exchange factor 2 |
| KAT6A | K(lysine) acetyltransferase 6A |
| DDB2 | damage-specific DNA binding protein 2 |
| EED | embryonic ectoderm development |
| STRN | striatin, calmodulin binding protein |
| CCR4 | C-C motif chemokine receptor 4 |
| COL1A1 | collagen, type I, alpha 1 |
| CANT1 | calcium activated nucleotidase 1 |
| FLI1 | Friend leukemia virus integration 1 |
| EWSR1 | Ewing sarcoma breakpoint region 1 (EWS) |
| RUNX1 | runt-related transcription factor 1 (AML1) |
| NF2 | neurofibromatosis type 2 gene |
| NAB2 | NGFI-A binding protein 2 |
| BCL7A | B-cell CLL/lymphoma 7A |
| SETD1B | SET domain containing 1B |
| FOXR1 | forkhead box R1 |
| GATA1 | GATA binding protein 1 (globin transcription factor 1) |
| DGCR8 | DGCR8, microprocessor complex subunit |
| RARA | retinoic acid receptor, alpha |
| ETV1 | ets variant gene 1 |
| TFG | TRK-fused gene |
| LPP | LIM domain containing preferred translocation partner in lipoma |
| BAP1 | BRCA1 associated protein-1 (ubiquitin carboxy-terminal hydrolase) |
| TERT | telomerase reverse transcriptase |
| SMAD4 | SMAD family member 4 |
| RUNX1T1 | runt-related transcription factor 1; translocated to, 1 (cyclin D-related) |
| PLAG1 | pleiomorphic adenoma gene 1 |
| TSC2 | tuberous sclerosis 2 gene |
| RMI2 | RecQ mediated genome instability 2 |
| ERCC4 | excision repair cross-complementing rodent repair deficiency, complementation group 4 |
| XPA | xeroderma pigmentosum, complementation group A |
| PTCH1 | Homolog of Drosophila Patched gene |
| PML | promyelocytic leukemia |
| FEV | FEV protein - (HSRNAFEV) |
| CTCF | CCCTC-binding factor |
| A1CF | APOBEC1 complementation factor |
| PRKCB | protein kinase C beta |
| KTN1 | kinectin 1 (kinesin receptor) |
| ESR1 | estrogen receptor 1 |
| ARHGEF12 | RHO guanine nucleotide exchange factor (GEF) 12 (LARG) |
| ZNF331 | zinc finger protein 331 |
| CYSLTR2 | cysteinyl leukotriene receptor 2 |
| CLIP1 | CAP-GLY domain containing linker protein 1 |
| LASP1 | LIM and SH3 protein 1 |
| SH3GL1 | SH3-domain GRB2-like 1 (EEN) |
| NCKIPSD | NCK interacting protein with SH3 domain |
| DCTN1 | dynactin 1 |
| PAFAH1B2 | platelet-activating factor acetylhydrolase, isoform Ib, beta subunit 30kDa |
| BCORL1 | BCL6 corepressor-like 1 |
| LMNA | lamin A/C |
| RFWD3 | ring finger and WD repeat domain 3 |
| SND1 | staphylococcal nuclease and tudor domain containing 1 |
| TSC1 | tuberous sclerosis 1 gene |
| SMAD3 | SMAD family member 3 |
| COX6C | cytochrome c oxidase subunit VIc |
| DNAJB1 | DnaJ heat shock protein family (Hsp40) member B1 |
| CARD11 | caspase recruitment domain family, member 11 |
| HOXA11 | homeo box A11 |
| PPM1D | protein phosphatase, Mg2+/Mn2+ dependent 1D |
| FANCF | Fanconi anemia, complementation group F |
| KDM5A | lysine (K)-specific demethylase 5A, JARID1A |
| MGMT | O-6-methylguanine-DNA methyltransferase |
| KDM5C | lysine (K)-specific demethylase 5C (JARID1C) |
| ZNF384 | zinc finger protein 384 (CIZ/NMP4) |
| FH | fumarate hydratase |
| TRIP11 | thyroid hormone receptor interactor 11 |
| CCNB1IP1 | cyclin B1 interacting protein 1, E3 ubiquitin protein ligase |
| BCL11A | B-cell CLL/lymphoma 11A |
| TCL1A | T-cell leukemia/lymphoma 1A |
| TPM4 | tropomyosin 4 |
| LSM14A | LSM14A, SCD6 homolog A (S. cerevisiae) |
| PIK3R1 | phosphoinositide-3-kinase, regulatory subunit 1 (alpha) |
| CBLB | Cas-Br-M (murine) ecotropic retroviral transforming sequence b |
| MLLT1 | myeloid/lymphoid or mixed-lineage leukemia (trithorax homolog, Drosophila); translocated to, 1 (ENL) |
| KCNJ5 | potassium inwardly-rectifying channel; subfamily J; member 5 |
| CASP8 | caspase 8, apoptosis-related cysteine peptidase |
| CUX1 | cut-like homeobox 1 |
| PRCC | papillary renal cell carcinoma (translocation-associated) |
| CDKN2A | cyclin-dependent kinase inhibitor 2A (p16(INK4a)) gene |
| CRTC3 | CREB regulated transcription coactivator 3 |
| NFKB2 | nuclear factor of kappa light polypeptide gene enhancer in B-cells 2 (p49/p100) |
| GRM3 | glutamate metabotropic receptor 3 |
| GATA3 | GATA binding protein 3 |
| RPL10 | ribosomal protein L10 |
| FOXO4 | forkhead box O4 |
| CSF3R | colony stimulating factor 3 receptor (granulocyte) |
| ZMYM3 | zinc finger MYM-type containing 3 |
| DCAF12L2 | DDB1 and CUL4 associated factor 12 like 2 |
| MAPK1 | mitogen-activated protein kinase 1 |
| TNFRSF14 | tumor necrosis factor receptor superfamily, member 14 (herpesvirus entry mediator) |
| PRDM16 | PR domain containing 16 |
| CIC | capicua homolog |
| KDM6A | lysine (K)-specific demethylase 6A, UTX |
| RBM10 | RNA binding motif protein 10 |
| BCOR | BCL6 corepressor |
| CNOT3 | CCR4-NOT transcription complex subunit 3 |
| MSN | moesin |
| TFE3 | transcription factor binding to IGHM enhancer 3 |
| CRLF2 | cytokine receptor-like factor 2 |
| TAL1 | T-cell acute lymphocytic leukemia 1 (SCL) |
| PABPC1 | poly(A) binding protein cytoplasmic 1 |
| TCEA1 | transcription elongation factor A (SII), 1 |
| NBN | nibrin |
| HEY1 | hairy/enhancer-of-split related with YRPW motif 1 |
| OMD | osteomodulin |
| CD28 | CD28 molecule |
| ITGAV | integrin subunit alpha V |
| TCF7L2 | transcription factor 7-like 2 |
| VTI1A | vesicle transport through interaction with t-SNAREs homolog 1A |
| FAS | Fas cell surface death receptor |
| WT1 | Wilms tumour 1 gene |
| PAX8 | paired box gene 8 |
| MITF | melanogenesis-associated transcription factor |
| ROBO2 | roundabout guidance receptor 2 |
| LEF1 | lymphoid enhancer binding factor 1 |
| SOX2 | SRY (sex determining region Y)-box 2 |
| BCL6 | B-cell CLL/lymphoma 6 |
| TFRC | transferrin receptor (p90, CD71) |
| TP63 | tumor protein p63 |
| SLC34A2 | solute carrier family 34 (sodium phosphate), member 2 |
| RSPO3 | R-spondin 3 |
| BMP5 | bone morphogenetic protein 5 |
| CCNC | cyclin C |
| ACKR3 | atypical chemokine receptor 3 |
| MACC1 | MET transcriptional regulator MACC1 |
| PWWP2A | PWWP domain containing 2A |
| CCND3 | cyclin D3 |
| NFKBIE | nuclear factor of kappa light polypeptide gene enhancer in B-cells inhibitor, epsilon |
| MYD88 | myeloid differentiation primary response gene (88) |
| FCRL4 | Fc receptor-like 4 |
| FUS | fusion, derived from t(12;16) malignant liposarcoma |
| TP53 | tumor protein p53 |
| FCGR2B | Fc fragment of IgG, low affinity IIb, receptor for (CD32) |
| PALB2 | partner and localizer of BRCA2 |
| AXIN1 | axin 1 |
| FUBP1 | far upstream element (FUSE) binding protein 1 |
| DCC | DCC netrin 1 receptor |
| CRTC1 | CREB regulated transcription coactivator 1 |
| JUN | jun oncogene |
| CEBPA | CCAAT/enhancer binding protein (C/EBP), alpha |
| CEP89 | centrosomal protein 89kDa |
| CCNE1 | cyclin E1 |
| HLF | hepatic leukemia factor |
| CLTC | clathrin, heavy polypeptide (Hc) |
| CD79B | CD79b molecule, immunoglobulin-associated beta |
| CCR7 | C-C motif chemokine receptor 7 |
| BRCA1 | familial breast/ovarian cancer gene 1 |
| ETV6 | ets variant gene 6 (TEL oncogene) |
| CCND2 | cyclin D2 |
| MYCN | v-myc myelocytomatosis viral related oncogene, neuroblastoma derived (avian) |
| LRIG3 | leucine-rich repeats and immunoglobulin-like domains 3 |
| GLI1 | GLI family zinc finger 1 |
| CCND1 | cyclin D1 |
| GPHN | gephyrin (GPH) |
| SLC45A3 | solute carrier family 45, member 3 |
| NKX2-1 | NK2 homeobox 1 |
| NIN | ninein (GSK3B interacting protein) |
| TCF12 | transcription factor 12 (HTF4, helix-loop-helix transcription factors 4) |
| BCL11B | B-cell CLL/lymphoma 11B (CTIP2) |
| WIF1 | WNT inhibitory factor 1 |
| HNF1A | HNF1 homeobox A |
| BTG1 | B-cell translocation gene 1, anti-proliferative |
| LCP1 | lymphocyte cytosolic protein 1 (L-plastin) |
| FOXO1 | forkhead box O1 |

**Table S6.** Predicted neoplasm comorbidity enriched MOA proteins that are labeled as cancer associated in the COSMIC(30) database using GWAS risk genes(2) as input.

| **Gene name** | **Gene description** |
| --- | --- |
| NR4A3 | nuclear receptor subfamily 4, group A, member 3 (NOR1) |
| AR | Androgen Receptor |
| ACKR3 | atypical chemokine receptor 3 |
| IL21R | interleukin 21 receptor |
| IL7R | interleukin 7 receptor |
| FKBP9 | FK506 binding protein 9 |
| EML4 | echinoderm microtubule associated protein like 4 |
| SF3B1 | splicing factor 3b, subunit 1, 155kDa |
| PPP6C | protein phosphatase 6, catalytic subunit |
| CCR7 | C-C motif chemokine receptor 7 |
| PPP2R1A | protein phosphatase 2, regulatory subunit A, alpha |
| SMAD2 | SMAD family member 2 |
| TSHR | thyroid stimulating hormone receptor |
| PIK3R1 | phosphoinositide-3-kinase, regulatory subunit 1 (alpha) |
| CHD2 | chromodomain helicase DNA binding protein 2 |
| MYD88 | myeloid differentiation primary response gene (88) |
| SH2B3 | SH2B adaptor protein 3 |
| ERBB3 | erb-b2 receptor tyrosine kinase 3 |
| CCR4 | C-C motif chemokine receptor 4 |
| P2RY8 | purinergic receptor P2Y, G-protein coupled, 8 |
| CYSLTR2 | cysteinyl leukotriene receptor 2 |
| SMAD3 | SMAD family member 3 |
| ERBB4 | erb-b2 receptor tyrosine kinase 4 |
| TRIP11 | thyroid hormone receptor interactor 11 |
| ERBB2 | v-erb-b2 erythroblastic leukemia viral oncogene homolog 2, neuro/glioblastoma derived oncogene homolog (avian) |
| NF1 | neurofibromatosis type 1 gene |
| GMPS | guanine monphosphate synthetase |
| KTN1 | kinectin 1 (kinesin receptor) |
| CLIP1 | CAP-GLY domain containing linker protein 1 |
| LMO2 | LIM domain only 2 (rhombotin-like 1) (RBTN2) |
| COX6C | cytochrome c oxidase subunit VIc |
| JAK2 | Janus kinase 2 |
| ANK1 | ankyrin 1 |
| HOOK3 | hook homolog 3 |
| CHCHD7 | coiled-coil-helix-coiled-coil-helix domain containing 7 |
| TSC1 | tuberous sclerosis 1 gene |
| PRDM2 | PR/SET domain 2 |
| JAK1 | Janus kinase 1 |
| BCORL1 | BCL6 corepressor-like 1 |
| MGMT | O-6-methylguanine-DNA methyltransferase |
| ELN | elastin |
| DCTN1 | dynactin 1 |
| MET | met proto-oncogene (hepatocyte growth factor receptor) |
| PAFAH1B2 | platelet-activating factor acetylhydrolase, isoform Ib, beta subunit 30kDa |
| CUX1 | cut-like homeobox 1 |
| JAK3 | Janus kinase 3 |
| HOXA11 | homeo box A11 |
| EGFR | epidermal growth factor receptor (erythroblastic leukemia viral (v-erb-b) oncogene homolog, avian) |
| FGFR2 | fibroblast growth factor receptor 2 |
| NTRK3 | neurotrophic tyrosine kinase, receptor, type 3 |
| ABL1 | v-abl Abelson murine leukemia viral oncogene homolog 1 |
| FGFR1 | fibroblast growth factor receptor 1 |
| MYO5A | myosin VA (heavy chain 12, myoxin) |
| FES | FES proto-oncogene, tyrosine kinase |
| BMPR1A | bone morphogenetic protein receptor, type IA |
| RET | ret proto-oncogene |
| CDK4 | cyclin-dependent kinase 4 |
| FGFR4 | fibroblast growth factor receptor 4 |
| DDX6 | DEAD (Asp-Glu-Ala-Asp) box polypeptide 6 |
| SND1 | staphylococcal nuclease and tudor domain containing 1 |
| FLT4 | fms-related tyrosine kinase 4 |
| BRAF | v-raf murine sarcoma viral oncogene homolog B1 |
| CARS | cysteinyl-tRNA synthetase |
| FLT3 | fms-related tyrosine kinase 3 |
| EPHA7 | EPH receptor A7 |
| CSF1R | colony stimulating factor 1 receptor |
| PDGFRB | platelet-derived growth factor receptor, beta polypeptide |
| ACVR1 | activin A receptor, type I |
| ACVR2A | activin A receptor type 2A |
| FGFR3 | fibroblast growth factor receptor 3 |
| CHEK2 | CHK2 checkpoint homolog (S. pombe) |
| MAP3K13 | mitogen-activated protein kinase kinase kinase 13 |
| CDK12 | cyclin-dependent kinase 12 |
| TGFBR2 | transforming growth factor beta receptor II |
| BCL2 | B-cell CLL/lymphoma 2 |
| CASP8 | caspase 8, apoptosis-related cysteine peptidase |
| BTK | Bruton agammaglobulinemia tyrosine kinase |
| CEP89 | centrosomal protein 89kDa |
| EPHA3 | EPH receptor A3 |
| NTRK1 | neurotrophic tyrosine kinase, receptor, type 1 |
| ALK | anaplastic lymphoma kinase (Ki-1) |
| MYH11 | myosin, heavy polypeptide 11, smooth muscle |
| DDR2 | discoidin domain receptor 2 |
| PDGFRA | platelet-derived growth factor, alpha-receptor |
| KIT | v-kit Hardy-Zuckerman 4 feline sarcoma viral oncogene homolog |
| KDR | vascular endothelial growth factor receptor 2 |
| ESR1 | estrogen receptor 1 |
| PHF6 | PHD finger protein 6 |
| PTK6 | protein tyrosine kinase 6 |
| MAP2K2 | mitogen-activated protein kinase kinase 2 |
| CDK6 | cyclin-dependent kinase 6 |
| KCNJ5 | potassium inwardly-rectifying channel; subfamily J; member 5 |
| CTNNA2 | catenin alpha 2 |
| MAP2K4 | mitogen-activated protein kinase kinase 4 |
| TEC | tec protein tyrosine kinase |
| RARA | retinoic acid receptor, alpha |
| ABL2 | c-abl oncogene 2, non-receptor tyrosine kinase |
| MAP3K1 | mitogen-activated protein kinase kinase kinase 1, E3 ubiquitin protein ligase |
| MAP2K1 | mitogen-activated protein kinase kinase 1 |
| PIM1 | pim-1 oncogene |
| STK11 | serine/threonine kinase 11 gene (LKB1) |
| SYK | spleen tyrosine kinase |
| HRAS | v-Ha-ras Harvey rat sarcoma viral oncogene homolog |
| ARAF | A-Raf proto-oncogene, serine/threonine kinase |
| SEPT6 | septin 6 |
| MYH9 | myosin, heavy polypeptide 9, non-muscle |
| ARHGAP5 | Rho GTPase activating protein 5 |
| PPM1D | protein phosphatase, Mg2+/Mn2+ dependent 1D |
| SEPT9 | septin 9 |
| PRKAR1A | protein kinase, cAMP-dependent, regulatory, type I, alpha (tissue specific extinguisher 1) |
| BUB1B | BUB1 budding uninhibited by benzimidazoles 1 homolog beta (yeast) |
| ZNF384 | zinc finger protein 384 (CIZ/NMP4) |
| ERG | v-ets erythroblastosis virus E26 oncogene like (avian) |
| 5-Sep | septin 5 |
| GNAS | guanine nucleotide binding protein (G protein), alpha stimulating activity polypeptide 1 |
| DNM2 | dynamin 2 |
| PRKACA | protein kinase cAMP-activated catalytic subunit alpha |
| GNA11 | guanine nucleotide binding protein (G protein), alpha 11 (Gq class) |
| AKT2 | v-akt murine thymoma viral oncogene homolog 2 |
| KRAS | v-Ki-ras2 Kirsten rat sarcoma 2 viral oncogene homolog |
| SGK1 | serum/glucocorticoid regulated kinase 1 |
| RAC1 | ras-related C3 botulinum toxin substrate 1 (rho family, small GTP binding protein Rac1) |
| BCL10 | B-cell CLL/lymphoma 10 |
| NRAS | neuroblastoma RAS viral (v-ras) oncogene homolog |
| GRM3 | glutamate metabotropic receptor 3 |
| RHOA | ras homolog family member A |
| RHOH | ras homolog family member H |
| ITK | IL2-inducible T-cell kinase |
| CREB1 | cAMP responsive element binding protein 1 |
| ATIC | 5-aminoimidazole-4-carboxamide ribonucleotide formyltransferase/IMP cyclohydrolase |
| WRN | Werner syndrome (RECQL2) |
| PABPC1 | poly(A) binding protein cytoplasmic 1 |
| GNAQ | guanine nucleotide binding protein (G protein), q polypeptide |
| ERCC3 | excision repair cross-complementing rodent repair deficiency, complementation group 3 (xeroderma pigmentosum group B complementing) |
| DNAJB1 | DnaJ heat shock protein family (Hsp40) member B1 |
| ITGAV | integrin subunit alpha V |
| IDH1 | isocitrate dehydrogenase 1 (NADP+), soluble |
| ZNF331 | zinc finger protein 331 |
| BCL11A | B-cell CLL/lymphoma 11A |
| STRN | striatin, calmodulin binding protein |
| MSH6 | mutS homolog 6 (E. coli) |
| MSH2 | mutS homolog 2 (E. coli) |
| POLD1 | DNA polymerase delta 1, catalytic subunit |
| CBLB | Cas-Br-M (murine) ecotropic retroviral transforming sequence b |
| DDX5 | DEAD (Asp-Glu-Ala-Asp) box polypeptide 5 |
| FHIT | fragile histidine triad gene |
| KAT7 | lysine acetyltransferase 7 |
| CLTC | clathrin, heavy polypeptide (Hc) |
| RAF1 | v-raf-1 murine leukemia viral oncogene homolog 1 |
| ACSL3 | acyl-CoA synthetase long-chain family member 3 |
| KDSR | 3-ketodihydrosphingosine reductase |
| GPC3 | glypican 3 |
| DCAF12L2 | DDB1 and CUL4 associated factor 12 like 2 |
| DDX3X | DEAD-box helicase 3, X-linked |
| BCOR | BCL6 corepressor |
| LMNA | lamin A/C |
| MLLT11 | myeloid/lymphoid or mixed-lineage leukemia (trithorax homolog, Drosophila); translocated to, 11 |
| SETDB1 | SET domain bifurcated 1 |
| MSN | moesin |
| KDM5C | lysine (K)-specific demethylase 5C (JARID1C) |
| SMC1A | structural maintenance of chromosomes 1A |
| HLA-A | major histocompatibility complex, class I, A |
| RPL10 | ribosomal protein L10 |
| EPS15 | epidermal growth factor receptor pathway substrate 15 (AF1p) |
| AKT3 | v-akt murine thymoma viral oncogene homolog 3 |
| DNMT3A | DNA (cytosine-5-)-methyltransferase 3 alpha |
| TOP1 | topoisomerase (DNA) I |
| SRC | SRC proto-oncogene, non-receptor tyrosine kinase |
| LCK | lymphocyte-specific protein tyrosine kinase |
| SLC45A3 | solute carrier family 45, member 3 |
| NF2 | neurofibromatosis type 2 gene |
| KDM5A | lysine (K)-specific demethylase 5A, JARID1A |
| POT1 | protection of telomeres 1 |
| EED | embryonic ectoderm development |
| DDX10 | DEAD (Asp-Glu-Ala-Asp) box polypeptide 10 |
| ALDH2 | aldehyde dehydrogenase 2 family (mitochondrial) |
| PRDM1 | PR domain containing 1, with ZNF domain |
| LATS2 | large tumor suppressor kinase 2 |
| NFKBIE | nuclear factor of kappa light polypeptide gene enhancer in B-cells inhibitor, epsilon |
| BMP5 | bone morphogenetic protein 5 |
| BTG1 | B-cell translocation gene 1, anti-proliferative |
| CARD11 | caspase recruitment domain family, member 11 |
| PMS2 | PMS2 postmeiotic segregation increased 2 (S. cerevisiae) |
| LATS1 | large tumor suppressor kinase 1 |
| DDIT3 | DNA-damage-inducible transcript 3 |
| VTI1A | vesicle transport through interaction with t-SNAREs homolog 1A |
| KIF5B | kinesin family member 5B |
| NRG1 | neuregulin 1 |
| CTNND1 | catenin delta 1 |
| EZH2 | enhancer of zeste homolog 2 |
| CNBD1 | cyclic nucleotide binding domain containing 1 |
| RECQL4 | RecQ protein-like 4 |
| DDB2 | damage-specific DNA binding protein 2 |
| CYLD | familial cylindromatosis gene |
| RAD17 | RAD17 checkpoint clamp loader component |
| PALB2 | partner and localizer of BRCA2 |
| CIITA | class II, major histocompatibility complex, transactivator |
| EIF4A2 | eukaryotic translation initiation factor 4A, isoform 2 |
| RAP1GDS1 | RAP1, GTP-GDP dissociation stimulator 1 |
| NIN | ninein (GSK3B interacting protein) |
| RAD51B | RAD51 paralog B |
| ACSL6 | acyl-CoA synthetase long-chain family member 6 |
| IDH2 | isocitrate dehydrogenase 2 (NADP+), mitochondrial |
| BLM | Bloom Syndrome |
| TCL1A | T-cell leukemia/lymphoma 1A |
| DICER1 | dicer 1, ribonuclease type III |
| KNSTRN | kinetochore localized astrin/SPAG5 binding protein |
| AKT1 | v-akt murine thymoma viral oncogene homolog 1 |

**Table S7.** COVID-19 differentially expressed genes from(31) ranked by their adjusted p-value (padj) that are mapped to the COSMIC(30) database.

| **Gene Name** | **Gene Description** | **padj** | **log2FoldChange** | **Up/down-regulated** |
| --- | --- | --- | --- | --- |
| NBN | nibrin | 2E-09 | 4.26 | Up-regulated |
| LCP1 | lymphocyte cytosolic protein 1 (L-plastin) | 2E-09 | 3.72 | Up-regulated |
| WAS | Wiskott-Aldrich syndrome | 1E-08 | 4.63 | Up-regulated |
| RHOH | ras homolog family member H | 4E-07 | 4.72 | Up-regulated |
| CSF3R | colony stimulating factor 3 receptor (granulocyte) | 4E-06 | 4.01 | Up-regulated |
| JUN | jun oncogene | 2E-05 | -3.57 | Down-regulated |
| BCL3 | B-cell CLL/lymphoma 3 | 6E-05 | 2.85 | Up-regulated |
| BAP1 | BRCA1 associated protein-1 (ubiquitin carboxy-terminal hydrolase) | 2E-04 | -4.36 | Down-regulated |
| PER1 | period homolog 1 (Drosophila) | 4E-04 | -4.34 | Down-regulated |
| RALGDS | ral guanine nucleotide dissociation stimulator | 7E-04 | -3.80 | Down-regulated |
| XPC | xeroderma pigmentosum, complementation group C | 1E-03 | -3.98 | Down-regulated |
| GATA2 | GATA binding protein 2 | 2E-03 | -3.95 | Down-regulated |
| NDRG1 | N-myc downstream regulated 1 | 2E-03 | -2.72 | Down-regulated |
| ATRX | alpha thalassemia/mental retardation syndrome X-linked | 2E-03 | -3.83 | Down-regulated |
| CTCF | CCCTC-binding factor | 3E-03 | -3.80 | Down-regulated |
| BCOR | BCL6 corepressor | 3E-03 | -3.82 | Down-regulated |
| TET2 | tet oncogene family member 2 | 3E-03 | 2.27 | Up-regulated |
| CD74 | CD74 molecule, major histocompatibility complex, class II invariant chain | 3E-03 | 2.34 | Up-regulated |
| DDX5 | DEAD (Asp-Glu-Ala-Asp) box polypeptide 5 | 3E-03 | -2.00 | Down-regulated |
| BCR | breakpoint cluster region | 4E-03 | -3.66 | Down-regulated |
| TGFBR2 | transforming growth factor beta receptor II | 4E-03 | -2.52 | Down-regulated |
| LZTR1 | leucine-zipper-like transcription regulator 1 | 5E-03 | -3.66 | Down-regulated |
| JAK3 | Janus kinase 3 | 5E-03 | 2.41 | Up-regulated |
| NFATC2 | nuclear factor of activated T-cells, cytoplasmic, calcineurin-dependent 2 | 5E-03 | -3.60 | Down-regulated |
| MLH1 | E.coli MutL homolog gene | 6E-03 | -3.56 | Down-regulated |
| ZMYM2 | zinc finger protein 198 | 6E-03 | -3.21 | Down-regulated |
| NAB2 | NGFI-A binding protein 2 | 6E-03 | -3.55 | Down-regulated |
| CANT1 | calcium activated nucleotidase 1 | 8E-03 | -3.14 | Down-regulated |
| TSC1 | tuberous sclerosis 1 gene | 8E-03 | -3.21 | Down-regulated |
| NF2 | neurofibromatosis type 2 gene | 9E-03 | -3.16 | Down-regulated |
| PML | promyelocytic leukemia | 1E-02 | 1.86 | Up-regulated |
| ACVR1 | activin A receptor, type I | 2E-02 | -3.18 | Down-regulated |
| VHL | von Hippel-Lindau syndrome gene | 2E-02 | 2.11 | Up-regulated |
| LMNA | lamin A/C | 2E-02 | -2.48 | Down-regulated |
| POU2AF1 | POU domain, class 2, associating factor 1 (OBF1) | 2E-02 | 2.75 | Up-regulated |
| DDB2 | damage-specific DNA binding protein 2 | 2E-02 | -3.16 | Down-regulated |
| RBM10 | RNA binding motif protein 10 | 2E-02 | -2.34 | Down-regulated |
| ATP1A1 | ATPase, Na+/K+ transporting, alpha 1 polypeptide | 2E-02 | -2.04 | Down-regulated |
| TMPRSS2 | transmembrane protease, serine 2 | 2E-02 | -2.89 | Down-regulated |
| HLA-A | major histocompatibility complex, class I, A | 2E-02 | 2.00 | Up-regulated |
| TNFRSF14 | tumor necrosis factor receptor superfamily, member 14 (herpesvirus entry mediator) | 2E-02 | -2.22 | Down-regulated |
| YWHAE | tyrosine 3-monooxygenase/tryptophan 5-monooxygenase activation protein, epsilon polypeptide (14-3-3 epsilon) | 2E-02 | 1.66 | Up-regulated |
| ARHGEF10L | Rho guanine nucleotide exchange factor 10 like | 2E-02 | -3.05 | Down-regulated |
| PWWP2A | PWWP domain containing 2A | 2E-02 | -3.05 | Down-regulated |
| NUMA1 | nuclear mitotic apparatus protein 1 | 2E-02 | -2.21 | Up-regulated |
| EZR | ezrin | 2E-02 | -1.95 | Down-regulated |
| APC | adenomatous polyposis of the colon gene | 3E-02 | -3.01 | Down-regulated |
| EED | embryonic ectoderm development | 3E-02 | -3.01 | Down-regulated |
| PLCG1 | phospholipase C, gamma 1 | 3E-02 | -2.60 | Down-regulated |
| IDH2 | isocitrate dehydrogenase 2 (NADP+), mitochondrial | 3E-02 | -2.40 | Down-regulated |
| SFRP4 | secreted frizzled related protein 4 | 3E-02 | -2.93 | Down-regulated |
| MAP3K13 | mitogen-activated protein kinase kinase kinase 13 | 3E-02 | 1.96 | Up-regulated |
| FES | FES proto-oncogene, tyrosine kinase | 3E-02 | -2.90 | Down-regulated |
| SGK1 | serum/glucocorticoid regulated kinase 1 | 3E-02 | -2.35 | Down-regulated |
| ID3 | inhibitor of DNA binding 3, HLH protein | 3E-02 | -2.34 | Down-regulated |
| NCKIPSD | NCK interacting protein with SH3 domain | 3E-02 | -2.78 | Down-regulated |
| CDK4 | cyclin-dependent kinase 4 | 3E-02 | 1.67 | Up-regulated |
| CALR | calreticulin | 4E-02 | 1.54 | Up-regulated |
| ZRSR2 | zinc finger (CCCH type), RNA-binding motif and serine/arginine rich 2 | 4E-02 | -2.86 | Down-regulated |
| TRRAP | transformation/transcription domain-associated protein | 4E-02 | -2.39 | Down-regulated |
| POLE | polymerase (DNA directed), epsilon, catalytic subunit | 4E-02 | -2.84 | Down-regulated |
| LMO2 | LIM domain only 2 (rhombotin-like 1) (RBTN2) | 4E-02 | 2.14 | Up-regulated |
| EPAS1 | endothelial PAS domain protein 1 | 4E-02 | -2.24 | Down-regulated |
| AXIN2 | axin 2 | 4E-02 | -2.82 | Down-regulated |
| AFF4 | AF4/FMR2 family, member 4 | 4E-02 | -2.06 | Down-regulated |
| PTPN13 | protein tyrosine phosphatase, non-receptor type 13 | 4E-02 | -2.80 | Down-regulated |
| ASXL1 | additional sex combs like 1 | 4E-02 | -2.08 | Down-regulated |
| CREB3L2 | cAMP responsive element binding protein 3-like 2 | 4E-02 | -2.33 | Down-regulated |
| SEPT9 | septin 9 | 4E-02 | -2.21 | Down-regulated |
| PDE4DIP | phosphodiesterase 4D interacting protein (myomegalin) | 4E-02 | -2.50 | Down-regulated |
| TRIP11 | thyroid hormone receptor interactor 11 | 4E-02 | -2.61 | Down-regulated |
| SMARCB1 | SWI/SNF related, matrix associated, actin dependent regulator of chromatin, subfamily b, member 1 | 4E-02 | -2.44 | Down-regulated |
| TPM4 | tropomyosin 4 | 5E-02 | 1.60 | Up-regulated |
| BRD3 | bromodomain containing 3 | 5E-02 | -2.74 | Down-regulated |
| SEPT6 | septin 6 | 5E-02 | 1.62 | Up-regulated |
| SPECC1 | sperm antigen with calponin homology and coiled-coil domains 1 | 5E-02 | -2.55 | Down-regulated |
| MLLT10 | myeloid/lymphoid or mixed-lineage leukemia (trithorax homolog, Drosophila); translocated to, 10 (AF10) | 5E-02 | -2.41 | Down-regulated |
| RPL10 | ribosomal protein L10 | 5E-02 | -1.56 | Down-regulated |
| FGFR1 | fibroblast growth factor receptor 1 | 5E-02 | -2.36 | Down-regulated |
| NOTCH2 | Notch homolog 2 | 5E-02 | -2.49 | Down-regulated |
| MYC | v-myc myelocytomatosis viral oncogene homolog (avian) | 5E-02 | -2.33 | Down-regulated |
| CHD4 | chromodomain helicase DNA binding protein 4 | 5E-02 | -1.75 | Down-regulated |
